# Variants in the proteasome regulator PSMF1 cause a phenotypic spectrum from early-onset Parkinson’s disease to perinatal lethality and disrupt mitochondrial function

**DOI:** 10.1101/2024.06.19.24308302

**Authors:** Francesca Magrinelli, Christelle Tesson, Plamena R. Angelova, Jose A. Rodriguez, Annarita Scardamaglia, Benjamin O’Callaghan, Simon A. Lowe, Ainara Salazar-Villacorta, Brian Hon-Yin Chung, Matthew Jaconelli, Barbara Vona, Noemi Esteras, Angela Mammana, Junko Shimazu, Anna Ka-Yee Kwong, Thomas Courtin, Shahryar Alavi, Reza Maroofian, Raja Nirujogi, Mariasavina Severino, Edoardo Monfrini, Clarissa Rocca, Patrick A. Lewis, Stephanie Efthymiou, Rebecca Buchert, Linda Sofan, Pawel Lis, Chloé Pinon, Guido J. Breedveld, Martin Man-Chun Chui, David Murphy, Vanessa Pitz, Mary B. Makarious, Simone Baiardi, Marina Volin, Marlene Cassar, Bassem A. Hassan, Sana Iftikhar, Peter Bauer, Michele Tinazzi, Marina Svetel, Bedia Samanci, Haşmet A. Hanağası, Basar Bilgiç, Francesco Cavallieri, Mario Santangelo, José A. Obeso, Monica M. Kurtis, Guillaume Cogan, Güneş Kiziltan, Tuğçe Gül-Demirkale, Hülya Tireli, Gülbün A. Yüksel, Gül Yalçın-Cakmakli, Bülent Elibol, Nina Barišić, Earny Wei-Sen Ng, Sze-Shing Fan, Tova Hershkovitz, Karin Weiss, Javeria Raza Alvi, Tipu Sultan, Issam Azmi Alkhawaja, Tawfiq Froukh, Abdollah E Hadeel Alrukban, Christine Fauth, Ulrich A. Schatz, Thomas Zöggeler, Michael Zech, Karen Stals, Vinod Varghese, Sonia Gandhi, Cornelis Blauwendraat, John A. Hardy, Alessio Di Fonzo, Vincenzo Bonifati, Tobias B. Haack, Aida M. Bertoli-Avella, Suzanne Lesage, Ayşe Nazlı Başak, Robert Steinfeld, Piero Parchi, James E.C. Jepson, Dario R. Alessi, PSMF1 Study Group, Alexis Brice, Hermann Steller, Andrey Y. Abramov, Kailash P. Bhatia, Henry Houlden

## Abstract

Dissecting biological pathways highlighted by Mendelian gene discovery has provided critical insights into the pathogenesis of Parkinson’s disease (PD) and neurodegeneration. This approach ultimately catalyzes the identification of potential biomarkers and therapeutic targets. Here, we identify *PSMF1* as a novel gene implicated in parkinsonism and childhood neurodegeneration. We find that biallelic *PSMF1* missense and loss-of-function variants co-segregate with phenotypes from early-onset PD to perinatal lethality with neurological manifestations across 17 pedigrees with 24 affected subjects, showing clear genotype-phenotype correlation. *PSMF1* encodes the proteasome regulator PSMF1/PI31, a highly conserved, ubiquitously expressed partner of the 20S proteasome and neurodegeneration-associated F-box-O 7 and valosin-containing proteins. We demonstrate that *PSMF1* variants impair mitochondrial membrane potential, dynamics and mitophagy, and may affect proteasomal abundance and assembly in patient-derived fibroblasts. Furthermore, *Drosophila* and mouse models of *PSMF1* loss of function exhibit age-dependent motor impairment, as well as brain-wide mitochondrial membrane depolarization and dopaminergic neurodegeneration in aged flies, and diffuse gliosis in mice. Collectively, our findings unequivocally link defective PSMF1 to early-onset parkinsonism and neurodegeneration, and suggest proteasomal and mitochondrial dysfunction as mechanistic contributors.

---

Parkinson’s disease (PD), the second-most prevalent neurodegenerative disorder worldwide, is projected to grow exponentially with no cure currently available.^1,2^ Notably, up to 15% of PD cases are linked to mono- and biallelic pathogenic or risk variants in single genes.^3^ Monogenic forms of PD and parkinsonism have unveiled crucial pathomechanisms that also underpin sporadic forms, highlighting a complex neurobiological network.^1,4^ This involves α-synuclein synthesis and dynamic equilibrium (*SNCA*), degradation of cytoplasmic components and organelles via the autophagy-endolysosomal system (*GBA1*, *LRRK2*, *RAB32*, *VPS35*), proteostasis through the ubiquitin-proteasome system (*PRKN*, *FBXO7*), mitochondrial functions (*PRKN*, *PINK1*, *FBXO7*), oxidative stress response (*DJ-1*) and the regulation of neuroinflammation (*LRRK2*).^1,4,5^ Disruption in these pathways culminates in the loss of dopaminergic neurons in the substantia nigra pars compacta, manifesting clinically as parkinsonism.^1,5^ Exploring pathways highlighted by Mendelian gene discovery fosters the identification of biomarkers and development of disease-modifying strategies for PD, as exemplified with LRRK2 inhibitors.^6-8^

Perturbations of the ubiquitin-proteasome system and mitochondrial processes are recognized as pivotal, interconnected drivers of neurodegeneration in PD, ultimately contributing to dopaminergic neuron vulnerability through proteotoxicity and bioenergetic failure.^9-13^ Within this framework, *PSMF1*, which encodes the proteasome regulator PSMF1/PI31,^14,15^ emerges as a new key player.

PSMF1 is a highly conserved 31-kDa protein which is ubiquitously expressed across human tissues.^15,16^ *In vitro*, PSMF1 inhibits peptide hydrolysis by the 20S proteasome through its C-terminal proline-rich domain, via either direct binding or competitive binding to the 20S activating particles PA700 and PA28.^15,17^ However, PSMF1 can also stimulate 26S proteasome-mediated proteolysis *in vitro*.^18^ *In vivo* studies demonstrate a biological function of PSMF1 in promoting protein breakdown. PSMF1 can stimulate 26S assembly,^19^ and *Psmf1* inactivation in fruit flies (*Drosophila melanogaster*) and mice (*Mus musculus*) causes accumulation of poly-ubiquitinated aggregates, which are exclusive substrates of the 26S proteasome.^18-21^ Furthermore, PSMF1 promotes proteolysis in *Drosophila*, mouse, yeast and plants.^18,22-24^ Finally, it mediates fast transport of proteasomes between neurosomes and synapses and is required for synapse maintenance and neuronal survival in *Drosophila* and mice.^20,22^ Notably, PSMF1 interacts with key proteins implicated in human neurodegeneration. For instance, PSMF1 is a high-affinity binding partner of F-box only protein 7 (FBXO7), whose genetic defects cause juvenile-onset PD/parkinsonism,^25^ through their respective N-terminal <u>F</u>BXO7/<u>P</u>SMF1 (FP) domain.^26^ In human fibroblasts, the ablation of this heterodimer by an *FBXO7* missense variant leads to reduced expression and stability of PSMF1 and FBXO7 as well as proteasomal and mitochondrial dysfunction.^27^ Intriguingly, FBXO7 prevents PSMF1 from proteolytic cleavage in *Drosophila* and mouse, suggesting that FBXO7 loss may cause proteasomal impairment by PSMF1 inactivation.^18,28^ Additionally, PSMF1 directly binds valosin-containing protein (VCP),^29^ whose defective functions are linked to amyotrophic lateral sclerosis, frontotemporal dementia and Huntington disease,^30,31^ with VCP and PSMF1 acting as *in vitro* up- and down-regulator of proteasomal activity, respectively.^29^ Finally, *PSMF1* variants have also been associated with Alzheimer’s disease.^32,33^

This study identifies *PSMF1* as a novel gene implicated in PD, parkinsonism and early human neurodegeneration. By extensive data mining and functional validation, we establish that biallelic *PSMF1* missense and loss-of-function variants cause a phenotypic spectrum ranging from early-onset PD to perinatal lethality with neurological manifestations, with clear genotype-phenotype correlation. We explore the organellar and cellular consequences of PSMF1 deficiency *in vitro* by performing live imaging of mitochondria in patient-derived cultured fibroblasts, and proteasome assays of their protein lysates. Finally, we explore its pathological consequences *in vivo* and *ex vivo* in *Drosophila* and mouse models.

## RESULTS

### Identification of biallelic *PSMF1* variants in affected subjects from 17 ethnically diverse families

In a nonconsanguineous family (Pedigree A; Fig.1a) with the male proband (A-II-2) affected by early-onset parkinsonism, patient-parent trio exome sequencing (ES) revealed he was compound heterozygote for one missense variant (c.724C>G p.Arg242Gly) and one splice variant (c.282+2T>A) in the candidate gene *PSMF1* (NM_006814.5). Through initial data sharing, we identified a consanguineous family (Pedigree B; Fig.1a) with two siblings (B-II-3; B-II-4) manifesting with early-onset PD and harboring the same *PSMF1* missense variant detected in Pedigree A in the homozygous state.^34^ To replicate the association of biallelic *PSMF1* variants with PD/parkinsonism and further delineate *PSMF1*-related disorder, we applied a genotype-first approach by interrogating large next-generation sequencing datasets from collaborative networks and platforms, including Queen Square Genomics (Pedigrees C-I-L-M), several diagnostic and research genetic laboratories worldwide (Pedigrees C-D-E-F-G-H-M-O), 100,000 Genomes Project, UK Biobank (UKB), The Accelerating Medicines Partnership program for Parkinson’s disease (AMP-PD), Solve-RD, CENTOGENE (Pedigrees B-D-N), GeneDx, Baylor Genetics, Genesis (Pedigree F), ClinVar, VarSome, Lifera Omics (Pedigree N), GeneMatcher (Pedigrees J-K-P-Q).^35^ Pedigrees identified from multiple sources were deduplicated.

**Figure 1.**
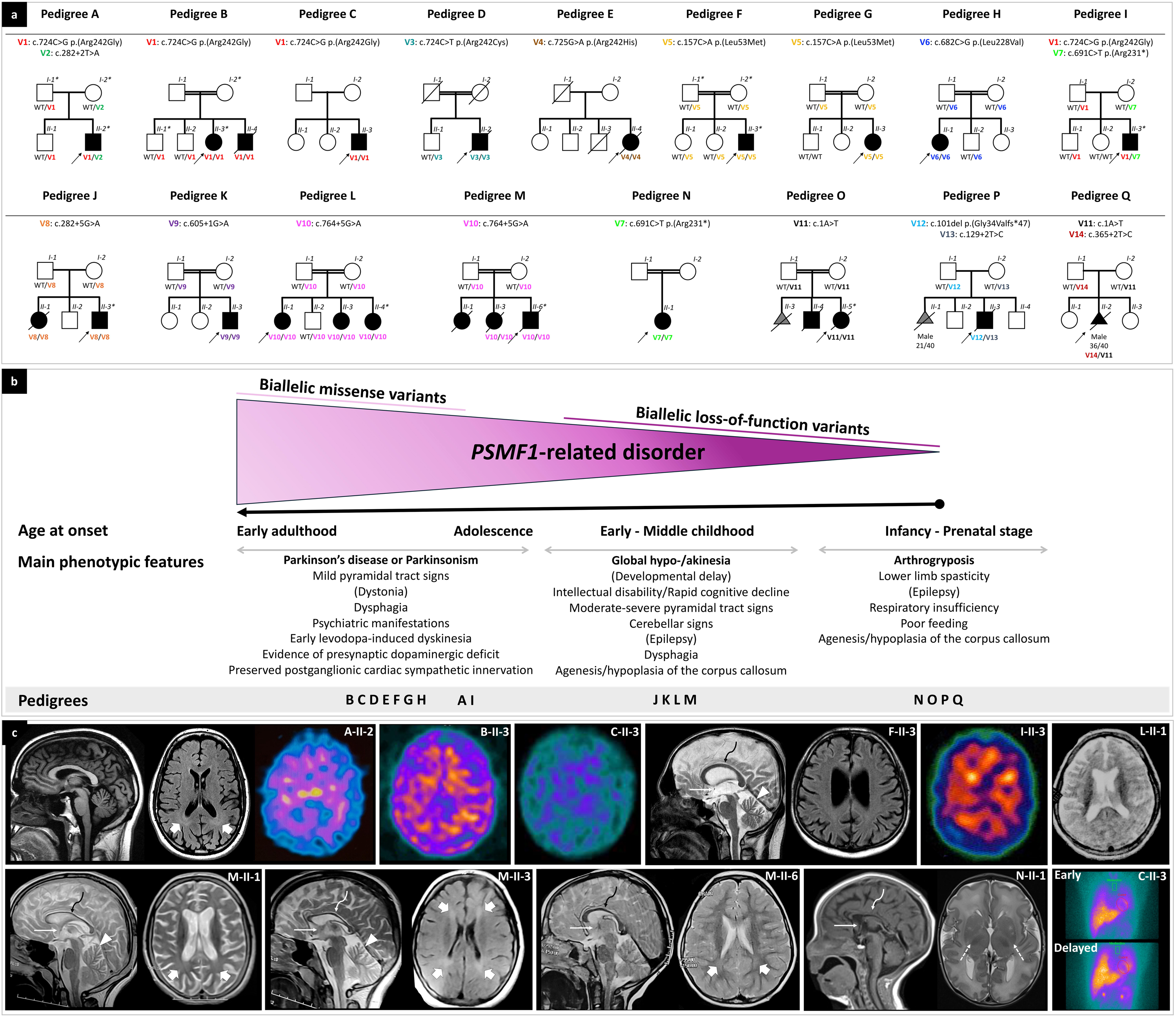
Pedigrees and phenotypic features of individuals with biallelic *PSMF1* variants. **(a) Simplified pedigrees of *PSMF1* families.** Pedigrees of 17 *PSMF1* families are presented, with the results of segregation analysis of *PSMF1* variants. In the diagrams, males are represented by squares, females by circles, and fetuses by triangles. A diagonal line across a symbol indicates a deceased individual. Fetal gender and gestational age at death (*n* weeks/40 weeks) are reported below their symbol, if available. Consanguineous marriages are denoted by double lines between symbols, while probands are marked with an arrow. Individuals exhibiting a neurological disease phenotype are represented by black filled symbols. Fetuses aborted for undetermined cause (O-II-3, P-II-1) are represented by a gray filled symbol. Roman numerals indicate generations, whereas Arabic numbers denote individuals within each generation. Variants are referenced to *PSMF1* transcript NM_006814.5 and displayed with different font colors (Fig. 2a; Supplementary File 1). “WT” designates the *PSMF1* wild-type allele. An asterisk identifies dermal fibroblast donors. **(b) Genotype-phenotype correlation in *PSMF1*-related disorder.** Schematic representation of the core phenotypic features associated with biallelic variants in *PSMF1*, with indication of age at onset and main clinical manifestations in subgroups of subjects belonging to the study cohort. Increasingly more severe phenotypes correspond to progressively more deleterious mutational effects. **(c) Neuroimaging and ¹²³I-MIBG myocardial scintigraphy features of *PSMF1*-related disorder.** Brain MRI with sagittal T1- or T2-weighted images and axial FLAIR or T2-weighted images of individuals A-II-2 (25-30 years), F-II-3 (25-30 years), L-II-1 (10-15 years), M-II-1 (10-15 years), M-II-3 (10-15 years), M-II-6 (0-5 years) and N-II-1 (neonatal period). Mild-to-moderate enlargement of the cerebral subarachnoid spaces is noted in subjects A-II-2, F-II-3, L-II-1, M-II-1 and M-II-3. Enlargement of the subarachnoid spaces of the superior cerebellar vermis is present in individuals F-II-3, M-II-1 and M-II-3. Additional faint T2/FLAIR-signal alterations are visible in the periventricular white matter in individuals A-II-2, M-II-1, M-II-3 and M-II-6 (thick arrows). There is hypoplasia of the corpus callosum (curved arrows) and small anterior commissure (thin arrows) in subjects F-II-3, L-II-1, M-II-1, M-II-3, M-II-6 and N-II-1. There is lack of myelination of the posterior limbs of the internal capsules in patient N-II-1 (dashed arrows), associated with foci of T2 hyperintensity at the level of the putamina. DaTscan of subjects A-II-2, B-II-3, C-II-3 and I-II-3 reveals severely reduced tracer uptake in the striatum bilaterally, with increased background activity. ^123^I-MIBG myocardial scintigraphy measured 15 minutes (early registration) and four hours (late registration) after intravenous ^123^I-MIBG application was normal in proband C-II-3 (30-35 years). ^123^I-MIBG = ^123^I-metaiodobenzylguanidine.

### Biallelic *PSMF1* variants cause a phenotypic spectrum from early-onset PD or parkinsonism to perinatal lethality with neurological manifestations

A total of 24 affected subjects from 17 unrelated families of European, Asian and African ancestry were identified (Fig.1a). Genotype was determined for all but two individuals (M-II-1, O-II-4; DNA not available), who were included due to striking clinical resemblance to their affected siblings carrying homozygous *PSMF1* variants in the context of parental consanguinity. One aborted fetus (Q-II-2) was included based on *PSMF1* variants identified on reanalysis of the ES data from prenatal genetic testing and phenotypic overlap with the proband of Pedigrees O-P, who died during infancy. Pregnancy losses in Pedigrees O-P were excluded due to insufficient pheno-genotypic information. Clinical features and neuroimages are highlighted in Fig.1b-c and described in Table 1.

**Table 1.** Phenotypic features and genotype of 24 subjects with *PSMF1*-related disorder belonging to 17 families. AC = anterior commissure; ASD = atrial septal defect; ASM = anti-seizure medication(s); CBZ = carbamazepine; C-HET = compound heterozygote; CC = corpus callosum; CGH = comparative genomic hybridization; CK = creatine kinase; CTG = cardiotocography; COPD = chronic obstructive pulmonary disease; DBS = deep brain stimulation; DD = developmental delay; DDH = developmental dysplasia of the hip; DEXA = bone density scan; ECG = electrocardiogram; EEG = electroencephalogram; F = female; GA = gestational age; GDD = global developmental delay; GER(D) = gastroesophageal reflux (disorder); GI = gastrointestinal; HOM = homozygote; ICD = impulse control disorder; LEV = levetiracetam; M = male; MDD = motor developmental delay; MIBG = ^123^I-metaiodobenzylguanidine scintigraphy; mo(s) = month(s); MRI = magnetic resonance imaging; MRS = magnetic resonance spectroscopy; N/A = not applicable or not available; PD = Parkinson’s disease; QF-PCR = quantitative fluorescence polymerase chain reaction; SLDD = speech and language developmental delay; SPECT = single-photon emission computed tomography; US = ultrasound; VEP = visual evoked potentials; VPA = valproic acid; wk(s) = week(s); yr(s) = year(s). *Clinical details not available (O-II-4 reported with a phenotype similar to his sister O-II-5).

The cohort comprised 12 males and 12 females. Parental consanguinity was reported in 10 of 17 pedigrees (58.8%). Probands’ parents were healthy. Based on the phenotypic features and severity, we distinguished three subgroups across subjects with *PSMF1*-related disorder and recognized clear genotype-phenotype correlation (Fig.1b).

In Pedigrees A-B-C-D-E-F-G-H-I, affected individuals manifested with early-onset PD closely resembling well-established recessive monogenic forms (e.g. *PRKN*, *PINK1*, *DJ-1*, *FBXO7*), or with parkinsonism as the core phenotypic feature. After unremarkable neurodevelopmental milestones, motor symptoms began between the first and fifth decade of life, particularly during late adolescence or the third decade. In this subgroup, parkinsonism was most often accompanied by mild pyramidal tract signs, moderate-severe dysphagia occurring early in the disease course and psychiatric manifestations. Brain MRI was normal or detected minor abnormalities, including mild-to-moderate cerebral or mild cerebellar atrophy (Fig.1c). Five probands (A-II-2, B-II-3, C-II-3, D-II-2, I-II-3) had markedly abnormal presynaptic dopaminergic imaging (Fig.1c). In two probands (A-II-2, C-II-3), normal tracer uptake on ¹²³I-metaiodobenzylguanidine (¹²³I-MIBG) myocardial scintigraphy indicates preserved cardiac sympathetic innervation (Fig.1c). Affected individuals had different degrees of response to dopaminergic treatments and tended to develop early motor and non-motor fluctuations and levodopa-induced dyskinesia. Two probands (D-II-2, E-II-4) underwent bilateral subthalamic nucleus deep brain stimulation associated with good response which was, nevertheless, blurred subsequently by fairly rapid disease deterioration. Three patients (B-II-4, D-II-2, E-II-4) died nearly two decades after symptom onset. All affected individuals in this subgroup harbored *PSMF1* missense variants, either in the homozygous state (Pedigrees B-C-D-E-F-G-H) or in compound heterozygosity with a predicted loss-of-function variant (Pedigrees A-I).

Affected individuals belonging to Pedigrees J-K-L-M presented with miscellaneous movement disorders, encompassing parkinsonism, moderate-to-severe pyramidal tract signs, ataxia or a combination thereof, most often in the context of neurodevelopmental delay and associated with mild-to-moderate intellectual disability, epilepsy and cognitive decline. They exhibited facial or skeletal dysmorphic features. In this subgroup, two probands (J-II-3, K-II-3) had sensorineural hearing loss. Brain MRI almost invariably revealed hypoplasia of the corpus callosum and/or various degrees of cerebral or vermian cerebellar atrophy (Fig.1c). Four patients (J-II-1, M-II-1, M-II-3, M-II-6; Fig.1a) died during adolescence because of rapidly progressive neurological deterioration starting early in the second decade of life. All affected individuals within this subgroup carried homozygous *PSMF1* splice variants.

The third subgroup includes affected subjects from Pedigrees N-O-P-Q, who presented with severe neurological manifestations since prenatal development, including arthrogryposis multiplex congenita, abnormal fetal movements (suspected for in utero seizures), epilepsy, profound developmental delay, limb spasticity, severe respiratory insufficiency and poor feeding. Death occurred during infancy in all affected individuals within this subgroup, except for proband N-II-1, who is currently dependent on full medical support for all vital functions. Agenesis or hypoplasia of the corpus callosum was detected in three probands (N-II-1, O-II-5, Q-II-2; Fig.1c). A history of pregnancy losses (O-II-3, P-II-1) preceding the index case was reported in two of these pedigrees (Fig.1a). All affected subjects in this subgroup had biallelic *PSMF1* loss-of-function variants, of which at least one predicted to cause complete loss of function.

### Validation and functional analysis of 14 missense and loss-of-function variants in *PSMF1*

We detected 14 different *PSMF1* mutant alleles, including five missense and nine loss-of-function variants (six splice, one nonsense, one start-loss and one frameshift; Fig.1a-2a). Variant pathogenicity predictions and computational analysis are detailed in Supplementary File 1. In all subjects with genomic DNA available, segregation analysis confirmed that biallelic *PSMF1* variants co-segregated with neurological disease phenotypes. Moreover, parents of affected individuals harbored one heterozygous *PSMF1* variant, and unaffected siblings of affected subjects were either heterozygotes for one *PSMF1* mutant allele or homozygotes for the *PSMF1* wild-type allele (Fig.1a). Among 22 affected subjects with genotype available, 18 were homozygotes and 4 were compound heterozygotes for *PSMF1* variants (Fig.1a).

**Figure 2.**
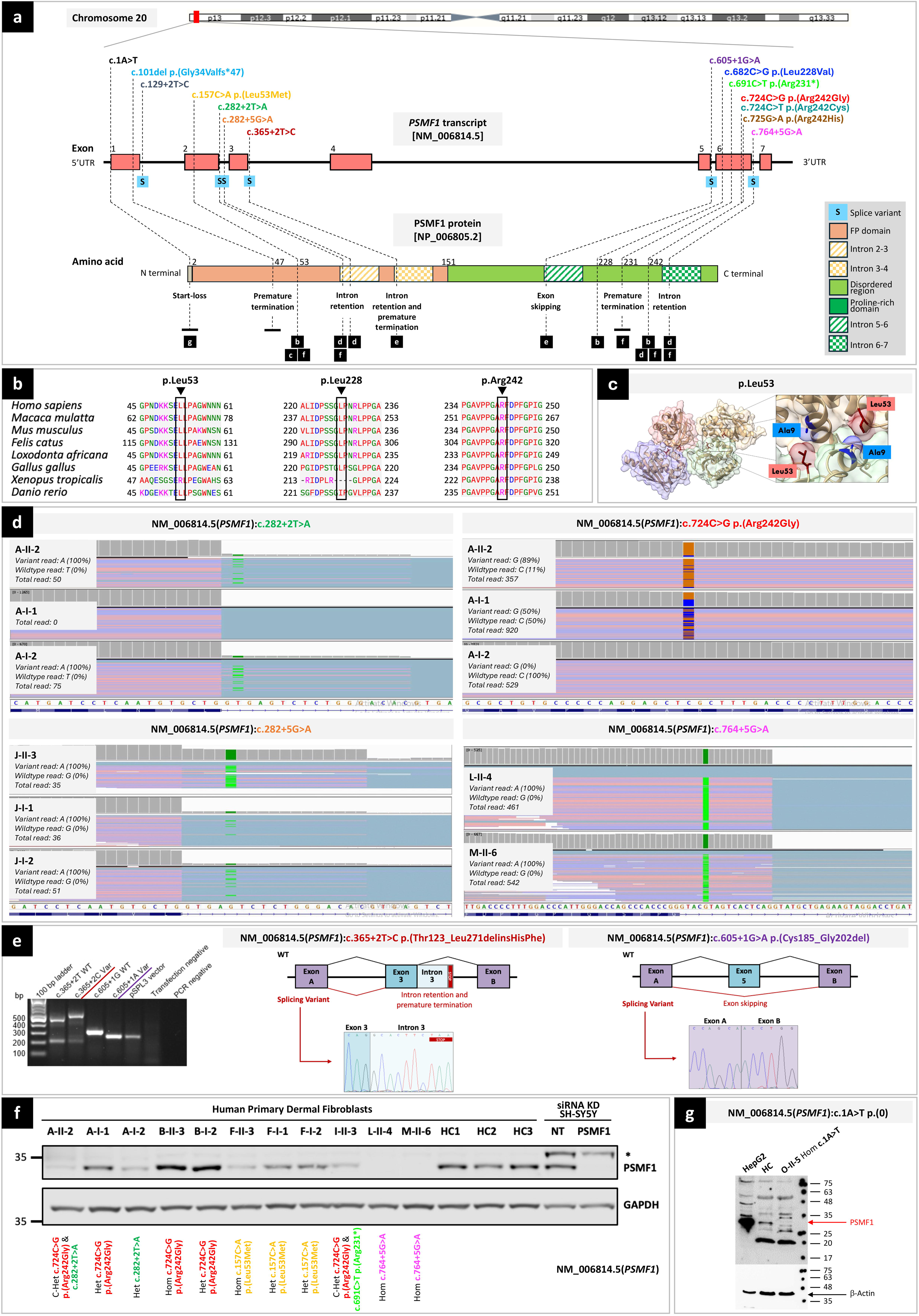
Location, computational analyses and functional characterization of *PSMF1* variants. **(a) Schematic of *PSMF1* and its protein product with location of 14 variants identified in this study and their effect.** *Upper part.* Chromosome 20 showing position of *PSMF1* on 20p13. *Middle part.* Schematic of *PSMF1* with variants identified in the human disease gene discovery study cohort. Introns are not to scale. Exon numbers are according to the canonical transcript (NM_006814.5). *Lower part.* Schematic of PSMF1 protein with amino acid changes linked to disease (reference sequence NP_006805.2). Black labels below individual *PSMF1* variants guide to corresponding panels of Figure 2 (b–g), where computational or functional characterization of these variants is presented. **(b) Interspecies alignment** showing strong evolutionary conservation of the amino acids affected by the *PSMF1* missense variants identified in this study across species, down to invertebrates. **(c) Localization of the p.Leu53 residue within the PSMF1–PSMF1 homotetramer crystal structure**, revealing a role for this residue in PSMF1 homodimerization. The residue p.Leu53 is involved in the *PSMF1* variant detected in Pedigrees F–G (Fig. 1a; Supplementary File 4). Image derived from PDB 4OUH using ChimeraX. **(d) RNA sequencing.** Integrative Genomics Viewer (IGV) screen captures showing results of RNA sequencing for Pedigree A (*upper panels*), Pedigree J (*left lower panel*) and probands L-II-4 and M-II-6 (*right lower panel*). For transcripts with *PSMF1* splice variants c.282+2T>A (A-I-2, A-II-2), c.282+5G>A (J-I-1, J-I-2, J-II-3), and c.764+5G>A (L-II-4, M-II-6), IGV enables visualization of (partial) intron retention. **(e) Minigene (splicing) assays.** *Left panel.* Agarose gel of the RT-PCR from the minigene cDNA. *Middle panel.* Splicing schematic of the NM_006814.5(*PSMF1*):c.365+2T>C variant showing intron retention with stop codon sequence marked. *Right panel.* Splicing assay of the NM_006814.5(*PSMF1*):c.605+1G>A variant shows exon skipping. **(f) PSMF1 immunoblots for human primary dermal fibroblasts from available probands, unaffected relatives and healthy controls** indicating loss of PSMF1 immunoreactivity in most affected individuals. Immunoblots for non-targeting siRNA and PSMF1 siRNA KD SH-SY5Ys shown alongside indicate specificity of the anti-PSMF1 antibody used. **(g) Western blot to functionally characterize the start-loss variant NM_006814.5(*PSMF1*):c.1A>T.** Protein lysates (30 μg) from HepG2 cells, and either control or proband (O-II-5) fibroblasts were analyzed by Western blotting using a polyclonal anti-PSMF1 antibody. HepG2 cells express PSMF1 at high levels. Control but not proband fibroblasts express PSMF1. β-actin expression was used as loading control. C-Het = compound heterozygote; Het = heterozygote; Hom = homozygote; siRNA KD = small interfering RNA knockdown.

All *PSMF1* variants identified in the families were absent or ultra-rare in the heterozygous state and not reported in the homozygous state in gnomAD,^36^ except for the missense variant c.725G>A p.Arg242His, which had low frequency and four homozygous entries in gnomAD v4.1.0 (Supplementary File 1). Interestingly, this mutant allele was detected in homozygosity in proband E-II-4, who exhibited the latest PD onset (45-50 years) among individuals in this study cohort. This may explain the presence of this *PSMF1* variant in gnomAD v4, a reference dataset primarily composed of individuals not selected for severe (pediatric) Mendelian disease, which could reflect its late-onset presentation and/or incomplete penetrance. Variant frequencies in other population and disease datasets, including 100,000 Genomes Project, UKB, *All of Us* Public Data Browser, and Queen Square Genomics are detailed in Supplementary File 1.

*PSMF1* missense variants identified in this study showed strong evolutionary conservation across species down to invertebrates (Fig.2b; Supplementary File 1A). All *PSMF1* non-splice variants were predicted with a damaging effect according to most *in silico* prediction tools (Supplementary File 1A). All *PSMF1* non-frameshift variants had a Combined Annotation Dependent Depletion v1.7 Phred score greater than or equal to 22.4 (CADD Phred range: 22.4-37), placing them among the top ∼0.6% most deleterious substitutions predicted in the human genome. Pathogenicity prediction tools such as PolyPhen2, SIFT4G, PROVEAN and MutationTaster predicted the functional impact of all *PSMF1* missense variants to be damaging/deleterious in most cases. *In silico* splice predictions of six *PSMF1* splice variants using six computational tools (SpliceSiteFinder-like, MaxEntScan, NNSPLICE, GeneSplicer, SpliceAI, and AbSplice) unanimously indicated reduction or complete abolishment of native splice donor sites according to most tools (Supplementary File 1B).

Five *PSMF1* variants were detected in more than one family (Pedigrees A-B-C-I: c.724C>G p.Arg242Gly; Pedigrees F-G: c.157C>A p.(Leu53Met); Pedigrees I-N: c.691C>T p.Arg231*; Pedigrees L-M: c.764+5G>A; Pedigrees O-Q: c.1A>T; Fig.1a). Specifically, the missense variant c.724C>G p.(Arg242Gly) recurred in four families either in the compound heterozygous (Pedigrees A-I) or homozygous (Pedigrees B-C) state. Furthermore, the missense variant c.724C>T p.(Arg242Cys) detected in Pedigree D involved the same nucleotide as the previous mutant allele. Finally, the missense variant c.725G>A p.(Arg242His) affected the same amino acid residue as the two previous mutant alleles. Intriguingly, although *PSMF1* variants were distributed across the gene (Fig.2a), three different *PSMF1* variants detected in a total of six families with PD/parkinsonism (Pedigrees A-B-C-D-E-I) involved the p.Arg242 residue, which could therefore represent a “hotspot codon”. Among splice variants, two were contiguous (c.282+2T>A in Pedigree A; c.282+5G>A in Pedigree J).

#### Autozygosity mapping

Given the apparent autosomal recessive inheritance mode of *PSMF1*-related disorder and the high prevalence of consanguineous pedigrees in our cohort, we performed autozygosity mapping to guide variant filtering and investigate potential alternative molecular diagnoses. Using AutoMap^37^ with default parameters (including a minimum ROH size threshold ≥ 1 Mb), we identified that the *PSMF1* locus fell within a region of homozygosity (ROH) in affected individuals carrying homozygous *PSMF1* variants who were born to consanguineous parents (Pedigrees B-D-F-G-H-K-L-M-N-O) and in the proband C-II-3, despite no reported parental consanguinity (Supplementary File 2). In contrast, no ROH encompassing *PSMF1* was detected by AutoMap in their unaffected family members with ES data available, nor in probands E-II-4 and J-II-1. However, in proband E-II-4, additional analysis for ROH ≥ 0.01 Mb revealed a 300-kb homozygous block including *PSMF1*. No ROH on chromosome 20 was found in proband J-II-3 or her affected sister J-II-1 (Supplementary File 2). Importantly, across all affected individuals harboring homozygous *PSMF1* variants, we did not identify any additional homozygous pathogenic variants or strong candidate variants within the ROHs that could account for the clinical phenotype observed.

#### Haplotype analysis

Haplotype analysis of the *PSMF1* missense variant c.724C>G p.(Arg242Gly), detected in homozygosity in three affected individuals from two families of different geographic origins (B-II-3, B-II-4; C-II-3), revealed distinct patterns of flanking variants. Similarly, analysis of the *PSMF1* splice variant c.764+5G>A, identified in homozygosity in the probands from two unrelated families from different regions (J-II-1; K-II-6), also showed differing haplotype backgrounds. In these analyses, the resolution of ES data is not enough to conclude whether this variant might have been inherited from a very ancient common ancestor (Supplementary File 3A-C). In contrast, haplotype analysis of the *PSMF1* missense variant c.157C>A p.(Leu53Met), found in homozygosity in probands from two unrelated families from the same country (F-II-3, G-II-3), revealed a shared homozygous block of 2.78 Mb. We hypothesized this shared genomic region to be an identity-by-descent haplotype and used the ExHap tool^38^ to calculate the age of the most recent common ancestor (MRCA). We deduced that the *PSMF1* variant c.157C>A p.(Leu53Met) is a founder variant that emerged between 720 to 556 years ago (Supplementary File 3B).

#### In silico modelling of PSMF1 missense variants

To assess the functional consequences of putative pathogenic *PSMF1* missense variants, modelling of the three-dimensional organization of the PSMF1-FBXO7 heterotetrameric complex was carried out using AlphaFold 3 (Supplementary File 4).^39^ While the p.Leu228 and p.Arg242 residues are located in PSMF1 regions with low-confidence structural prediction, the p.Leu53 residue sits directly on the interface between PSMF1 monomers within the complex, forming hydrogen bonds with p.Val6 and p.Ala9 (Fig.2c). Substitution of methionine for the leucine at codon 53 likely disrupts these interactions, weakening the quaternary structure of the predicted heterotetramer. Hence, the PSMF1 variant p.Leu53Met detected in Pedigrees F-G (Fig.1a) could perturb the function of the complex, acting as a loss-of-function variant. Further *in vitro* and cellular studies are required to test the impact of the p.Leu53Met variant, as well as to gain insight into the effect of coding variants in the C-terminal domain of PSMF1.

#### Functional characterization of PSMF1 variants

*RNA sequencing (RNA-seq).* In Pedigree A, fibroblast RNA-seq with transcript visualization through Integrative Genomics Viewer (IGV) and analysis through the Detection of RNA Outlier Pipeline (DROP) revealed partial intron retention in transcripts with the *PSMF1* splice variant c.282+2T>A, which the proband (A-II-2) and his mother (A-I-2) harbored in the heterozygous state (Fig.1a-2d). In proband A-II-2, who is a compound heterozygote for the *PSMF1* variants c.724C>G (p.Arg242Gly) and c.282+2T>A, sequencing read visualization around the missense variant revealed a wild-type to missense allele ratio of 11%:89%. This skewed ratio suggests nonsense-mediated mRNA decay (NMD) of the allele in *trans* carrying the splice variant (Fig.1a-2d). PSMF1 was not detected as aberrant expression (AE) or aberrant splicing (AS) outlier in these individuals. In Pedigree J, fibroblast RNA-seq of the proband (J-II-3) and whole-blood RNA-seq of his parents (J-I-1, J-I-2) with transcript visualization by IGV showed intron retention in transcripts with the *PSMF1* splice variant c.282+5G>A, which was detected in the homozygous state in the proband and in the heterozygous state in his parents (Fig.1a-2d). Here, DROP pipeline detected PSMF1 aberrant splicing of most transcript (OUTRIDER, PSMF1 as AE outlier, with fold change 0.13, z-score -9.84, FDR 6.29×10^-14^; FRASER2, PSMF1 as AS outlier, with |Δψ| 0.73, FDR 2.85×10^-12^). Finally, in probands L-II-4 and M-II-6, fibroblasts RNA-seq revealed that the *PSMF1* splice variant c.764+5G>A caused intron retention (Fig.2d), and PSMF1 was detected as significant splicing outlier in the two probands (FRASER2, PSMF1 as AS outlier, with |Δψ| 0.88, FDR 4.06×10^-5^ in L-II-4 and with |Δψ| 0.89, FDR 9.05×10^-6^ in M-II-6; OUTRIDER, PSMF1 not detected as AE outlier).

*Splicing (minigene) assay.* Minigene assay revealed that the *PSMF1* splice variant c.365+2T>C (Pedigree Q) led to intron retention (r.366_367inscacuucuaa) and premature termination (p.Thr123_Leu271delinsHisPhe; Fig.2e). Furthermore, the *PSMF1* splice variant c.605+1G>A (Pedigree K) was proven to result in skipping of exon 5 (r.553_606del) and an in-frame deletion (p.Cys185_Gly202del; Fig.2e).

To assess the potential functional impact of *PSMF1* splice variants in the substantia nigra, we analyzed PSMF1 transcript expression using data from the GTEx database. This revealed that the transcript with the highest expression in the substantia nigra (ENST00000333082.7) shares an identical coding sequence −including the same ATG start codon and canonical splice sites− with the MANE Select transcript (NM_006814.5; ENST00000335877.11), differing only in untranslated regions. Consequently, disruptions of canonical splice sites affecting the MANE Select transcript are expected to similarly affect the substantia nigra isoform ENST00000333082.7. In fibroblasts, the most abundant PSMF1 transcript (ENST00000479715.5) differs from the fibroblast-expressed isoform ENST00000333082.7 −which preserves the same coding sequence and canonical splice sites as the MANE Select transcript− only in a region corresponding to exon 2 and its flanking intronic sequences. This region includes the c.282+2T>A and c.282+5G>A variants, for which we demonstrated aberrant splicing in transcripts from patient-derived fibroblasts (Fig.2d). These findings demonstrate that the transcript isoform harboring the clinically relevant splice site is endogenously expressed in both fibroblasts and substantia nigra, supporting the pathogenic relevance of *PSMF1* splice-disrupting variants in a disease-relevant tissue.

##### Western blotting

Western blotting of protein lysate obtained from fibroblasts of probands L-II-4 and M-II-6 harboring the intron retaining PSMF1 variant c.764+5G>A in the homozygous state showed complete loss of anti-PSMF1 antibody immunoreactivity (Fig.2f). A severe reduction in anti-PSMF1 immunoreactivity was also observed for probands A-II-2, F-II-3 and I-II-3 compared to their unaffected parents (A-I-1, A-I-2, F-I-1, F-I-2) and/or healthy controls (Fig.2f). On the other hand, Western blotting of protein lysate from fibroblasts of proband B-II-3, who was homozygote for the PSMF1 missense variant c.724C>G p.(Arg242Gly) suggests that this variant does not impact protein stability and instead is most likely affecting protein function (Fig.2f). Finally, Western blotting of protein lysate obtained from fibroblasts of proband O-II-5 (Fig.1a), who was homozygote for the *PSMF1* start-loss variant c.1A>T, revealed that PSMF1 antibody immunoreactivity was abolished (Fig.2g).

### Proteomic analysis of human fibroblasts revealed a descending PSMF1 abundance profile from missense to loss-of function mutant alleles

Differential expression analysis of the proteomes of *PSMF1* proband- and heterozygous carrier-derived fibroblasts was obtained by nano-liquid chromatography combined with mass spectrometry. This revealed statistically significant reduction in PSMF1 in probands carrying homozygous *PSMF1* splice variants (J-II-3, L-II-4, M-II-6) compared to controls (Fig.3a). Statistically significant reduction in PSMF1 expression was also observed between probands harboring homozygous *PSMF1* splice variants and those with homozygous *PSMF1* missense mutations (B-II-3, F-II-3; Fig.3b). On the contrary, PSMF1 expression was not downregulated in probands carrying homozygous *PSMF1* missense variants compared to controls or carriers of a single heterozygous *PSMF1* missense variant (A-I-1, A-I-2, B-II-1, F-I-1, F-I-2; Fig.3c-d). Interestingly, other proteasome-associated proteins were significantly upregulated in probands with homozygous *PSMF1* missense variants compared to homozygotes for *PSMF1* splice variants, heterozygotes for a *PSMF1* missense variants and controls (Fig.3b-c-d), although the majority of these do not pass the log2 fold-change threshold of 1. Proteasome subunits B and proteasome activator complex subunits E were statistically significant in probands- and missense-homozygote mutations compared to heterozygous carriers and controls. An upward trend was evident in PSMF1 levels from probands to carriers of single heterozygous *PSMF1* splice or missense variants, approaching the highest PSMF1 levels recorded in controls (Fig.3e). FBXO7 was significantly downregulated in patients with homozygote splice variants compared to controls, albeit with a log2 fold-change in protein abundance <1 (Fig.3f). Further experiments including undertaking organelle enrichment studies are needed to comprehensively elucidate the mechanistic impacts of *PSMF1* missense or loss-of-function variants on protein abundance regarding proteasomal, mitochondrial and other relevant pathways (Supplementary File 5).

**Figure 3.**
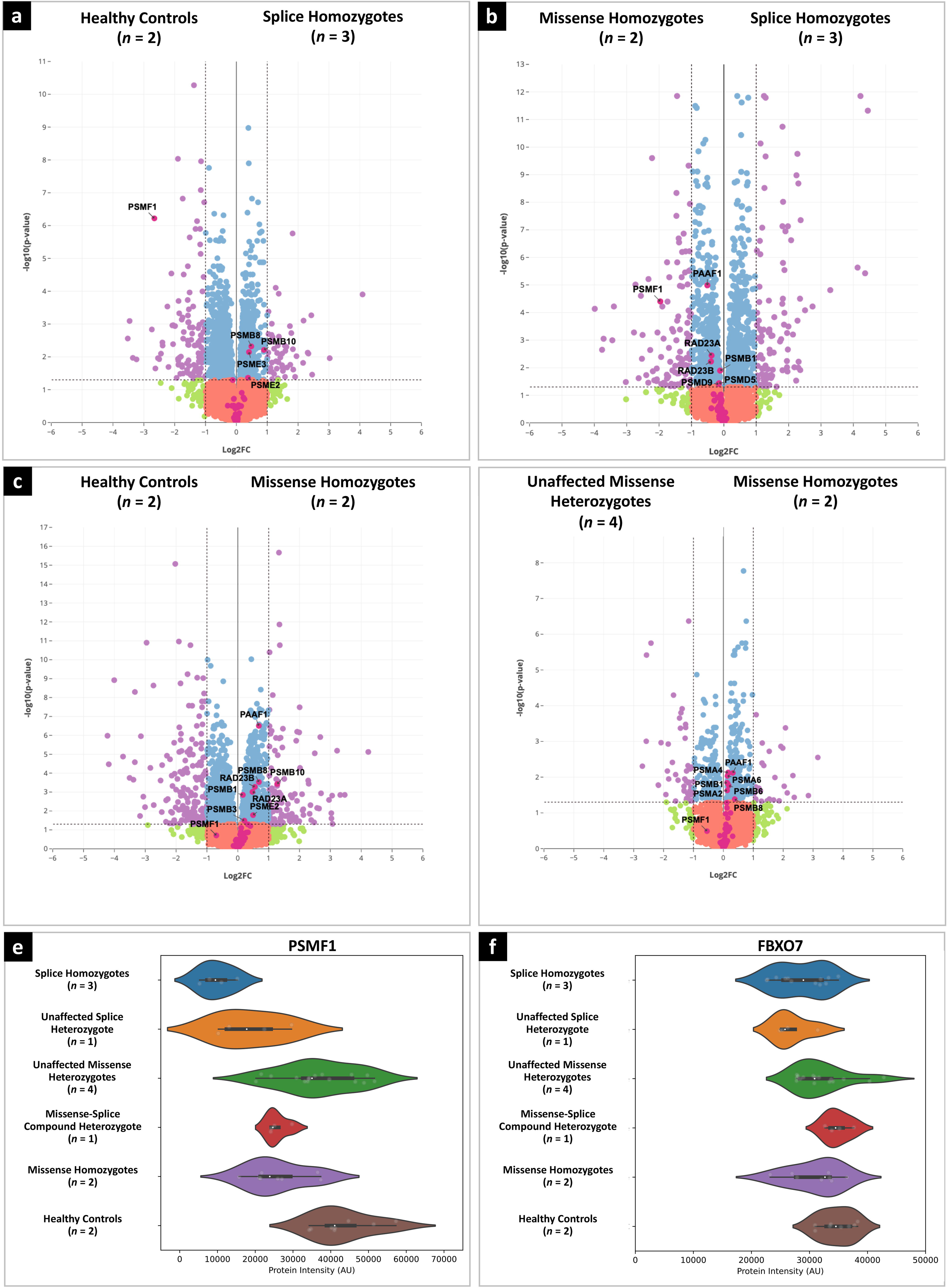
Quantitative proteomic analysis of *PSMF1* patient- and carrier-derived dermal fibroblasts. Volcano plots showing: **(a)** statistically significant downregulation of PSMF1 in affected individuals with homozygous *PSMF1* splice variants (*n* = 3; J-II-3, L-II-4, M-II-6) compared to healthy controls (HC); **(b)** Statistically significant reduction in PSMF1 expression in affected individuals with homozygous *PSMF1* splice variants (*n* = 3; J-II-3, L-II-4, M-II-6) compared to probands with homozygous *PSMF1* missense variants (*n* = 2; B-II-3, F-II-3); **(c)** PSMF1 expression was not downregulated in probands with homozygous *PSMF1* missense variants (*n* = 2; B-II-3, F-II-3) compared to HC or **(d)** unaffected individuals having a single heterozygous *PSMF1* missense variant (*n* = 4; A-I-1, B-II-1, F-I-1, F-I-2). Different colored dots within each quadrant of the volcano plot denote their respective p-value cut-off (0.05 > p OR p ≤ 0.05; y-axis) and log₂ fold-change (FC > 1 OR FC ≤ 1; x-axis). Additional pink-colored dots indicate proteins associated with the proteasomal pathway (as annotated within the Curtain web tool: https://curtain.proteo.info/#/), with differentially abundant proteins that passed the statistical significance threshold (p < 0.05) being further annotated by text within each volcano plot. Violin plots showing: **(e)** PSMF1 protein expression across different groups, including probands with homozygous *PSMF1* splice variants (*n* = 3; J-II-3, L-II-4, M-II-6), unaffected individuals with one heterozygous *PSMF1* splice variant (*n* = 1; A-I-2) or one heterozygous *PSMF1* missense variant (*n* = 4; A-I-1, B-II-1, F-I-1, F-I-2), proband with one *PSMF1* splice and one *PSMF1* missense variant in compound heterozygosity (*n* = 1; A-II-2), probands with homozygous *PSMF1* missense variants (*n* = 2; B-II-3, F-II-3), and healthy controls; **(f)** FBXO7 protein expression across the same groups. Patients with homozygous *PSMF1* splice variants (*n* = 3; J-II-3, L-II-4, M-II-6) exhibit a statistically significant reduction (p < 0.05) in protein expression vs HC following differential expression analysis, albeit with a log_2_ fold-change < 1. HC = healthy controls (*n* = 2; M:F = 0:1; age at skin biopsy = 19.5 ± 0.5 years).

### Heterozygous rare and low-frequency *PSMF1* variants are not associated with increased risk of sporadic Parkinson’s disease in the UKB and AMP-PD repositories

We looked for rare and low-frequency (minor allele frequency, MAF <0.005) *PSMF1* variants in patients with sporadic (non-familial) PD and controls in the UKB and AMP-PD repositories. Gene-burden analysis revealed that single heterozygous *PSMF1* variants do not contribute to the risk of developing sporadic PD (Supplementary File 6).

### Real-time quaking-induced conversion assay did not identify misfolded α-synuclein seeding activity in three *PSMF1* probands

We performed α-synuclein (α-syn) Real-Time Quaking-Induced conversion (RT-QuIC) assay in the CSF of proband A-II-2 and in the skin of probands A-II-2, B-II-3 and I-II-3 according to an established protocol.^40^ All α-syn RT-QuIC runs with both matrices were negative (Supplementary File 7), thus ruling out significant synucleinopathy as the substrate of nigrostriatal degeneration in these affected individuals.

### *PSMF1* variants can affect proteasomal abundance and assembly in human fibroblasts

Extensive evidence indicates that PSMF1 functions as a proteasome regulator and promotes proteolysis in multiple species, including *Drosophila*, mice, yeast, and plants.^18,22-24^ Furthermore, PSMF1 facilitates the recruitment of proteasomes at synaptic sites, and is essential for synapse maintenance and neuronal survival in *Drosophila* and mice.^20,22^ We preliminarily investigate the effects of *PSMF1* variants on proteasome functions in lysates from selected *PSMF1* proband-derived fibroblasts. Native gel assays revealed a reduction in the amount of both 20S and 26S proteasome particles in samples from *PSMF1* probands A-II-2 and M-II-6 compared to healthy controls, as well as a reduction in the ratio of assembled 26S particles (Supplementary File 8A). These observations are consistent with the previously reported role of PSMF1/PI31 in promoting 26S assembly from 19S and 20S sub-particles.^19^ In addition, we detected reduced alpha-7 subunit immunoreactivity in F-II-3 proband-derived fibroblast samples, as assessed by both native gel and Western blot analysis. This is in keeping with reduced amounts of proteasomes in this *PSMF1* proband (Supplementary File 8B).

### *PSMF1* variants cause mitochondrial dysfunction and alteration of mitochondrial dynamics and mitophagy in human fibroblasts

We explore the effects of *PSMF1* variants on mitochondrial functions and mitophagy by live imaging of skin fibroblast of *PSMF1* probands and carriers.

#### Mitochondrial membrane potential

Most processes in mitochondria depend on the mitochondrial membrane potential (MMP), whose detection in live cells is an important indicator of healthy mitochondrial functioning. Using tetramethylrhodamine methyl ester (TMRM) as fluorescent probe for MMP, we found that TMRM fluorescence in fibroblasts from *PSMF1* probands was similar or higher and in unaffected *PSMF1* carriers even higher than in controls (Fig.4a-b), which suggests mitochondrial hyperpolarization. Since MMP is mainly maintained by the function of the mitochondrial electron transport chain (mETC), any perturbation of mitochondrial respiration switches mETC to alternative mechanisms to maintain MMP in the physiological range, including reversal of the activity of the F0-F1-ATPase (complex V). Fibroblasts from controls or *PSMF1* carriers showed no effect or small increase in TMRM fluorescence after application of the mitochondrial complex V inhibitor oligomycin (2μg/ml; Fig.4c, upper quadrants), indicating that that enzyme is working as ATP synthase. On the contrary, addition of oligomycin to *PSMF1* proband-derived fibroblasts induced >50% depolarization (Fig.4c, lower quadrants), which suggests that complex V is working in ATPase mode due to pathological mETC functioning in these cells. Subsequent application of the inhibitor of mitochondrial complex I rotenone (5μM) induced decrease in TMRM fluorescence in control (by ∼50%) or *PSMF1* carriers’ fibroblasts (by ∼60%; Fig.4c, upper quadrants), whereas in *PSMF1* probands’ cells it induced complete mitochondrial depolarization, seen by the lack of effect of the mitochondrial uncoupler carbonyl cyanide p-trifluoromethoxyphenylhydrazone (FCCP; 1μM) on TMRM fluorescence after oligomycin and rotenone in these fibroblasts (Fig.4c, lower quadrants). In control or *PSMF1* carriers’ fibroblasts, subsequent application of FCCP induced further drop of the TMRM signal (by 30-40%) to a complete depolarization (Fig.4c, upper quadrants). Overall, control fibroblasts maintain MMP solely by the function of the mETC (including complexes I and II), while MMP maintenance (∼50%) in probands’ fibroblasts depends on ATP consumption by the F0-F1-ATPase and the activity of complex I. Relevantly, we previously observed similar changes in MMP maintenance in other cells with PD pathology, including *PINK1*-deficient cells, neurons with *SNCA* triplication and neurons with mitochondrial DNA variants.^41-43^

**Figure 4.**
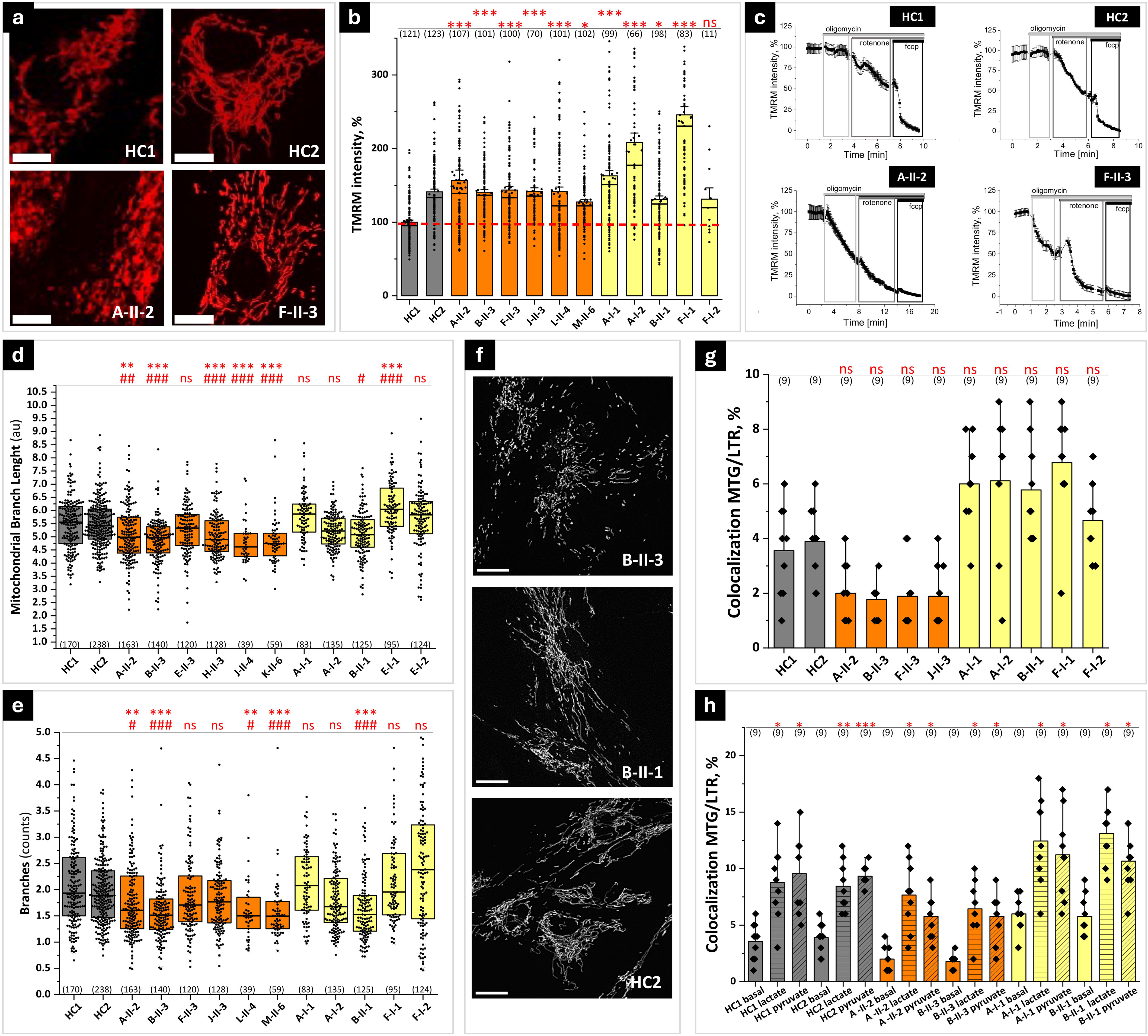
PSMF1 deficiency is associated with defects in mitochondrial bioenergetics and dynamics. **(a-c) Mitochondrial membrane potential status and maintenance: (a)** Representative images of tetramethylrhodamine methyl ester (TMRM)-loaded mitochondria in fibroblasts from healthy controls (HC1, HC2) and *PSMF1* probands (A-II-2, F-II-3; Fig.1a). Scale bar = 20 μm. **(b)** Quantification bar graphs depict the mean mitochondrial membrane potential for HC’ (gray bars; see also red dash line), probands’ (orange bars) and unaffected carriers’ (yellow boxes) mitochondria. Non-parametric Kruskal-Wallis ANOVA with post-hoc Dunn’s test for each group. Data are presented as mean ± standard error of the mean (SEM). *p < 0.05, **p < 0.01, ***p < 0.001, ns = not statistically significant. **(c)** Inhibitor analysis of the mitochondrial membrane potential maintenance and electron transport chain function in HC (*upper quadrants*) compared to probands’ (*lower quadrants*) mitochondria. **(d-f) Mitochondrial dynamics: (d)** Quantification box charts depicting average length of all mitochondrial rod and branches and **(e)** average number of branches analyzed in individual fibroblasts from HC’ (gray boxes), probands’ (orange boxes) and unaffected carriers’ (yellow boxes) fibroblasts. **(f)** Representative images of mitochondrial shape and dynamics in one unaffected carrier (B-II-1), one proband (B-II-3) and one HC (HC2). In **(d)** and **(e)** each dot represents a cell and total number of cells (*n*; shown in brackets) analyzed in 3-9 independent experiments (*N*). Non-parametric Kruskal-Wallis ANOVA with post-hoc Dunn’s test for each group. *p < 0.05, **p < 0.01, ***p < 0.001 compared to control HC1; # p < 0.05, ## p < 0.01, ### p < 0.001 compared to control HC2. Scale bar = 20 μm. **(g-h) Mitophagy: (g)** Basal mitophagy rate assessed as co-localization of LysoTracker Red DND-99 with MitoTracker Green in HC (gray bars), probands’ (orange bars) and unaffected carriers’ (yellow bars) fibroblasts. **(h)** Mitophagy rate after induction with either lactate or pyruvate. In (g) and (h) each dot represents the number of cover slips (*n*; shown in brackets) from 3-6 independent culturing conditions (*N*). Non-parametric Kruskal-Wallis ANOVA with post-hoc Dunn’s test for each group. Data are presented as mean ± standard error of the mean (SEM). *p < 0.05, **p < 0.01, ***p < 0.001, ns = not statistically significant.

#### Mitochondrial dynamics

Mitochondrial dynamics is essential for proper mitochondrial metabolism and function. Mitochondria in *PSMF1* proband- and control-derived fibroblasts differ not only by the TMRM signal intensity but also by their shape and size (Fig.4d-e-f). Further analysis of the mitochondrial shape showed that mitochondria in the probands’ fibroblasts appeared more fragmented and have shorter mitochondrial branch length (Fig.4d) and lower mean number of branches per network (Fig.4e) than control fibroblasts. Interestingly, fibroblasts of *PSMF1* carriers had even longer mitochondrial branch length and mean number of branches per network than in control and probands’ fibroblasts (Fig.4d-e-f). Therefore, *PSMF1* variants cause changes in mitochondrial dynamics, which could be secondary to an alteration of fission/fusion mechanisms or mitophagy.

#### Mitophagy

Damaged or depolarized mitochondria are engulfed by lysosomes during mitophagy. To detect this translocation by live imaging of participant-derived fibroblasts, we analyzed the co-localization of mitochondria loaded with the green-fluorescent dye MitoTracker® Green FM with lysosomes loaded with the red-fluorescent dye LysoTracker Red DND-99.^44^ Although basal mitophagy levels in fibroblasts are relatively low and the difference between PD and control cells is difficult to detect,^45^ we found that the percentage of co-localization in *PSMF1* probands’ fibroblasts was lower than in *PSMF1* carriers (Fig.4g). Mitophagy activation by 30mM lactate or 30mM pyruvate^45^ increased the percentage of mitochondria-lysosome co-localization in all fibroblasts, but to a higher extent in control and *PSMF1* carriers’ fibroblasts than in probands’ fibroblasts (Fig.4h).

### *Psmf1* knockdown in *Drosophila* causes age-dependent locomotor dysfunction, dopaminergic neurodegeneration, and mitochondrial membrane depolarization

We utilized the UAS/Gal4 system^46^ to induce pan-neuronal (*nSyb-Gal4*) RNA interference (RNAi)-mediated knockdown of *psmf1* in *Drosophila*. We similarly generated positive (knockdown of the established PD-related gene *prkn*) and negative (knockdown of *nsmase*, previously used as a negative control)^47^ controls.

#### Behavioral analysis (negative geotaxis climbing assay)

Motor performance of *psmf1*-knockdown flies was assessed using the classic negative geotaxis climbing assay (Fig.5a-b). We observed no difference in climbing performance between the three groups at 10 or 20 days after eclosure, but at 35 days both *prkn*- and *psmf1*-knockdown flies exhibited impaired motor performance in the form of a reduced proportion of flies climbing past a 5-cm threshold relative to a previously established *nsmase* negative control (Fig.5b). These observations are consistent with loss of psmf1 function causing motor dysfunction in aged flies to a degree comparable to established *Drosophila* PD models.^48,49^

**Figure 5.**
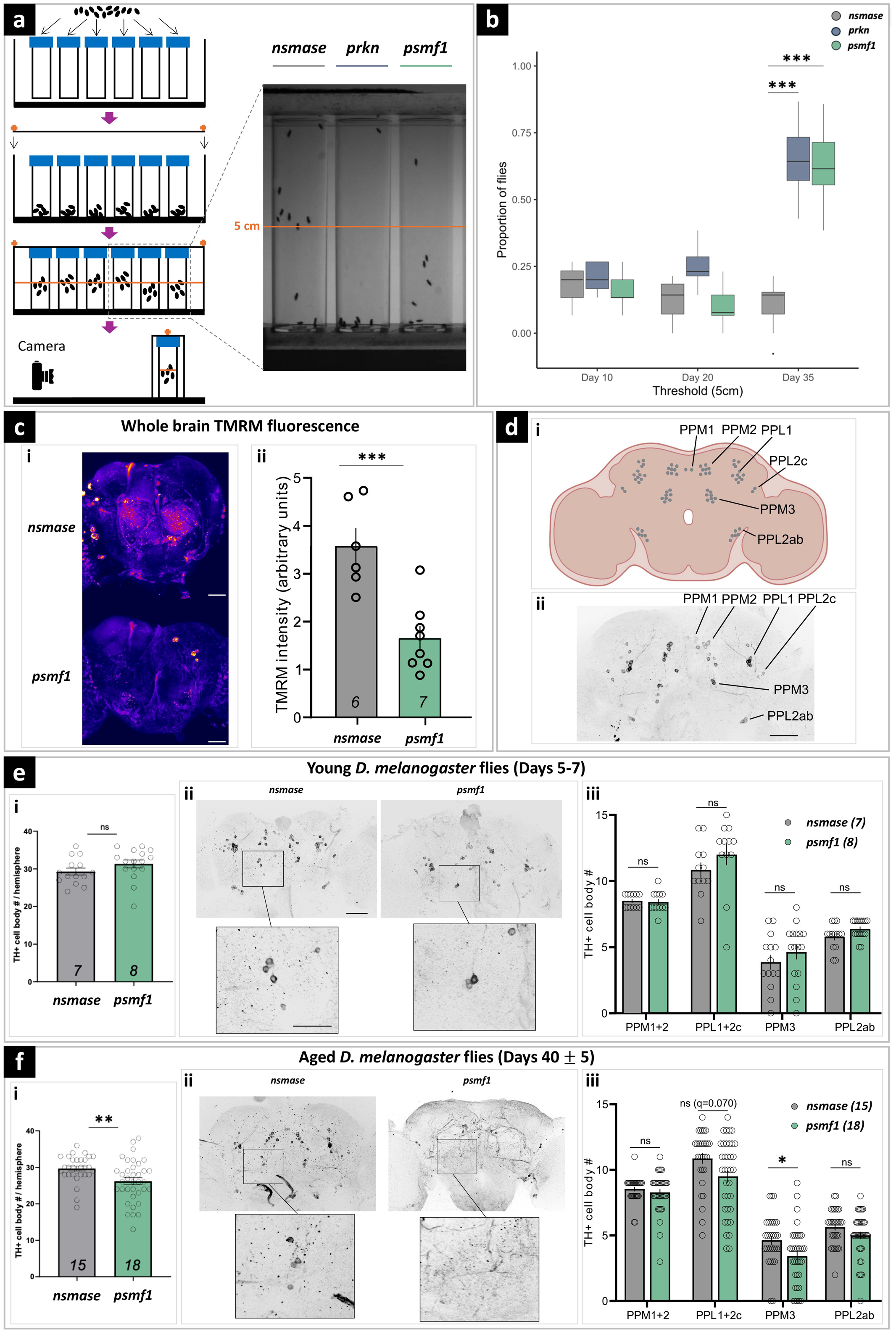
Effects of pan-neuronal *psmf1* knockdown in *Drosophila*. **(a)** Setup for comparative climbing test of three different *Drosophila* RNA interference (RNAi) lines (*nsmase*, *prkn*, *psmf1*) with *prkn* used as positive control (significant motor impairment) and *nsmase* as a negative control (no motor impairment). *H. sapiens* and *Drosophila* orthologues: *PRKN* (*H. sapiens*)/*prkn* (*Drosophila*); *PSMF1* (*H. sapiens*)/*psmf1* (*Drosophila*); *SMPD2* (*H. sapiens*)/*nsmase* (*Drosophila*). **(b)** Climbing test performed at Days 10, 20 and 35 after eclosure represented as percentage of flies that did not reach the 5-cm tube threshold. Significant motor defect was observed at Day 35 after eclosure for the *psmf1* and *prkn* lines compared to the negative control *nsmase*. Analyses are χ² (chi-squared) tests. ***p < 0.0005. **(c)** Whole-brain mitochondrial membrane potential. **(c-i)** Representative images of TMRM fluorescence in whole brain preparations from 35 days post-eclosure *psmf1*- and control *nsmase*-knockdown flies (scale bar = 50 μM). **(c-ii)** Quantification of mean TMRM fluorescence within region of interests (ROIs) encompassing the whole central brain. *N* values are number of brains. Analysis is Mann-Whitney U-test. ***p < 0.0005. **(d-i)** Schematic showing the location of dopaminergic neuron clusters in the adult *Drosophila* brain: PPM1-3, protocerebral posterior medial clusters 1-3; PPL1-2c, protocerebral posterior lateral clusters 1-2c. **(d-ii)** Representative image of anti-TH immunostaining in a control (*nsmase*-knockdown, 5 days after eclosure) brain, with the same neuronal clusters indicated (scale bar = 50 μM). Total number of TH+ cell bodies in young (5–7 days after eclosure) **(e-i)** and aged (40 ± 5 days after eclosure) **(f-i)** brains. *N* values are number of brains. Analysis is unpaired t-test. **p < 0.005. **(e-ii, f-ii)** Representative images of anti-TH immunostaining in the indicated genotype (scale bar = 50 μM), with enlarged sections showing PPM3 clusters (scale bar = 20 μM). **(e-iii, f-iii)** Number of TH+ cell bodies in indicated clusters per hemisphere (i.e. two data points per brain). Indicated *n* values are number of brains. Analyses are Mann-Whitney tests within clusters, with False Discovery Rate correction for multiple comparisons. *q < 0.05.

#### Mitochondrial membrane potential

Since we observed mitochondrial membrane depolarization in fibroblasts derived from *PSMF1* probands in response to complex I and V inhibitors, we used the *Drosophila* model to explore whether this occurred under normal physiological conditions in the context of an organismal model. At 35 days after eclosure, the stage at which motor defects became apparent, we observed a brain-wide decrease in TMRM fluorescence in *psmf1*-knockdown flies relative to *nsmase* negative controls, again comparable to reports in established *Drosophila* PD models (Fig.5c).^50^ These data indicate a highly conserved function of psmf1 between *D. melanogaster* and *H. sapiens*, and are consistent with loss of psmf1 function leading to mitochondrial dysfunction and depolarization.

#### Brain immunohistochemistry

Loss of prkn function in *Drosophila* causes progressive degeneration of dopaminergic neurons.^51^ We therefore tested whether this hallmark of PD was apparent in *psmf1*-knockdown flies. The location of dopaminergic neuron clusters in the *Drosophila* brain is well characterized, and they are readily visualized via immunostaining against tyrosine hydroxylase (TH) (Fig.5d). In young (5-7 days post-eclosure) flies there was no difference in the total number of TH^+^ cell bodies between *psmf1*- (mean ± SEM: 31.31 ± 1.06) and *nsmase*-knockdown (29.29 ± 0.946) brains (p = 0.170, unpaired t-test) (Fig.5e-i), or in the number of neurons within individual clusters (Fig.5e-ii and 5e-iii). However, we counted significantly fewer TH^+^ cell bodies in aged (40 ± 5 days post-eclosure) *psmf1*-knockdown (26.19 ± 0.965) brains relative to *nsmase* negative controls (29.67 ± 0.6701) (p = 0.0034, Mann-Whitney test) (Fig.5f-i), with significantly reduced numbers in the PPM3 cluster and a trend towards reduction in the PPL1+2c clusters (q = 0.07) (Fig.5e-ii and 5e-iii). This is consistent with loss of psmf1 function causing PD-like progressive degeneration of dopaminergic neurons in *Drosophila*.

### Conditional inactivation of *Psmf1* in mice leads to motor dysfunction and diffuse gliosis

*Mus musculus* Psmf1 protein is 83% identical and 91% similar to human PSMF1. Members of our team previously reported that *Psmf1* inactivation in mouse neurons causes age-progressive neurological phenotypes which culminate in neuronal cell death.^22^ To better examine the consequences of impaired Psmf1 function in the mouse, we bred mice with a tamoxifen-inducible Cre recombinase system with a Cre-ERT2 fusion gene under the control of the human ubiquitin C promoter (UBC-Cre-ERT2)^52^ with mice containing a loxP-flanked (floxed) third exon of *Psmf1* (*Psmf1*^fl^),^22^ thus obtaining conditional knockout (CKO) of *Psmf1*. This strain enables conditional inactivation of *Psmf1* upon tamoxifen injection. To assess recombination efficiency, these mice also carried a transgenic Cre reporter, Ai14,^53^ which has a floxed STOP cassette preventing transcription of a CAG promoter-driven red fluorescent protein variant (tdTomato) inserted into the Gt(ROSA)26Sor locus. Upon successful Cre-mediated recombination, Ai14 mouse tissues express tdTomato red fluorescence. Depending on the amount of tamoxifen injected, we obtained recombination efficiency ranging from 30% to over 90% (Fig.6a; Supplementary File 9).

**Figure 6.**
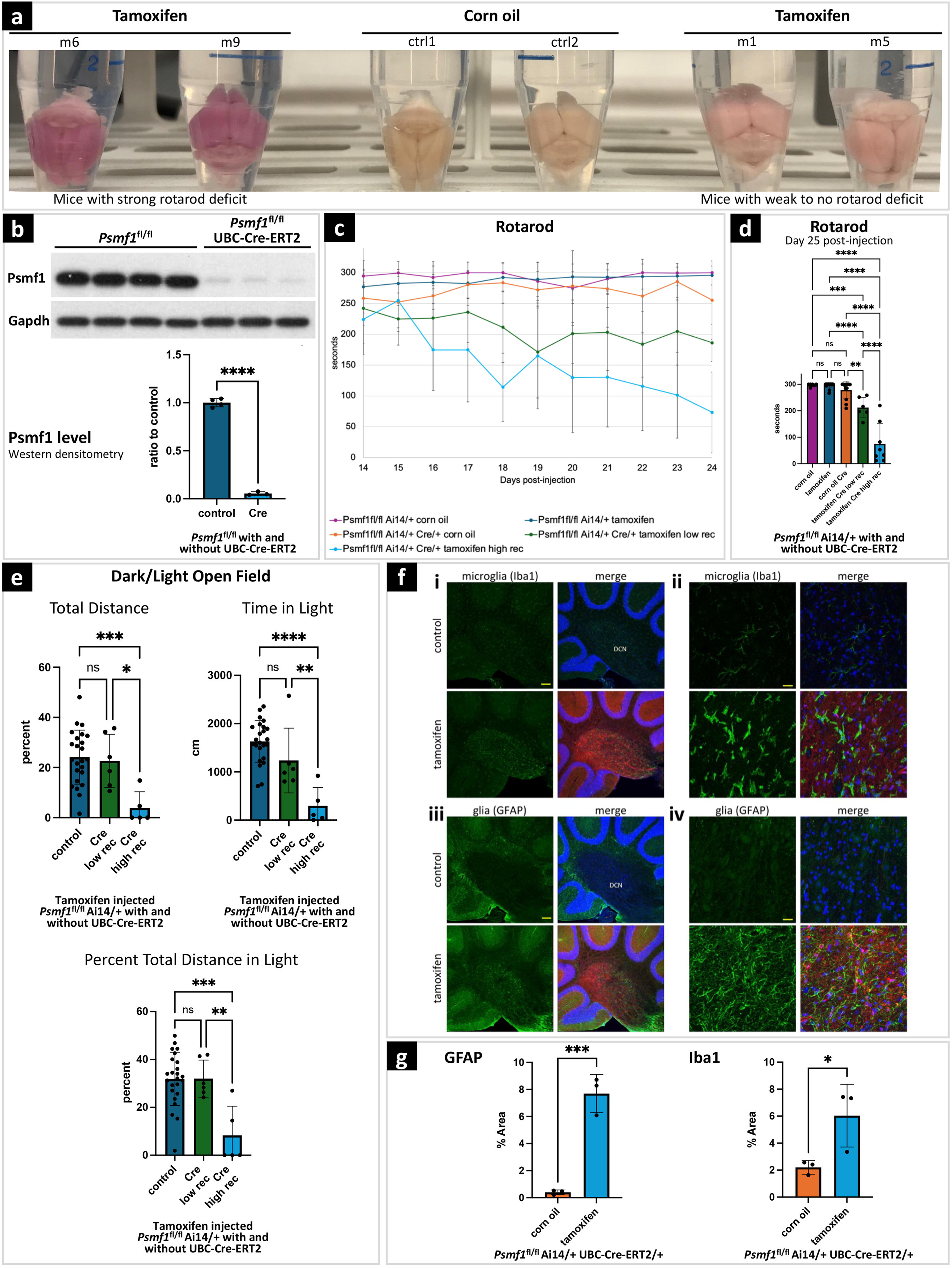
Behavioral tests and neuropathology in mice with conditional *Psmf1* inactivation. **(a)** *Psmf1*^fl/fl^ UBC-Cre-ERT2/+ Ai14/+ brains post perfusion (28 days after injection of tamoxifen or corn oil as a control). Tamoxifen-induced recombination induced tdTomato (red) expression and marked cells in which *Psmf1* was inactivated (Supplementary File 9). Successful tamoxifen-induced recombination was macroscopically assessed based on the color of dissected mouse brains, which appeared dark pink in mice with severe motor deficit in the rotarod assay (m6, m9) and light pink in mice with mild to no motor deficit in the rotarod assay (m1, m5) compared to controls (ctrl1, ctrl2), which reflected high (∼90%) or low (30–50%) recombination rates, respectively. *Psmf1*^fl/fl^ Ai14 UBC-Cre-ERT2 mice were injected with either corn oil (control) or tamoxifen to induce Cre recombination of floxed *Psmf1* exon 3 for loss of Psmf1 and a floxed stop cassette in Ai14 for expression of tdTomato to evaluate recombination efficiency. Mice were sacrificed 28 days post injection. **(b)** Quantification of Psmf1 protein levels in mice. Western blot analysis for Psmf1 and Gapdh of whole brain extracts from mice 20 days post tamoxifen injection for high recombination rate. Less than 6% Psmf1 remains in tamoxifen-injected Cre mice. Quantification of Western blots by densitometry, *n* = 4 *Psmf1*^fl/fl^, *n* = 3 *Psmf1*^fl/fl^ UBC-Cre-ERT2. T-test, ****p < 0.0001. **(c–d) Rotarod assay.** In the rotarod assay, mice with high recombination rates (∼90%) developed severe motor deficits (light blue line in **(c)** and light blue bar in **(d)**, whereas the motor performance of mice with low (30–50%) recombination rates was mildly or not impaired (green line in **(c)** and green bar in **(d)**). Panel **(c)** shows single rotarod trials for individual mice on each day. X-axis indicates days of testing after the initial four-day training period. Day 1 corresponds to Day 14 after tamoxifen injection. In panel **(d)**, each bar represents the average of three trials for a mouse on Day 25 post tamoxifen injection (i.e., 12 days after the initial four-day training period). Error bars indicate standard deviations. One-way ANOVA with Tukey post-hoc test. *n* = 6 *Psmf1*^fl/fl^ Ai14/+ corn oil, *n* = *Psmf1*^fl/fl^ Ai14/+ tamoxifen, *n* = 12 *Psmf1*^fl/fl^ Ai14/+ UBC-Cre-ERT2/+ corn oil, *n* = 6 *Psmf1*^fl/fl^ Ai14/+ UBC-Cre-ERT2/+ tamoxifen low recombination rate, and *n* = 8 *Psmf1*^fl/fl^ Ai14/+ UBC-Cre-ERT2/+ tamoxifen high recombination rate. **p < 0.01, ***p < 0.001, **\*\***p < 0.0001. **(e) Dark/light open field assay.** Mice with high recombination rates (∼90%, fourth subgroup) strongly avoided walking in the open field, which reflects severe anxiety-related behavior. In contrast, the motor performance of mice with low recombination rates (30–50%, third subgroup) was comparable to controls. One-way ANOVA with Tukey post-hoc test. *n* = 24 *Psmf1*^fl/fl^ Ai14/+ tamoxifen, *n* = 6 *Psmf1*^fl/fl^ Ai14/+ UBC-Cre-ERT2/+ tamoxifen low recombination rate, and *n* = 5 *Psmf1*^fl/fl^ Ai14/+ UBC-Cre-ERT2/+ tamoxifen high recombination rate. **p < 0.01, ***p < 0.001, **\*\*\*\***p < 0.0001. **(f)** Sagittal brain sections stained with either Iba1 (green) for microglia or GFAP for glia (astrocytes) and with Hoechst 33342 for nuclei (blue). TdTomato (red) fluorescence indicates recombination efficiency. **(i)** In the deep cerebellar nuclei (DCN), reactive microglia in tamoxifen-induced mice indicated gliosis induction. Scale bar = 100 μm. **(ii)** Higher magnification in the DCN shows morphologically distinct ramified microglia with more amoeboid appearance upon loss of Psmf1. Scale bar = 20 μm. **(iii)** In the DCN of tamoxifen-induced mice, we observe an increase in glia staining strongly for GFAP in the DCN, a marker for astrogliosis (of note, GFAP also stains the Bergmann glia in the molecular layer). Scale bar = 100 μm. **(iv)** Higher magnification image of **(iii)**. Scale bar = 20 μm. **(g)** Quantification of percent area positive for signal of either GFAP or Iba1 staining in the DCN. T-test, *n* = 3 *Psmf1*^fl/fl^ Ai14/+ UBC-Cre-ERT2/+ corn oil and *n* = 3 *Psmf1*^fl/fl^ Ai14/+ UBC-Cre-ERT2/+ tamoxifen. *p < 0.05, ***p < 0.001.

#### Behavioral tests

##### Rotarod assay

We first assessed motor performances of *Psmf1* CKO mice using the rotarod test (Fig.6b-c). *Psmf1*^fl/fl^ Ai14 UBC-Cre-ERT2 mice were injected with tamoxifen or corn oil as control at 6-7 weeks of age, while *Psmf1*^fl/fl^ Ai14 mice without Cre were injected with tamoxifen as an additional control for tamoxifen. Ten days after the last injection, mice were trained for four days on the rotarod and then tested once on each day after for 11 days (Fig.6b-c) and thrice on the subsequent day. Initially the performance of tamoxifen-injected *Psmf1*^fl/fl^ Ai14 UBC-Cre-ERT2 mice with a high recombination rate (>90% of neurons as assessed by Ai14) displayed a deficit by day 12, whereas tamoxifen-injected *Psmf1*^fl/fl^ Ai14 UBC-Cre-ERT2 mice with a low recombination rate (approximately 30% of neurons as assessed by Ai14) showed only a mild deficit (Fig.6b-c).

##### Open field assay

Since individuals with *PSMF1*-related PD or parkinsonism present with severe anxiety, we also tested *Psmf1*^fl/fl^ Ai14 UBC-Cre-ERT2 mice in the open field assay with dark insert. Anxiety-like behavior of rodents is frequently accompanied by reduced exploration. On day 26 after tamoxifen injection, *Psmf1*^fl/fl^ Ai14 UBC-Cre-ERT2 mice displayed a clear difference to controls, consistent with increased anxiety (Fig.6d-e-f).

#### Brain immunohistochemistry

We observed gliosis in various brain regions 28 days after tamoxifen injection in *Psmf1*^fl/fl^ Ai14 UBC-Cre-ERT2 mice. Many neurodegenerative diseases are associated with gliosis, where glial cells become reactive and proliferate generally prior to the formation of aggregates, tangles and plaques, including PD.^54-58^ Significantly, we detected both microgliosis, as documented by increased staining of microglia with anti-Iba1 and a change in morphology of the microglia from ramified to more amoeboid, and astrogliosis, as demonstrated by elevated GFAP-staining of astrocytes (Fig.6g; Supplementary Files 10-11).

## DISCUSSION

Here we have provided extensive phenotypic, genetic and functional evidence to establish *PSMF1* as a novel human disease gene implicated in parkinsonism, childhood neurodegeneration, and perinatal lethality. Specifically, we reported 17 pedigrees in which biallelic *PSMF1* missense and loss-of-function variants co-segregated with a clinical spectrum ranging from early-onset PD resembling well-established recessive monogenic forms (e.g. *PRKN*, *PINK1*, *DJ-1*, *FBXO7*) to perinatal lethality with neurological manifestations. Notably, we observed a clear genotype-phenotype correlation. Individuals carrying *PSMF1* missense variants had normal neurodevelopmental milestones and manifested with parkinsonism between the first and fifth decade of life. Their clinical picture often encompassed mild pyramidal tract involvement, early dysphagia and psychiatric manifestations, and severely abnormal presynaptic dopaminergic imaging was consistent with nigro-striatal degeneration. These individuals showed various response to antiparkinsonian therapies and developed early motor and non-motor fluctuations and dyskinesia. Notably, the observation of preserved ^123^I-MIBG myocardial uptake and negative α-syn RT-QuIC assays in individuals with *PSMF1*-related PD/parkinsonism parallels the biomarker pattern observed in other early-onset monogenic forms of PD, such as those associated with variants in *PRKN* and *PINK1*.^59-62^ This observation supports the hypothesis that *PSMF1*-related PD/parkinsonism may result from alternative molecular pathways that do not (primarily) involve α-synuclein aggregation. Although we did not observe an enrichment of rare coding and splice variants in *PSMF1* among non-familial PD cases compared to controls in the UKB and AMP-PD datasets, further screening of large-scale PD repositories - including familial cases and regulatory variants - is necessary to better establish the frequency of this disorder and the potential contribution of heterozygous *PSMF1* variants to PD risk. Moving forward into the disease spectrum, patients harboring biallelic *PSMF1* splice variants exhibited parkinsonism and/or other movement disorders and neurological manifestations since childhood or adolescence. Most often, these occurred in the context of neurodevelopmental delay and showed rapid progression leading to death within the second decade. Finally, subjects with two *PSMF1* loss-of-function variants, of which at least one was predicted to abolish *PSMF1* expression, presented with neurological manifestations and structural brain abnormalities since the prenatal period and died within infancy. Intriguingly, *PSMF1* loss-of-function variants were almost invariably associated with agenesis or hypoplasia of the corpus callosum, which consists of millions of axons connecting the cerebral hemispheres. Since Psmf1 has been demonstrated to mediate neurosome-to-synapse fast transport and be crucial for synapse maintenance and neuronal survival in *Drosophila* and mice,^20,22^ it is tempting to speculate that loss of PSMF1 in humans affects axonal pathways leading to failed axonogenesis/synaptogenesis or premature degeneration of these neural structures, which would fuel the notion of altered neurodevelopment as primer of neurodegeneration.

*PSMF1* encodes the evolutionarily highly conserved proteasome regulator PSMF1/PI31, which is ubiquitously expressed in human tissues, including the brain. *PSMF1* shows strong protein expression in fetal brain tissue compared to non-brain tissues and elevated gene expression during the prenatal period across all brain regions.^63^ This could explain the widespread brain effects of its deficiency, not limited to the dopaminergic system. Notably, deep phenotyping of our study cohort revealed a tendency towards obesity and glucose intolerance, early osteoporosis/osteopenia and severe infections. Further delineation of *PSMF1*-related disorder will clarify whether PSMF1 deficiency might have multisystemic involvement in humans.

Our findings suggest that different *PSMF1* mutant alleles drive disease via multiple loss-of-function mechanisms. In particular, the three missense variants p.Arg242Gly, p.Arg242Cys and p.Arg242His detected in six pedigrees with early-onset PD or parkinsonism affect a highly conserved residue within the natively disordered proline-rich C terminal of PSMF1. *In vitro* characterization of the complex human PSMF1/bovine 20S proteasome has demonstrated that PSMF1 Arg242 is inserted into the proteasome catalytic site, which results in an inhibitory mechanism.^64^ Considering the evidence of PSMF1 acting as a 20S proteasome inhibitor and a 26S proteasome upregulator *in vitro* as well as a proteasome enhancer *in vivo*, further studies are needed to establish whether variants at the Arg242 residue determine loss of function by converting PSMF1 from a proteasome inhibitor to a proteasome substrate, or by disrupting proteasome positive regulation, thus supporting the role of proteasome inhibition in the pathogenesis of PD. Similarly, the missense variant p.Leu53Met, identified in two consanguineous families descended from a distant common ancestor, is located at the dimerization interface of PSMF1 and is likely to disrupt hydrophobic interactions within the PSMF1 homodimer, possibly influencing its activity on the proteasome and the stability of the PSMF1-FBXO7 heterocomplex. Finally, *PSMF1* predicted and proven loss-of-function variants in our cohort are associated with moderate-to-severe phenotypes within the reported spectrum. We acknowledge that the lack of biological material beyond DNA from probands E-II-4 and H-II-1 precluded functional validation of the pathogenicity of their private *PSMF1* missense variants, c.725G>A p.(Arg242His) and c.682C>G p.(Leu228Val). Additionally, further analysis is needed to establish why biallelic *PSMF1* splice variants lead to a milder, non-perinatal lethal phenotype compared to *PSMF1* putative or confirmed complete loss-of-function variants, particularly for those splice variants without evidence of alternative splicing from RNA-seq data.

PSMF1 is a high-affinity binding partner to FBXO7 in *Drosophila* and mammalian cells.^26,27^ Phenotypic features of *PSMF1* patients are reminiscent of those of *FBXO7*-related disorder, such as a clinical spectrum including juvenile-onset PD/parkinsonism, pyramidal tract signs only, or a combination thereof, initial response to levodopa and early occurrence of levodopa-induced dyskinesia.^25^ Human neuropathology is not available for *PSMF1*- and *FBXO7*-related disorders, but dopaminergic imaging indicates dramatic reduction in the nigrostriatal dopaminergic innervation in both disorders.^25^ None of the *PSMF1* variants identified in our cohort is predicted to affect amino acid residues at the interface between PSMF1 and FBXO7. Therefore, it remains elusive whether PSMF1 deficiency induces PD/parkinsonism *per se*, or by secondary disruption of FBXO7 functions in humans. Recently, ablation of the FBXO7-PSMF1 interaction by an *FBXO7* missense variant has been demonstrated to affect expression and stabilization of these proteins as well as proteasomal and mitochondrial pathways in human fibroblasts.^27^ Importantly, *Fbxo7* variants cause site-specific proteolytic cleavage and reduced *Psmf1* activity in *Drosophila* and mice.^18,27,28^ Moreover, transgenic *Psmf1* expression in *Fbxo7*-null mice can robustly suppress neurological defects.^21^ This suggests that loss of Fbxo7 causes disease to a considerable extent by attenuating Psmf1 functions. Since FBXO7 acts synergically with PINK1 and PRKN to regulate basal mitochondrial functions and stress-induced mitophagy,^65,66^ we explored mitochondrial pathways by live imaging of *PSMF1* patient-derived fibroblasts and documented mitochondrial dysfunction and alteration of mitochondrial dynamics and mitophagy. Additionally, we provided preliminary evidence that *PSMF1* variants may affect proteasomal abundance and assembly in human fibroblasts. Further studies are needed to fully characterize the consequences of *PSMF1* variants on mitochondrial homeostasis and, most importantly, to delineate their effects on proteasomal functions, whose disruption is likely to be the primary pathomechanism driving *PSMF1*-related pathology.

Besides documenting a similar age-dependent locomotor dysfunction in *psmf1*- and *prkn*-knockdown *Drosophila*, our study demonstrates that *psmf1* deficiency leads to progressive dopaminergic neurodegeneration, accompanied by brain-wide mitochondrial membrane depolarization in old flies, thus paralleling observations in other *Drosophila* PD models and mitochondrial dysfunctions observed in *PSMF1* human fibroblasts. Comparison between *Psmf1* knockout mouse models (previous data and current study) and humans with *PSMF1*-related disorder reveals striking parallelism. Similarly to *Psmf1*-null mice, which die at advanced embryonic stages,^22^ humans with biallelic complete or putative severe loss-of-function *PSMF1* variants (Pedigrees N-O-P-Q) exhibited severe neurological manifestations and structural brain abnormalities since prenatal development and died within infancy. On the contrary, conditional *Psmf1* knockout mice develop age-dependent motor dysfunction,^22^ which is in keeping with motor phenotypes and the survival into adolescence or adulthood of patients harboring biallelic *PSMF1* splice and missense variants which are predicted to cause partial loss of function. Brain autopsy of our mouse model revealed diffuse micro- and astrogliosis with ameboid microglia, which likely reflects early neurodegeneration stages and the timing at which mice were sacrificed before neuropathology could be more informative. Further studies are needed, including expression of hypomorphic and knockin *Psmf1* variants in mice, to provide detailed insights into the neuropathology of *PSMF1*-related disorder.

In conclusion, this study establishes a causal relationship between biallelic *PSMF1* variants and early-onset PD and neurodegeneration as well as perinatal lethality in humans. Our data reveal impairment of mitochondrial membrane potential, dynamics and mitophagy in *PSMF1* patient-derived fibroblasts, suggesting disruption of mitochondrial pathways as a contributing pathomechanism in PSMF1 deficiency. Additionally, we demonstrate that *Psmf1* inactivation in *Drosophila* and mouse leads to age-dependent motor dysfunction, with wide-brain mitochondrial membrane depolarization and loss of dopaminergic neurons in aged flies as well as diffuse gliosis in mice. Evidence presented herein, along with previous literature, suggests that PSMF1 is a key factor in converging pathways of human neurodegeneration. In support of this, recent findings demonstrate that Psmf1 deficiency in mice can be completely rescued with a *Psmf1* transgene, and transgenic *Psmf1* expression can also robustly suppress phenotypes of *Fbxo7*-knockout mice.^21^ Taken together, our study –when considered alongside these complementary observations–highlights the importance of elucidating the role of PSMF1 in human neurobiology and establishes a proof of concept for the potential exploration of PSMF1 gene therapy as a strategy to treat human neurodegenerative disorders.

## METHODS

Written informed consent for diagnostic and research activities and for publication of clinical information, neuroimages and results was obtained from all study participants or their parents/guardians. Samples and data collection and analysis were not performed blind to the conditions of the experiments. No human fibroblasts, animals or data points have been excluded from the analyses.

### Patient recruitment, phenotypic assessment, sampling and primary dermal fibroblast culture

Clinical and prenatal ultrasound assessments were conducted by experienced pediatric and adult neurologists and obstetrician-gynecologists, respectively. A detailed clinical proforma was completed by recruiting clinicians and clinical geneticists of affected individuals. Neuroimaging was reviewed by an experienced paediatric neuroradiologist (Ma.Se.). Genomic DNA was extracted from whole blood or amniotic fluid cells (proband Q-II-2) and RNA was extracted from whole blood (parents J-I-1, J-I-2) according to standard procedures and protocols. In proband J-II-3, a muscle biopsy from the left anterior thigh was performed according to standard procedures and protocols. Punch skin biopsies of proband A-II-2 and his unaffected parents (A-I-1, A-I-2), proband B-II-3 and her unaffected brother (B-II-1), proband F-II-3 and his unaffected parents (F-I-1, F-I-2) and probands J-II-3, L-II-4 and M-II-6 (Fig.1a) were collected according to standard procedures and protocols for primary dermal fibroblast culture for RNA and protein lysate extraction as well as for live cell imaging experiments. A punch skin biopsy was obtained from proband O-II-5 for primary dermal fibroblast culture to extract protein lysate and perform Western blotting. Punch skin biopsies of individuals B-II-1 and I-II-3 were obtained at a later stage of the study and used for primary dermal fibroblast culture to extract protein lysate and perform Western blotting and proteasome assays. Human primary dermal fibroblasts and SH-SY5Y cells were cultured in Dulbecco’s Modified Eagle Medium (DMEM) supplemented with 10% v/v fetal bovine serum (FBS) and maintained in a humidified tissue culture incubator at 37°C, 5%/95% CO_2_/air mixture. To generate *PSMF1* knockdown SH-SY5Y, 1×10^6^ cells were reverse transfected with 100nM siRNA smartpools (non-targeting, Dharmacon D-001206-13-05; PSMF1 targeting, Dharmacon M-012320-01-0005) in 6-well plate format using 0.3% v/v DharmaFECT1 (Dharamacon T-2001-01). To perform α-syn RT-QuIC analysis, in probands A-II-2, B-II-3 and I-II-3, 3-mm skin punch biopsies were obtained unilaterally, after intradermal anesthesia, from the dorsal cervical region at the C7-C8 dermatomes and from the distal leg 10 cm proximally to the lateral malleolus, according to guidelines.^67^ Skin samples were immediately frozen in dry ice. In proband A-II-2, on the same day of skin biopsy, a lumbar puncture was performed at the L3-L4 level, according to guidelines.^40,68^ CSF was collected in 5 ml polypropylene tubes (Sarstedt, code number 63.504.027), centrifuged at 1000g for 10 min at room temperature, and divided into aliquots. Skin and CSF samples were stored at -80°C until α-syn RT-QuIC analysis was performed.

### Genetic studies

#### Exome and genome sequencing and analysis

ES of individuals A-I-1, A-I-2, A-II-2, C-II-3, D-II-2, I-II-3, L-II-1, M-I-1, M-I-2, M-II-6 was performed by Macrogen (Europe or South Korea) as detailed elsewhere.^69^ Briefly, target enrichment was performed with 2μg genomic DNA using the SureSelectXT Human All Exon Kit version 6 (Agilent) to generate barcoded ES libraries. Libraries were sequenced on the HiSeqX platform (Illumina) with 50x coverage. Quality assessment of the sequence reads was performed by generating QC statistics with FastQC. The bioinformatics filtering strategy included screening for only exonic and donor/acceptor splicing variants. In accordance with the pedigree and phenotype, priority was given to rare variants (<0.01% in public databases, including 1000 Genomes project, NHLBI Exome Variant Server, Complete Genomics, and Exome Aggregation Consortium [ExAC v0.2]) that were fitting a recessive (homozygous or compound heterozygous) or a *de novo* model and/or variants in genes previously linked to movement disorders, neurodevelopmental delay and other neurological disorders. ES of individuals B-II-2, B-II-3, B-II-4, G-II-3 and H-II-1 was performed at the ICM IGenSeq core facility. Exons were captured using the Roche V.3 kit, followed by a massively parallel sequencing on NextSeq500 system (Illumina). Read alignment and variant calling were done using an in-house pipeline. Briefly, FastQC was used to check the quality of the reads and low-quality reads were removed using Trimmomatic. Sequencing data were then aligned to the human reference genome hg19 using the bwa suite36 and variant calling was performed using GATK HaplotypeCaller1. ES data of individuals B-II-2, B-II-3, B-II-4, G-II-3 and H-II-1 were analyzed using VaraFT2 software. We filtered all homozygous variants shared by the two affected individuals that possibly affected the cDNA or localized in the splice site region (-8/+11bp from exon/intron junction), with a MAF <1% in the gnomAD public database and localized in region of loss of heterozygosity. Genome sequencing (GS) of E-II-4 was performed through the NIH facility. Read alignment and variant calling were done using Dragen version 07.021.624.3.10.4 (Illumina). Research for *PSMF1* variants was done using an in-house R script (https://gitlab.com/icm-institute/corti-corvol/variants_tiers_gene). ES of probands D-II-2 and N-II-1 was carried out by CENTOGENE as previously described,^70^ and GS was performed in individual B-II-3 by CENTOGENE as detailed elsewhere.^71^ ES of proband F-II-3 was performed using Agilent SureSelect Human All Exon V6 R2 kit with Illumina, NOVASeq6000 (Macrogen, Korea). Raw data obtained from ES were processed using the SEQ platform (Genomize, Istanbul, Turkey). Burrows-Wheeler Aligner (BWA) was used to align with the human reference genome, hg19. Freebayes was applied for variant calling, and the PCR duplicates were removed using the DeDup tool. ES of J-II-1 and J-II-3 was performed as described elsewhere.^72^ ES of proband K-II-3 and his parents K-I-1 and K-I-2 were performed on a Novaseq 6000 platform (Illumina, San Diego, California) using IDT xGen Exome Research Panel V2 Kit (Integrated DNA Technologies, Coralville, Iowa). The average read depth was at least 100 reads, and the average percentage coverage of Refseq genes at 20X was at least 94%. The bioinformatic analysis was performed using the Franklin variant interpretation platform (Genoox, Tel Aviv, Israel). Variants were filtered based on their effect on the protein (missense, nonsense, frameshift, splice-site) and a minor allele frequency <0.01 in general population databases (e.g. gnomAD), and the Rambam Genetics Institute internal database of over 3000 Israeli samples. ES of individual M-II-3 was carried out as reported previously.^73^ ES of subjects O-II-5, P-I-1, P-I-2 and P-II-3 was performed as detailed elsewhere.^74^ ES of subjects Q-I-1, Q-I-2 and Q-II-2 were carried out as previously reported.^75^ Sanger sequencing was used to confirm next-generation sequencing findings and perform family segregation whenever possible. Recommendations of the Human Genome Variation Society were used to describe the cDNA and protein sequence variants using NM_006814.5 and NP_006805.2 as the reference. All next-generation sequencing findings were verified by Sanger sequencing while performing family segregation analysis in-house or in the referring centers.

#### Frequency, pathogenicity prediction and computational analysis of PSMF1 variants

As a reference for potentially neutral variants and frequency of variants in the healthy population, gnomAD (all released versions) was inspected for all identified *PSMF1* variants (Supplementary File 1). We also screened the 100,000 Genomes Project, UKB, *All of Us* Public Data Browser, and Queen Square Genomics exome repositories for the *PSMF1* variants identified in our study cohort (Supplementary File 1). Pathogenicity prediction of *PSMF1* non-splice variants was assessed using CADD, PolyPhen2 HVAR, SIFT4G, PROVEAN, MutationTaster and Human Splicing Finder (HSF) to predict the impact of variants on the protein structure and function (Supplementary File 1A). To examine the conservation of substituted amino acid positions, we used Genomic Evolutionary Rate Profiling (Supplementary File 1) and visual multiple sequence alignment of PSMF1 across multiple different species by inputting PSMF1 protein sequences for each species retrieved from UniProt in Clustal Omega (Fig.2b).^76^ For *in silico* splice predictions of six splice variants, we used the computational tools SpliceSiteFinder-like, MaxEntScan, NNSPLICE, GeneSplicer, SpliceAI, and AbSplice (Supplementary File 1B).

#### Autozygosity mapping

In all affected individuals having homozygous *PSMF1* variants and their unaffected family members with raw ES or GS data available (Pedigrees B-C-D-E-F-G-H-J-L-M-N-O), regions of homozygosity ≥ 1 Mb were identified using the AutoMap software (https://automap.iob.ch/) by inputting VCF files (Variant Call Format) files (Supplementary File 2).^37^ Whenever needed, we used UCSC LiftOver tool for conversion of hg19 VCF files to hg38 prior to perform autozygosity mapping.

#### Haplotype analysis

Haplotype analysis was performed by plotting a color banding of variants flanking three *PSMF1* variants, each detected in the homozygous state in the affected individuals of two families, including c.724C>G p.(Arg242Gly) in Pedigrees B-C (B-II-3, B-II-4, C-II-3), c.157C>A p.(Leu53Met) in Pedigrees F-G (F-II-3, G-II-3) and c.764+5G>A in Pedigrees L-M (L-II-1, M-II-6). By comparing the banding patterns, the status of being a recurrent or founder variant was investigated. When the banding pattern suggested a founder variant (F-II-3, G-II-3), we supposed the shared region to be an identity-by-descent haplotype and calculated the age of the most recent common ancestor (MRCA) using the ExHap tool (Supplementary File 3).^38^

### RNA studies

#### RNA isolation and sequencing

Total RNA was extracted from fibroblasts of proband A-II-2 and his parents (A-I-1, A-I-2) and probands J-II-3, L-II-4 and M-II-6 using the RNeasy Mini Kit (Qiagen) according to the manufacturer’s protocol. In J-I-1 and J-I-2, total RNA was isolated from peripheral whole blood treated with red blood cell lysis buffer (Qiagen) and white blood cells were collected for RNA extraction. RNA was extracted using TRIzol™ reagent (Thermo Fisher Scientific, Waltham, Massachusetts, USA) according to manufacturer’s instruction. RNA samples with an RNA integrity number (RIN) ≥7 was used for RNA-seq. cDNA libraries were prepared by KAPA mRNA HyperPrep Kit (Roche Diagnostics, Tucson, AZ, USA) according to the manufacturer protocol. Briefly, poly-A containing mRNA was collected and fragmented to 200-300 bp. Fragmented mRNA was then used as template for first-strand cDNA synthesis by random hexamer. In the second strand cDNA synthesis, blunt-end double stranded cDNAs were generated for 3’ adenylation and indexed adaptor ligation. The adaptor-ligated libraries enriched by polymerase chain reaction were sequenced as 151-bp paired-end runs on Illumina® NovaSeq 6000 platform (Illumina, San Diego, CA, USA) at the Center for PanorOmic Sciences (CPOS), University of Hong Kong. Data cleaning of the transcriptomics profile in fastq format was performed by AfterQC (v0.9.6). The sequence was aligned to the hg19/GRCh37 human reference genome and outputted as bam files using STAR (v2.5.2a, https://github.com/alexdobin/STAR) in two-pass mode. RNA sequencing data were processed with DROP v1.3.3 workflow,^77^ which integrates the statistical algorithm OUTRIDER^78^ and FRASER2^79,80^ for detection of aberrant expression (AE) and aberrant splicing (AS) respectively by computing significant levels of extreme read count values. Briefly, the DROP pipeline enables the systematic detection of AE and AS outliers by comparing an individual’s RNA sequencing. AE refers to the differential expression of the two alleles of a gene in a diploid organism. Normally, both alleles are expressed at similar levels. However, one allele can be significantly over- or under-expressed due to coding, non-coding, regulatory, or structural variants. Detecting AE outliers can help identify functional effects of non-coding or regulatory variants, such as promoter or splice mutations. AS refers to abnormal RNA splicing patterns that differ from the expected transcript structure. This can include exon skipping, intron retention, or usage of cryptic splice sites. Since disease-causing variants can disrupt splicing, identifying AS events can reveal the molecular mechanism behind pathogenic variants that may be missed by standard DNA sequencing. Both OUTRIDER^78^ and FRASER2^79,80^ algorithms utilize a denoising autoencoder to control for the hidden confounder effects. In OUTRIDER for detection of aberrant expression, a false discovery rate (FDR) ≤0.05 was used as the cut-off for calling significant aberrant expression outliers.^67^ In FRASER2, split reads spanning the exon–exon junction and nonsplit reads spanning the splice sites were counted for splicing event calling. The Intron Jaccard Index is computed using split and non-split reads for capturing several types of aberrant splicing. Significant aberrant splicing events were defined with FDR ≤0.1. ^79,80^

#### Minigene splicing assay

When biological material for obtaining RNA was not available, an *in vitro* assay targeting each exonic region was performed through amplification from genomic DNA to functionally test the predicted impact of *PSMF1* splice variants, as previously described.^81^ Primers to assay the *PSMF1* c.365+2T>C (exon 3) and c.605+1G>A (exon 5) variants are shown in Supplementary File 12. Following amplification, clean-up and restriction enzyme digestion (*Xho*I, *Bam*HI) of the PCR amplicon and pSPL3 exon trapping vector was performed prior to ligation between the vector-containing exons A and B of the linearized vector that was transformed into DH5α competent cells (NEB 5-alpha, New England Biolabs, Frankfurt, Germany), plated and incubated overnight. Colonies with the wild-type containing sequences were selected for correct size using colony PCR with an SD6 forward and the PSMF1 exon-specific reverse primer. Primers for site-directed mutagenesis were designed with NEBase Changer version 1.3.3 (New England Biolabs, https://nebasechanger.neb.com/). Site-directed mutagenesis followed the manufacturer protocol. Clones were selected for overnight LB-amp culture. Plasmids were isolated and Sanger sequence verified. The sequence-confirmed vectors with mutant and wild-type sequence were transfected into HEK 293T cells (ATCC, Manassas, VA, USA) with a density of 2×10^5^ cells per mL. 2µg of the respective pSPL3 vectors was transiently transfected using 6µL of FuGENE 6 Transfection Reagent (Promega, Walldorf, Germany). Empty vector and transfection negative reactions were included as controls. The transfected cells were harvested 24 hours after transfection. Total RNA was prepared using miRNAeasy Mini Kit (Qiagen, Hilden, Germany). Approximately 2µg of RNA was reverse transcribed using the High-Capacity cDNA Reverse Transcription Kit (Applied Biosystems, Thermo Fisher Scientific, Waltham, MA, USA) following the manufacturer protocols. The cDNA was PCR amplified using vector-specific SD6 forward and SA2 reverse primers. Splicing assay could not be performed for the *PSMF1* c.129+2T>C (exon 1) variant. Since the first exon of a gene does not have a splice acceptor site, we aimed to introduce a variant using site-directed mutagenesis in order to induce a strong splice acceptor site. However, by permutating the bases around the beginning of exon 1 to each A-T-G-C alternate base, we did not encounter a strong candidate that would have caused a new splice acceptor site to trick exon 1 to be included in the minigene. No sequence in that area resembled branch sequence or a polypyrimidine tract.

### Proteomic studies

#### Protein structure modelling

The crystal structure for the PSMF1 homotetramer was accessed through the Protein databank repository, accession number 4OUH.^82^ Images were generated using ChimeraX.^83^ Structural modelling of the FBXO7/PSMF1 heterotetrameric complex was performed using AlphaFold 3^39^ with primary sequences obtained from UniProt: FBXO7 (Q9Y3I1) and PSMF1 (Q92530). Detailed intermolecular interactions were predicted with BIOVIA Discovery Studio Visualizer 2024 using the Non-bond Interaction Monitor. All structures were visualized using PyMOL 3.0.

#### Western blotting

Human primary fibroblasts from probands, unaffected parents and healthy controls, in addition to Non-targeting siRNA and PSMF1 siRNA KD SH-SY5Ys were lysed with ice-cold RIPA-lysis buffer consisting of: 50mM Tris-HCl (pH=7.4), 150mM NaCl, 1% w/v triton-X-100, 0.5% sodium deoxycholate, 0.1% w/v sodium dodecyl sulphate (SDS), 1x PhosSTOP phosphatase inhibitors and 1x cOmplete™, Mini, EDTA-free Protease Inhibitors (all Sigma). 25µg of protein lysate were separated through SDS polyacrylamide gel electrophoresis (PAGE) using 4-12% NuPAGE Bis-Tris Mini Protein Gels and XCell SureLock Mini Cell (Thermo Fisher Scientific), before transfer to 0.45 µm Immobilon®-FL PVDF membrane (Sigma) using Mini Trans-Blot Cell (Bio-Rad). Blots were blocked with 5% w/v milk in PBS with 0.01% tween-20 (PBST; 1h, room temperature (RT); Sigma). Blots were incubated with the following primary antibodies diluted in 3% w/v milk in PBS-T (4°C, overnight): mouse anti-GAPDH IgG2b (1:10000; Abcam Cat# ab110305, RRID:AB_10861081) and rabbit anti-PSMF1 IgG (1:1000; Invitrogen Cat# PA5-21603, RRID:AB_11153838). Blots were then washed 3x with PBST prior to incubation with fluorescently conjugated secondary antibodies diluted in PBST supplemented with 0.02% SDS (2h, RT): donkey anti-mouse IgG Alexa Fluor Plus 680 (1:40,000; Invitrogen Cat# A32788, RRID:AB_2762831) and donkey anti-rabbit IgG Alexa Fluor Plus 800 (1:40,000; Invitrogen Cat# A32808, RRID:AB_ 2762837). Blots were washed 3x with PBST and 1x with PBS prior to near-infrared (NIR) detection using an Odyssey CLx imager (LI-COR Biosciences, RRID:SCR_014579). Protein lysates were isolated from HepG2 cells, control-derived fibroblasts and proband O-II-5-derived fibroblasts (with homozygous *PSMF1* variant c.1A>T) using RIPA buffer (Thermo Fisher Scientific) according to the manufacturer protocol. 30μg protein lysates were separated by PAGE (18% gel), semi-dry blotted and incubated with a polyclonal anti-PSMF1 IgG Ab (Thermo Fisher Scientific cat. no. PA5-21603) at 4°C overnight. After washing and a 1-hour incubation with goat anti-rabbit IgG HRP, the blot was washed again before being developed with ECL-plus (Thermo Fisher Scientific) for 3 minutes. After stripping, the same blot was incubated with monoclonal β-actin antibody, washed and incubated with rabbit anti-mouse IgG HRP before being developed for 2 minutes with ECL-plus.

#### Quantitative proteomics in human cultured dermal fibroblasts

##### Sample preparation

Proband- and carrier-derived cultured dermal fibroblasts were harvested in lysis buffer containing 2% SDS (w/v), 100mM TEAB pH 8.5, with complete EDTA-free protease inhibitor cocktail (Roche) and PhosSTOP phosphatase inhibitor cocktail tablets (Roche). Protein concentrations were determined via BCA assay (Thermo Fisher Scientific) and 5µg protein aliquoted per sample for proteomic analysis. Samples were reduced with 10mM TCEP for 30 minutes at 60°C, followed by alkylation of cysteine residues with 40mM IAA for 30 minutes at room temperature in the dark. 20% SDS was then added to achieve a final concentration of 5% SDS in samples, which were then acidified by addition of trifluoracetic acid to a final concentration of 1%. Samples were subsequently processed for on-column tryptic digestion using S-Trap^™^ micro columns (Protifi, USA).^84,85^ Briefly, samples were diluted 6x with wash buffer (90% methanol, 10% 100mM TEABC) and loaded onto micro columns, with centrifugation at 1000g for 1 minute and flow-through discarded. After sample loading, the S-Trap^™^ micro columns were washed four times with 150μl wash buffer followed by centrifugation at 1000g, 1 minute. On-column digestion was performed by incubating 40μl (1μg) of Trypsin/Lys-C mix (MS grade, Promega, UK) in 50mM TEABC solution (pH 8) at 47°C for 1 hour and 20 minutes, followed by incubation at 22°C overnight. The samples were then eluted with addition of 40μl 50mM TEABC (pH 8), 40μl 0.15% (v/v) formic acid (FA), and 3x 40μl 80% acetonitrile (ACN), 0.15% FA. Peptides were then dried using a vacuum centrifuge at room temperature and stored at -20°C until mass spectrometry analysis.

##### Quantitative proteomic analysis

Peptides were resuspended in 0.1% formic acid supplemented with 0.015% N-Dodecyl-B-D-Maltoside (DDM). 200ng of peptides were analyzed with a Vanquish Neo nano-liquid chromatography system in-line with an Orbitrap Astral mass spectrometer (Thermo Fisher Scientific). Peptides were trapped and eluted using an Acclaim™ PepMap™ 100 C18 HPLC column (3μm particle size, 75μm diameter, 150mm length) and separated using an EASY-Spray™ PepMap™ Neo UHPLC column (2μm C18 particle, 75μm diameter,150mm length) with buffer A: 0.1% formic acid and buffer B: 80%ACN, 0.1% formic acid. Peptides were separated across a 13-minute gradient, and the data was acquired using narrow window DIA mode. MS1 scans performed at a resolution of 240,000 across a m/z range of 380-980 and measured using ultra-high field orbitrap mass analyzer, with a normalized AGC target of 500% and maximum injection time of 3 milliseconds. DIA scans were performed of the precursor mass range with an isolation window of 4m/z, scan range of 150-2000m/z and a normalized collision energy of 25%.

##### Raw mass spectrometry data analysis

Raw data were searched using DIA-NN (version 1.8.1) against a predicted spectral library generated from the reviewed human UniProt database (downloaded January 2023; 20383 entries with isoforms). Spectra were searched with strict Trypsin specificity (cleavage at K or R residues), allowing a maximum of two missed cleavages. Peptides with an amino acid length of 7-30 were considered. N-terminal methionine excision and oxidation were specified as variable modifications and cysteine carbamidomethylation as a fixed modification, with a maximum of one variable modification permitted.

##### Statistical analysis and data visualization

Statistical analysis and visualization of DIA-NN output files was performed with Python (version 3.11.5, Jupyter Notebook; Project Jupyter) using packages pandas (2.0.3), matplotlib (3.7.2), seaborn (0.12.2), numpy (1.26.1) and sci-kit learn (1.3.0); before upload to CURTAIN web tool (https://curtain.proteo.info/#/, Supplementary File 13). Proteins identified by a single peptide were excluded, with log2-transformed protein intensities then filtered by 100% detection in at least one sample condition. Missing values were imputed by random draws from a Gaussian distribution (width 0.3 and downshift 1.8). The Limma package (version 3.56.2; R, version 4.3.1) was used to fit the data to a linear model using the lmfit function, with eBayes correction to compute moderated t-statistics and false-discovery rate controlled by Benjamini-Hochberg correction. All mass spectrometry proteomics data have been deposited to the ProteomeXchange Consortium via the PRIDE partner repository with the dataset identifier: PXD053147.

### Frequency of *PSMF1* variants in the UK Biobank and Accelerating Medicines Partnership-Parkinson’s Disease Initiative and gene burden

We searched for PSMF1 missense (nonsynonymous, nonframeshift indels) and loss-of-function (splice, nonsense, frameshift) variants in the ES data within the UK Biobank (UKB, https://www.ukbiobank.ac.uk/)^86^ and extracted exonic variants from GS data from the Accelerating Medicines Partnership-Parkinson’s Disease Initiative (AMP-PD, https://amp-pd.org/).^87^ Although we acknowledge the importance of investigating regulatory variants, our analysis focused on coding variants to prioritize those with clearer functional consequences, in line with the objectives of a gene discovery study.

Processing of these cohorts was detailed elsewhere.^88^ In brief, the UKB is a large-scale biomedical dataset with clinical and genetic information from ∼500,000 participants.^86^ After filtering and removing those recruited in a genetic study, the database consists of a total of 45,857 individuals (22,040 males), of which 38,051 are controls, 1,105 cases, 6,033 individuals with a parent that is diagnosed with PD, and 668 individuals with a sibling that is diagnosed with PD (6,701 proxy cases). Among controls, individuals with age at recruitment <59 years, any reported neurological disorders (category 2406 and field codes: Dementia/42018, Vascular dementia/42022, FTD/42024, ALS/42028, Parkinsonism/42030, PD/42032, PSP/42034, MSA/42036), a parent with PD or dementia (field codes: 20107 and 20110) were filtered out.^86,88^

GS data from multiple datasets, including the Parkinson’s Progression Markers Initiative (PPMI), the Parkinson’s Disease Biomarkers Program (PDBP), the Harvard Biomarker Study (HBS), BioFIND, SURE-PD3, and STEADY-PD3, were obtained as part of the Accelerating Medicines Partnership in Parkinson’s Disease (AMP-PD) initiative. The AMP-PD cohorts (PPMI, PDBP, HBS, BioFIND, SURE-PD3, and STEADY-PD3) followed the GATK Best Practices guidelines established by the Broad Institute’s joint discovery pipeline, as well as additional details provided elsewhere.^87^ Data processing and quality control (QC) procedures have been described previously.^87,89^ All individuals included in the analysis were of European ancestry through principal component analysis using HapMap3 European ancestry populations. This selection was motivated by the predominance of genetic data from European individuals in the datasets and was made to ensure consistency and minimize bias due to population stratification. The AMP-PD datasets consist of unrelated individuals and publicly available cohorts only. Similar filtering parameters were used for the AMP-PD databases resulting in a total of 4,007 individuals (2,197 males), with 2,556 being controls and 1,451 being PD cases.^87,88^

Analyses were restricted to non-familial PD cases, who make up the majority of available data in both cohorts (AMP-PD, UKB). Familial PD cases are limited in number, and their inclusion would have not substantially increased statistical power for this rare-variant analysis. Variants were annotated using ANNOVAR, including the databases refGene, avsnp150, and clinvar_20220320.^90^ Variants were then selected/filtered for nonsynonymous (missense), stopgain, stoploss, nonframeshift, frame, splice (loss of function). For both analyses, we applied an upper MAF limit of 0.005, which captured most *PSMF1* variants (Supplementary File 6). Raw genotype data files were generated to retrieve information on the allelic status (heterozygous, homozygous for reference allele, homozygous for non-reference allele). Following variant extraction, single variant association tests were performed using PLINK v1.90b4.4 (http://pngu.mgh.harvard.edu/purcell/plink/).^91^ To assess whether the average effect of genetic variants in this gene is associated with PD, burden testing was performed using Sequencing Kernel Association optimal unified test (Skat-O), Combined Multivariate and Collapsing (CMC) Wald test and CMC burden testing. This process, including variant annotation using SnpEff and SnpSift softwares, and the Loss-of-Function (LoF) Transcript Effect Estimator (LOFTEE) has been previously described.^88^

### α-Synuclein real-time quaking-induced conversion assay

Punch skin biopsies were processed as detailed elsewhere.^92,93^ Briefly, thawed skin samples were washed 3 times in cold 1X phosphate-buffered saline (PBS) and homogenized at 1% (wt/vol) using a gentleMACS Octo Dissociator (Miltenyi Biotec, Bergisch Gladbach, Germany) in cold 1X PBS supplemented with Complete Protease Inhibitor Mixture (Roche, Basel, Switzerland). Skin homogenates were centrifuged at 1,000g for 10 minutes at 4°C, and supernatants were stored at -80°C until use. We performed α-syn RT-QuIC as previously described.^40,92,94^ Briefly, we preloaded black 96-well plates with a clear bottom (Nalgene Nunc International, Rochester, NY) with six 0.8-mm silica beads (OPS Diagnostics, Lebanon, NJ) per well. CSF and skin samples were thawed and vortexed before use. For the CSF assay, 15 μL of sample were added as seed to 85 μL of reaction mix containing 40 mM phosphate buffer, pH 8.0, 170 mM NaCl, 10 μM thioflavin-T, 0.0015% sodium dodecyl sulfate, and 0.1 g/L recombinant α-synuclein filtered with a 100-kDa-molecular-weight cutoff filter (Amicon centrifugal filters; Merck Millipore, Burlington, MA). For the skin assay, 2 μL of skin homogenate at 10^-2^ final dilution were added to 98 μL of reaction mix containing 40 mM phosphate buffer, pH 8.0, 170 mM NaCl, 10 μM thioflavin-T, and 0.1 g/L of recombinant α-syn which was produced in house.^94^ The plate was loaded, sealed with a plate sealer film (Nalgene Nunc International) and incubated into a FLUOstar Omega (BMG LABTECH, Ortenberg, Germany) plate reader at 42°C with intermittent double-orbital shaking at 400 rpm for 1 minute, followed by a 1-minute rest. Thioflavin-T fluorescence was measured every 45 minutes using 450-nm excitation and 480-nm emission filters. We ran 4 replicates per sample. Replicates were deemed positive if at least 2 of the 4 replicates gave a fluorescence signal higher than the positivity threshold, arbitrarily set at 30% of the median I_max_ value of the positive control replicates during the 30-hour run. RT-QuIC fluorescence responses were analyzed and plotted using GraphPad Prism 9.0 for Windows.

### Proteasome activity assays

Native gel analyses were performed to evaluate the relative abundance of different proteasome particles as previously described.^19^ 5 to 10 µg of extracts from *PSMF1* patient-derived fibroblasts and healthy controls dissolved in native sample buffer (Bio-Rad, 1610738) and separated on 3-8 % Criterion XT Tris-Acetate gels (Bio-Rad) under nondenaturing conditions and subsequently transferred onto PVDF membrane for standard Western blot analysis. In parallel, samples were also analyzed by standard Western blot analysis. The different proteasome particles were detected with an anti-20S Alpha7 subunit antibody (Enzo, BML-PW8110) that recognized the 20S core particle and its complexes with 19S regulatory particles (26S). PSMF1 was detected with either an anti-PSMF1 antibody (Abcam, ab96845) or anti-PSMF1 antibody (Invitrogen, PA5-21603). β-ACTIN was detected with an Anti-β-ACTIN (13E5) antibody (Cell Signaling Technology, 5125S).

### Mitochondrial studies in human cultured dermal fibroblasts

Live cell imaging of cultured dermal fibroblasts for studying mitochondrial bioenergetic state and function was performed in cell donors from Pedigrees A-B-F-J-L-M (Fig.1a). Images were acquired using Zeiss LSM 980 and LSM 710 confocal microscopes equipped with META detection system and 40× oil immersion objective. Images were analyzed using ZEN 2009 software and Fiji 2 software.

#### Measurement of mitochondrial membrane potential (MMP) using TMRM

For measurements of MMP, dermal fibroblasts plated on 25-mm glass coverslips were loaded for 40 minutes at room temperature with 25nM tetramethylrhodamine methylester (TMRM; Invitrogen), a cationic fluorescent dye that enters healthy mitochondria, in a HEPES buffered saline solution (HBSS) composed of 156mM NaCl, 3mM KCl, 2mM MgSO4, 1.25mM KH2PO4, 2mM CaCl2, 10mM glucose and 10mM HEPES; pH 7.35. The dye remained present in the media at the time of recording. Confocal images were obtained using a Zeiss 980 LSM Airy or 710 VIS CLSM equipped with a META detection system and a 40× oil immersion objective. TMRM was excited using the 561 nm laser line and fluorescence was measured above 580nm. For basal MMP measurements, Z-stack images were obtained by confocal microscopy and analyzed using ZEN 2009 software (Zeiss). TMRM values for control cases were set to 100% and the PSMF1 deficient and family members fibroblasts values were expressed relative to controls. For analysis of response to mitochondrial toxins, images were recorded in a time course-dependent manner from a single focal plane and analyzed using ZEN Zeiss software (Zeiss).

To analyze the maintenance mechanism of MMP, the response to mitochondrial toxins (oligomycin, rotenone FCCP) was studied. TMRM was used in the redistribution mode to assess MMP, and therefore a reduction in TMRM fluorescence represents mitochondrial depolarization. Basal TMRM levels = 100%. TMRM fluorescence after complete depolarization by FCCP = 0%.

#### Dynamics

We further studied the mitochondrial dynamics in the TMRM z-stacks by using the Mitochondrial Network Analysis (MiNA) toolset, a simplified pipeline to analyze the mitochondrial network morphology and fragmentation using fluorescence images and three-dimensional stacks using Fiji 2 software following the creators’ protocol,^95^ we studied the average length of all mitochondrial rods and branches in each cell (Fig.4d) and the number of branches per network in each cell (Fig.4e).

#### Mitophagy

To study mitophagy via mitochondrial and lysosomal co-localization, cells were loaded with 200nM MitoTracker Green FM and 50nM LysoTracker Red in HBSS for 30 minutes before experiments. Confocal images were obtained using a Zeiss 710 confocal microscope equipped with 40X oil immersion objective. The 488nm argon laser line was used to excite MitoTracker Green fluorescence which was measured between 505 and 530nm. Illumination intensity was kept to a minimum (about 1% of laser output) to avoid phototoxicity and the pinhole set to give an optical slice of approximately 2μm. For LysoTracker Red, the 543nm Ne/He laser line was used with measurement above 650nm. All data presented were obtained from a minimum of five coverslips and three different cell preparations.

### *Drosophila melanogaster* modelling and experiments

#### Drosophila stocks

In *Drosophila*, targeted knockdown of *psmf1* and associated positive/negative controls was achieved using the UAS-Gal4 system. We obtained targeted knockdown of *psmf1* expression in conjunction with the UAS-Gal4 system of cell-type-specific transgene expression by RNA-interference (RNAi). Transgenic RNAi flies were generated according to a previously published protocol.^96^ Orthologues were identified through the “DIOPT ortholog finder”,^97^ with *psmf1* (FBgn0033669), *prkn* (FBgn0041100) and *nsmase* (FBgn0035421) being considered as fly orthologues of human *PSFM1*, *PRKN* and *SMPD2*, respectively. Transgenic flies were obtained from the Vienna Drosophila Resource Center (VDRC) Stock Center. We crossed the RNAi lines *psmf1* (VDRC ID: 28858), *prkn* (VDRC ID: 47636) and *nsmase* (VDRC ID: 107062) with an nSyb-Gal4 strain to drive pan-neuronal RNAi expression. Growing flies were exposed to a standard 12h:12h light-dark schedule at 25°C and 60% relative humidity.

#### Behavioral test (negative geotaxis assay)

Adult flies of the selected genotypes were tested for behavioral analyses at Days 10, 20 and 35. Based on previous studies and behavioral tests, *prkn* and *nsmase* were considered as positive and negative control, respectively. We assayed *Drosophila* locomotor activity using the climbing test based on negative-geotaxis analysis. Three experimental replicates were performed, each test including at least 15 flies. The test was performed on all lines five times at each time point after a 1-minute rest period. Only females were tested. Flies failing to cross the tube at its midpoint (5-cm) within 10 s from stimulation (repulsion) were interpreted as having a motor impairment. ImageJ software was used to automatically count flies.^98^ The experiment was performed in triplicate (*n* = 3), with each test including at least 15 flies. All lines were tested five times at each time point, following a 1 -min rest period. Only female flies were used in the assay. Flies that did not reach the middle of the tube (5 cm) within 10 seconds after stimulation were considered to exhibit locomotor impairment. Fly counts were automatically performed using ImageJ. Statistical analyses related to the negative geotaxis assay were conducted in R (version 4.1.2) using native packages. Comparisons between two genotypes were performed using the χ² (chi-squared) test.

#### Measurement of mitochondrial membrane potential using TMRM

Flies of the genotype and age indicated were dissected in phosphate buffered saline (PBS: Sigma-Aldrich). Whole brains were incubated in 100nM TMRM in PBS for 30 minutes, and imaged immediately using a Zeiss LSM 710 confocal microscope with an EC ‘Plan-Neofluar’ 20x/0.50 M27 air objective, taking z-stacks through the entire brain with step sizes of 1-2 μm. Analyses were performed in ImageJ: ROIs were drawn around the central brain and mean fluorescence with a background subtraction applied was taken.

#### Immunostaining and TH+ neuron imaging

Flies of the age and genotype indicated were dissected in PBT. Whole brains were fixed with a 20-min room temperature incubation in 4% paraformaldehyde (MP biomedicals) and blocked with a one-hour room temperature incubation in 5% normal goat serum in PBS + 0.3% Triton-X (Sigma-Aldrich). Brains were incubated overnight in primary antibody, washed three times in PBT, and incubated overnight in secondary antibody. Primary antibody was rabbit anti-tyrosine hydroxylase (Abcam) at 1:500, and secondary was AlexaFluor Goat anti-rabbit 647 (ThermoFisher Scientific) at 1:1000. Brains were mounted in SlowFade (ThermoFisher Scientific) and imaged as above. TH^+^ cell bodies were manually counted in ImageJ while blinded to genotype.

### *Mus musculus* modelling and experiments

#### Mice

*Psmf1* knockout-first (KOF) mice were bred with a strain expressing FLP1 recombinase to generate a *Psmf1* allele with the third exon flanked by 2 loxP sites (*Psmf1*^fl^ and *Psmf1*^tm1c(EUCOMM)Hmgu^) as previously described.^22^ *Psmf1*^fl/fl^ mice expressed normal levels of Psmf1, were viable and fertile, and appeared phenotypically normal. However, introduction of a Cre driver successfully inactivated *Psmf1* and resulted in tissue-specific or time-specific loss of Psmf1 protein. UBC-Cre-ERT2 (007001) and Ai14 (007914) mice were purchased from Jackson Laboratories. All experiments in *Mus musculus* were performed in the USA as required by the United States Animal Welfare Act and NIH’s policy to ensure appropriate care and use of laboratory animals for research, and according to established guidelines and supervision by the Institutional Animal Care and Use Committee of The Rockefeller University. Mice were housed in accredited facilities of the Association for Assessment of Laboratory Animal Care in accordance with NIH guidelines.

#### Tamoxifen injection

Tamoxifen was dissolved in corn oil overnight at 37°C to a final concentration of 20mg/ml. Injection dose was determined by weight, using approximately 75mg tamoxifen/kg body weight (about 94-100μl of tamoxifen solution for an adult mouse). Tamoxifen was administered by intraperitoneal injection into 6 to 7-week-old mice for 3 consecutive days. During the injection period and for one week after, the mice were monitored for any adverse reaction due to the treatment.

#### Behavioral tests assessing locomotion and anxiety-related behavior

##### Rotarod test

We used the rotarod test to assess the motor performance of mice on a rotating accelerating rod (Ugo Basile model). The time-to-fall (rotarod score), i.e., the cumulative time (seconds) a mouse maintained its balance on the rotarod before falling off, was recorded. The maximum latency was set at 300 seconds per trial. A fall was recorded if the mouse held on to the rod for two consecutive revolutions. Mice treated with tamoxifen or corn oil (controls) started a four-day training period on the rotarod 10 days after the final injection. On day 14 post injection, the time-to-fall for one trial was recorded. The trial was repeated for 10 days to follow the progressive deficit in the tamoxifen treated *Psmf1* UBC-Cre-ERT2 mice. On day 25 post injection, three trials were conducted for each mouse and average latency to fall was determined.

##### Open field test

We assessed motor activity as a reaction to an unknown environment (anxiety-related behavior). At 26 days post tamoxifen or corn oil injection mice were assayed for locomotion and anxiety in an open field chamber (42 × 42 × 30.5 cm) with a dark enclosure (half of open field chamber) for 10 minutes. Data were analyzed with Fusion 3.2 software (Open field chamber and software from AccuScan Instruments Inc.). Briefly, mice were moved to testing room 60 minutes prior to testing to acclimate. Then, mice were placed in the dark enclosure, and activity was recorded for 10 minutes.

##### Statistics

We use t-test for pairwise comparisons and one-way ANOVA with Tukey post hoc test for when more than two groups were compared. These were calculated using GraphPad Prism software.

#### Immunofluorescence of mouse brain sections

At Day 28 after final tamoxifen injection, mice were perfused with phosphate-buffered saline (PBS) followed by 4% paraformaldehyde (PFA) in PBS. Brains were removed and postfixed in 4% PFA PBS overnight at 4°C. They were then washed in PBS, soaked in 15% sucrose in PBS overnight at 4°C or until they sank, and then soaked in 30% sucrose in PBS overnight at 4°C. The brains were embedded in Richard-Allan Scientific Neg-50 Frozen Section Medium, and 50-μm sagittal sections were made using a Leica CM 3050S cryostat. The floating cryostat sections were permeabilized and blocked overnight in PBS with 0.3% Triton X-100 and 5% Normal Donkey Serum (NDS) at 4°C. The sections were then incubated overnight with primary antibodies diluted in PBS with 0.3% Triton X-100 and 5% NDS. Subsequently, the sections were washed 3 times for 20 minutes each in PBS with 0.1% Triton X-100 and incubated overnight with Alexa Fluor conjugated secondary antibodies diluted in PBS with 0.3% Triton X-100 and 5% NDS at 4°C. The sections were then washed for 30 minutes 4 times in PBS with 0.1% Triton X-100 and mounted with Molecular Probes ProLong Diamond antifade mounting medium. Images were captured on a Zeiss confocal LSM 780 with Zen software. Primary antibodies used were anti-Tyrosine Hydroxylase (TH) Rb pAb (1/500) Abcam ab112, anti-Tyrosine Hydroxylase (TH) Mouse mAb IgG1κ (clone LNC1) (1/500) Millipore MAB318, mCherry Chicken IgY (1/1000) Novus NBP-2-25158, GFAP chicken IgY (1/2000) EnCor CPCA-GFAP, GFAP rabbit IgG (1/500) Dako Cytomation Z0334, Iba1 rabbit IgG (1/1000) Wako 019-19741 and SQSTM1/p62 mouse IgG2a (1/500) Abcam ab56416. For the GFAP and Iba1 quantification, percent area in a 45.16 mm squared area of a maximum intensity projection of a confocal stack 6 µm thick for GFAP, 8 µm thick for Iba1 in the deep cerebellar nuclei was determined by thresholding in Fiji (ImageJ) software. This was done for three *PSMF1*^fl/fl^ Ai14/+ UBC-Cre-ERT2/+ mice per treatment, that is corn oil (control) or tamoxifen injected.

## Data Availability

All data produced in the present study are contained in the manuscript and supplementary material or available upon reasonable request to the authors.

https://curtain.proteo.info/#/6fa3a44a-7708-424b-972b-1f10e585b78e

https://curtain.proteo.info/#/acc6f322-a5b8-498b-ac30-c1143add90c1

https://curtain.proteo.info/#/f96a5ff0-27ab-4392-93ce-b8f667ad6ea7

https://curtain.proteo.info/#/6b5c0cb0-c151-41fe-a44b-0682b1aa8a9f

https://curtain.proteo.info/#/da26e84d-a928-41d2-8f09-0192c7576f46

https://curtain.proteo.info/#/9e2152d9-9c7f-4ce0-8f28-e9074c642474

https://curtain.proteo.info/#/6081d90f-31cf-4d6c-b43c-c4b22774f904

https://curtain.proteo.info/#/95f2c1f9-86f1-4d9f-bede-6acd7a0a338f

## Supplementary Material

- Supplementary Files 1-13

## *PSMF1* Study Group

Francesca Magrinelli, Andrey Y. Abramov, Issam Azmi Alkhawaja, Hadeel Abdollah E Alrukban, Dario R. Alessi, Javeria Raza Alvi, Plamena R. Angelova, Simone Baiardi, Solomiia Bandrivska, Ayşe Nazlı Başak, Peter Bauer, Aida M. Bertoli-Avella, Kailash P. Bhatia, Basar Bilgiç, Vincenzo Bonifati, Alexis Brice, Guido J. Breedveld, Rebecca Buchert, Francesco Cavallieri, Brian Hon-Yin Chung, Martin Man-Chun Chui, Thomas Courtin, Alessio Di Fonzo, Stephanie Efthymiou, Bülent Elibol, Noemi Esteras, Sze-Shing Fan, Christine Fauth, Tawfiq Froukh, Tuğçe Gül-Demirkale, John A. Hardy, Tobias B. Haack, Haşmet A. Hanağası, Tova Hershkovitz, Henry Houlden, Sana Iftikhar, Matthew Jaconelli, James E.C. Jepson, Güneş Kiziltan, Monica M. Kurtis, Anna Ka-Yee Kwong, Suzanne Lesage, Patrick A. Lewis, Pawel Lis, Simon A. Lowe, Angela Mammana, Reza Maroofian, Edoardo Monfrini, David Murphy, Earny Wei-Sen Ng, Raja Nirujogi, José A. Obeso, Benjamin O’Callaghan, Piero Parchi, Chloé Pinon, Jose A. Rodriguez, Clarissa Rocca, Ainara Salazar-Villacorta, Bedia Samanci, Mario Santangelo, Annarita Scardamaglia, Ulrich A. Schatz, Mariasavina Severino, Junko Shimazu, Linda Sofan, Karen Stals, Robert Steinfeld, Hermann Steller, Tipu Sultan, Marina Svetel, Christelle Tesson, Hülya Tireli, Vinod Varghese, Barbara Vona, Marina Volin, Karin Weiss, Gül Yalçın-Cakmakli, Gülbün A. Yüksel, Michael Zech, Thomas Zöggeler.

## Acknowledgments

The authors are indebted to the patients participating in this study as well as their families. For their kind support to this study, we are most grateful to Dr Andrew B. Singleton (National Institutes of Health, Bethesda, USA), Prof. Manju Kurian and Dr Zane Jaunmuktane (University College London, London, United Kingdom), Prof. Olaf Riess and Dr German Demidov (University of Tübingen, Tübingen, Germany), Dr Julia Baptista (Exeter Genomics Laboratory, Exeter, United Kingdom), Dr Erin Torti (GeneDx), Dr Jill Anne Mokry (Baylor Genetics), Prof. Stephan Züchner (University of Miami, Miami, USA), Dr Khadijah Bakur and Dr Fowzan Alkuraya (Lifera Omics), Dr Rami Abou Jamra (University of Leipzig, Leipzig, Germany), Dr Nikola Kresojević (University Clinical Center of Serbia, Belgrade, Serbia), Dr Michele Matarazzo (HM CINAC, Madrid, Spain), Prof. Naci Karaağaç (Istanbul University, Istanbul, Turkey), Dr Bernd Wilken (Klinikum Kassel, Kassel, Germany), Dr Laura Mahoney Sanchez (The Francis Crick Institute, London, United Kingdom), Prof. Christine Klein, Dr Joanne Trinh and Dr Manu Sharma (University of Lübeck, Lübeck, Germany). Cr.Te., Th.Co., Cl.Pi, Ma.Ca, Ba.A.Ha., Gu.Co., Su.Le. and Al.Br. thank the DNA and cell bank of the ICM, Sorbonne Université, Paris, France, for sample preparation and acknowledge that part of this work was carried out on the iGenSeq and DAC core facilities of the ICM. Jo.Ro., Ju.Sh., Ma.Vo. and He.St. are grateful to Beily Durbin for her logistical support to the study. Ed.Mo. Fr.Ca., Ma.Sa. and Al.DF. thank Dr Giulia Di Rauso and Dr Valentina Fioravanti (Neurology Unit, Azienda USL-IRCCS of Reggio Emilia, Reggio Emilia, Italy), and to Dr Stefano Amidei and Dr Giombattista Sallemi (Department of Neurology, Carpi Hospital, Modena, Italy). Tu.Gü. and Ay.N.Ba. gratefully acknowledge Suna and İnan Kıraç Foundation and the use of the services and facilities of the Koç University Research Center for Translational Medicine (KUTTAM), Istanbul, Turkey, funded by the Presidency of Turkey, Head of Strategy and Budget. Ni.Ba. thanks Dr Gordana Mustač (Zadar General Hospital, Zadar, Croatia). Ch.Fa. and To.B.Ha. are members of the European Reference Network on Rare Congenital Malformations and Rare Intellectual Disability ERN-ITHACA [EU Framework Partnership Agreement ID: 3HP-HP-FPA ERN-01-2016/739516]. Mi.Ze. is a member of the Medical and Scientific Advisory Council of the Dystonia Medical Research Foundation and a member of the Governance Council of the International Cerebral Palsy Genomics Consortium. He.Ho. thanks Prof. Vincenzo Salpietro (University of L’Aquila, L’Aquila, Italy). We used data from the UK Biobank Resource under Application Number 33601. This study used the high-performance computational capabilities of the Biowulf Linux cluster at the National Institutes of Health (http://hpc.nih.gov). Data used in the preparation of this article were obtained from the AMP-PD Knowledge Platform. For up-to-date information on the study, visit https://www.amp-pd.org. AMP-PD—a public–private partnership—is managed by the FNIH and funded by Celgene, GSK, the Michael J. Fox Foundation for Parkinson’s Research, the National Institute of Neurological Disorders and Stroke, Pfizer, and Verily. We would like to thank AMP-PD for the publicly available GS data, including cohorts from the Fox Investigation for New Discovery of Biomarkers (BioFIND), the Parkinson’s Progression Markers Initiative (PPMI), and the Parkinson’s Disease Biomarkers Program (PDBP). The Parkinson’s Disease Biomarker Program (PDBP) consortium is supported by the National Institute of Neurological Disorders and Stroke (NINDS) at the National Institutes of Health. A full list of PDBP investigators can be found at https://pdbp.ninds.nih.gov/policy. Harvard Biomarker Study (HBS) is a collaboration of HBS investigators (full list of HBS investigators found at https://www.bwhparkinsoncenter.org/biobank) and funded through philanthropy and NIH and Non-NIH funding sources. The HBS Investigators have not participated in reviewing the data analysis or content of the manuscript. This research was made possible through access to data in the National Genomic Research Library, which is managed by Genomics England Limited (a wholly owned company of the Department of Health and Social Care). The National Genomic Research Library holds data provided by patients and collected by the NHS as part of their care and data collected as part of their participation in research. The National Genomic Research Library is funded by the National Institute for Health Research and NHS England. The Wellcome Trust, Cancer Research UK and the Medical Research Council have also funded research infrastructure. We gratefully acknowledge *All of Us* participants for their contributions, without whom part of this research would not have been possible. We also thank the National Institutes of Health’s *All of Us* Research Program for making available the Public Data Browser (Genomic Variants) screened in this study. The data used for the analysis of PSMF1 transcripts described in this manuscript were obtained from the GTEx Portal (GTEx Transcript Browser) on 18 July 2025.

