## Supplementary material for "Variants in the proteasome regulator PSMF1 cause a phenotypic spectrum from early-onset Parkinson’s disease to perinatal lethality and disrupt mitochondrial function": Table 1

**Table 1. Simplified phenotypic features and genotype of 24 subjects with *PSMF1*-related disorder belonging to 17 families**

| **Pedigree** | **Pedigree A** | **Pedigree B** | | **Pedigree C** | **Pedigree D** | **Pedigree E** | **Pedigree F** | **Pedigree G** | **Pedigree H** | **Pedigree I** |
|---|---|---|---|---|---|---|---|---|---|---|
| **Individual** | **A-II-2** | **B-II-3** | **B-II-4** | **C-II-3** | **D-II-2** | **E-II-4** | **F-II-3** | **G-II-3** | **H-II-1** | **I-II-3** |
| **Sex/Current age** | M/25-30 yrs | F/35-40 yrs | M/Deceased (35-40 yrs) | M/(35-40 yrs) | M/Deceased (40-45 yrs) | F/Deceased (65-70 yrs) | M/30-35 yrs | F/35-40 yrs | F/30-35 yrs | M/20-25 yrs |
| **Parental consanguinity** | No | Yes | Yes | No | Yes | No | Yes | Yes | Yes | No |
| **Family history** | Unremarkable | One sibling affected | One sibling affected | Unremarkable | PD; deaths in childhood | Unremarkable | Unremarkable | Unremarkable | Unremarkable | Unremarkable |
| **Core phenotype** | Parkinsonism | Parkinsonism | Parkinsonism | Parkinsonism | Parkinsonism | Parkinsonism | Parkinsonism | Parkinsonism | Parkinsonism | Parkinsonism |
| **Age at onset** | 20-25 yrs | 15-20 yrs | 15-20 yrs | 25-30 yrs | 25-30 yrs | 45-50 yrs | 15-20 yrs | 15-20 yrs | 0-5 yrs | 0-5 yrs |
| **Symptom at onset** | Bradykinesia, dysphagia, anxiety | Right hand tremor | Right hand tremor | Right upper limb rigidity, right hand tremor and reduced dexterity | Micrographia | Left upper limb tremor | Hand tremor, left foot dragging, anxiety | Bradykinesia, right hand tremor, right foot dystonia | Dystonic head tremor | Clumsiness, frequent falls without fall reflex |
| **Neurological manifestations** | | | | | | | | | | |
| **Motor features** | Parkinsonism (bradykinesia, rigidity, postural instability).<br>Eye movement abnormalities.<br>Dysarthria.<br>Gait festination.<br>Balance difficulty.<br>Pyramidal signs (hyperreflexia, Babinski sign, ankle clonus).<br>Mild cerebellar signs. | Parkinsonism (bradykinesia, rigidity, rest tremor, postural instability).<br>Dysarthria.<br>Freezing of gait.<br>Balance difficulty. | Parkinsonism (bradykinesia, rigidity, rest tremor, postural instability).<br>Blepharospasm.<br>Dysarthria.<br>Postural and kinetic tremor.<br>Freezing of gait to tiptoe walking.<br>Pyramidal signs (hyperreflexia). | Parkinsonism (bradykinesia, rigidity, rest tremor).<br>Postural tremor. | Parkinsonism (bradykinesia, rigidity, postural instability).<br>Blepharospasm.<br>Pyramidal signs (hyperreflexia)<br>Freezing of gait<br>Falls.<br>Tiptoe walking. | Parkinsonism (bradykinesia, rigidity, rest tremor, postural instability).<br>Restless leg syndrome.<br>Freezing of gait. | Parkinsonism.<br>Postural tremor.<br>Pyramidal signs (hyperreflexia, ankle clonus). | Parkinsonism (bradykinesia, rigidity, rest tremor).<br>Postural and kinetic tremor.<br>Falls.<br>Mild pyramidal signs (hyperreflexia of the patellar tendon reflex) | Tremulous segmental dystonia.<br>Parkinsonism (bradykinesia, rigidity, rest tremor) | Parkinsonism (bradykinesia, rigidity, rest tremor).<br>Action tremor.<br>Blepharospasm.<br>Hyperreflexia.<br>Myoclonus.<br>Frequent episodes of freezing.<br>Lower limb muscle hypotrophy.<br>Pes cavus.<br>Ankle joint contractures. |
| **Response to treatment** | Mild-to-moderate response to LD.<br>Early-onset LD-induced dyskinesia. | Mild-to-moderate response to LD.<br>Early-onset LD-induced dyskinesia. | No response to LD.<br>Mild response to DA.<br>Early-onset dyskinesia. | Mild response to DA.<br>Good response to recent LD initiation. | Initial good response to LD.<br>Severe motor fluctuations and LD-induced dyskinesia.<br>Initial good response to DBS. | Initial good response to LD.<br>Severe motor fluctuations and LD-induced dyskinesia.<br>Initial good response to DBS. | Moderate response to LD.<br>LD-induced dyskinesia. | Response to LD only in the early phase.<br>Mild response to DA. | Poor response to LD. | Initial good response to LD. |
| **Brain MRI (age)** | Mild cerebral atrophy, faint T2 hyperintensity in the posterior periventricular WM (20-25 yrs) | Normal. | Mild atrophy of the inferior cerebellar vermis. | Small hemosiderin deposit adjacent to the frontal horn of the right lateral ventricle. | Normal. | Normal. | Mild-to-moderate cerebral atrophy, thin CC, small AC, small midbrain, mild atrophy of superior cerebellar vermis (25-30 yrs) | Normal (25-30 yrs) | Mild cerebellar cortical atrophy. | Retrocerebellar cyst. |
| **Other instrumental investigations** | DaTscan: abnormal.<br>Myocardial MIBG: normal.<br>DEXA: osteopenia. | EEG: normal.<br>EMG/NCS: normal. | EEG: normal.<br>EMG/NCS: normal.<br>Muscle biopsy: normal. | DaTscan: abnormal.<br>Myocardial MIBG: normal. | F-DOPA-PET: abnormal.<br>FDG-PET: temporal hypometabolism. | - | US: no organomegaly.<br>Ophthalmological assessment: normal.<br>Normal CK, lactate, pyruvate, copper, caeruloplasmin and 24-hour urinary copper excretion. | Brain perfusion SPECT: reduced perfusion in the left thalamus. | EEG/Video-EEG: normal. | DaTscan: abnormal.<br>Low testosterone level.<br>EEG: diffuse theta activity.<br>VEP: prolonged latency.<br>Brain MRS: normal.<br>Muscle MRI: atrophy of gluteal muscles, with increased muscle fat.<br>Spine X-ray: scoliosis.<br>DEXA: severe osteopenia. |
| **Genotype *PSMF1*** (NM_006814.5) | C-HET<br>c.724C>G p.(Arg242Gly)<br>c.282+2T>A | HOM<br>c.724C>G p.(Arg242Gly) | HOM<br>c.724C>G p.(Arg242Gly) | HOM<br>c.724C>G p.(Arg242Gly) | HOM<br>c.724C>T p.(Arg242Cys) | HOM<br>c.725G>A p.(Arg242His) | HOM<br>c.157C>A p.(Leu53Met) | HOM<br>c.157C>A p.(Leu53Met) | HOM<br>c.682C>G p.(Leu228Val) | C-HET<br>c.724C>G p.(Arg242Gly)<br>c.691C>T p.(Arg231*) |

**Table 1 (continued). Simplified phenotypic features and genotype of 24 subjects with *PSMF1*-related disorder belonging to 17 families**

| **Pedigree** | | **Pedigree J** | | **Pedigree K** | **Pedigree L** | | | **Pedigree M** | | | **Pedigree N** |
|---|---|---|---|---|---|---|---|---|---|---|---|
| **Individual** | | **J-II-1** | **J-II-3** | **K-II-3** | **L-II-1** | **L-II-3** | **L-II-4** | **M-II-1** | **M-II-3** | **M-II-6** | **N-II-1** |
| **Sex/Current age** | | F/Deceased (15-20 yrs) | M/15-20 yrs | M/20-25 yrs | F/10-15 yrs | F/5-10 yrs | F/5-10 yrs | F/Deceased (15-20 yrs) | F/Deceased (15-20 yrs) | M/Deceased (15-20 yrs) | F/12-24 mos |
| **Parental consanguinity** | | No | No | Yes | Yes | Yes | Yes | Yes | Yes | Yes | Yes |
| **Family history** | | One sibling affected | One sibling affected | Unremarkable | Two siblings affected | Two siblings affected | Two siblings affected | Two siblings affected | Two siblings affected | Two siblings affected | Unremarkable |
| **Core phenotype** | | Parkinsonian-pyramidal syndrome | Spastic-ataxia syndrome | DD with CC agenesis | DD with CC hypoplasia | DD with CC hypoplasia | DD with CC hypoplasia | Parkinsonian-pyramidal syndrome with CC hypoplasia | Parkinsonian-pyramidal syndrome with CC hypoplasia | Parkinsonian-pyramidal syndrome with CC hypoplasia | Arthrogryposis with CC hypoplasia |
| **Age at onset** | | 0-5 yrs | 0-5 yrs | Early childhood | Early childhood | Early childhood | Early childhood | 10-15 yrs | 10-15 yrs | Infancy | Prenatal |
| **Symptom at onset** | | Limb spasticity | Limb spasticity | GDD | GDD | SLDD | GDD | Memory and attentional deficits | Memory and attentional deficits | Epilepsy | N/A |
| **Neurological manifestations** | | | | | | | | | | | |
| | **Motor features** | Parkinsonism.<br>Pyramidal signs (spasticity, hyperreflexia)<br>Strabismus<br>Ataxia | MDD<br>Pyramidal signs. (spasticity, hyperreflexia)<br>Ataxia | Global hypokinesia.<br>Clumsiness.<br>Mild hypotonia. | Hypokinesia.<br>Broad-based gait.<br>Talipes. | Hypokinesia | Hypokinesia.<br>Talipes. | Hypokinesia.<br>Pyramidal tract signs. | Hypokinesia.<br>Pyramidal tract signs. | Hypokinesia.<br>Pyramidal tract signs. | Arthrogryposis.<br>Limb spasticity.<br>Increased jitteriness.<br>GDD |
| | **Response to treatment** | N/A | N/A | N/A | Partial response to LEV. Good response to CBZ and VPA | N/A | N/A | N/A | Partial response to CBZ and VPA | Unspecified response to CBZ | N/A |
| **Brain MRI (age)** | | Atrophy of cerebral hemispheres, basal ganglia, thalami, brainstem and cerebellum, with mineralization of the lentiform nuclei. | Mild T2 signal alterations in the anterior midbrain bilaterally. | CC agenesis, malformation of the left cerebellar hemisphere, cerebellar vermis hypoplasia.<br>Arachnoid cyst. | Moderate cerebral atrophy, thin CC (10-15 yrs) | N/A | N/A | Progressive mild-to-moderate cerebral atrophy, CC hypoplasia, small AC, faint periventricular T2 signal alterations, mild superior cerebellar vermis atrophy (10-15 yrs) | Moderate cerebral atrophy, CC hypoplasia, small AC, faint periventricular T2 signal alterations, mild superior cerebellar vermis atrophy (10-15 yrs) | CC hypoplasia, small ACC, faint posterior periventricular T2 signal (0-5 yrs) | CC hypoplasia.<br>Absent myelination within the posterior portion of the internal capsule. Multiple foci of T2 hyperintensity within the lentiform nuclei (infancy). |
| **Other investigations** | | DaTscan: abnormal. | Muscle biopsy and enzymology: normal. | Prenatal US: hydrocephalus.<br>DEXA: osteopenia (on bisphosphonates). | EEG: multifocal epileptiform discharges | N/A | N/A | Sleep EEG (age 10-15 yrs): general disorganization of brain electrical activity without epileptiform discharges. | EEG: abnormal, with focus | EEG: initially normal, then showing epileptic activity mainly in the temporo-parietal regions. | EEG: normal.<br>Postnatal abdominal US: normal. |
| **Genotype *PSMF1*** (NM 006814.5) | | HOM<br>c.282+5G>A | HOM<br>c.282+5G>A | HOM<br>c.605+1G>A | HOM<br>c.764+5G>A | HOM<br>c.764+5G>A | HOM<br>c.764+5G>A | N/A | HOM<br>c.764+5G>A | HOM<br>c.764+5G>A | HOM<br>c.691C>T p.(Arg231*) |

**Table 1 (continued). Simplified phenotypic features and genotype of 24 subjects with *PSMF1*-related disorder belonging to 17 families**

| **Pedigree** | **Pedigree O** | | **Pedigree P** | **Pedigree Q** |
|---|---|---|---|---|
| **Individual** | **O-II-4** | **O-II-5** | **P-II-3** | **Q-II-2** |
| **Sex/Current age** | M/Deceased (0-1 yr) | F/Deceased (0-1 yr) | M/Deceased (0-1 yr) | M/Feticide (GA third trimester) |
| **Parental consanguinity** | Yes | Yes | No | No |
| **Family history** | One pregnancy loss; one sibling affected. | One pregnancy loss; one sibling affected. | One miscarriage | Unremarkable |
| **Core phenotype** | Arthrogryposis | Arthrogryposis with CC agenesis | Arthrogryposis | Arthrogryposis with CC agenesis |
| **Age at onset** | Prenatal | Prenatal | Prenatal | Prenatal |
| **Symptom at onset** | N/A* | N/A | N/A | N/A |
| **Neurological manifestations** | | | | |
| **Motor features** | N/A* | Myoclonus.<br>Arthrogryposis.<br>Limb spasticity.<br>Truncal hypotonia.<br>GDD. | Arthrogryposis. | Arthrogryposis |
| **Response to treatment** | N/A* | Poor response to ASM. | N/A | N/A |
| **Brain MRI (age)** | N/A* | CC hypoplasia. Hypomyelination. Progressive cerebral atrophy. Subdural hygromas (infancy). | Cavum septum pellucidum and cavum vergae | N/A |
| **Other investigations** | N/A* | CTG: pathological.<br>EEG: intermittent epileptic activity.<br>Prolonged ECG: intermittent bradycardia.<br>Postnatal echocardiography: ASD type II with left-right shunt.<br>Spine MRI: low-lying conus medullaris. | Prenatal US: polyhydramnios, clenched fists, clubfeet.<br>Neonatal head and spine US: reduced gyration, low-lying conus medullaris/tethered cord.<br>EEG: pathological pattern.<br>Postnatal echocardiography: unremarkable.<br>Postnatal abdominal US: left hydronephrosis and hydroureter. | Prenatal US (GA 20-25 wks): normal.<br>Prenatal US (GA 30-35 wks): polyhydramnios, suspected CC agenesis, small stomach.<br>Prenatal US (GA 35-40 wks): fetal growth restriction, abnormal fetal movements, CC agenesis, intracerebral cyst, arthrogryposis, thickened myocardium, small stomach, left hydronephrosis and hydroureter.<br>QF-PCR: negative.<br>CGH: negative. |
| **Genotype *PSMF1*** (NM_006814.5) | N/A* | HOM<br>c.1A>T | C-HET<br>c.101del p.(Gly34Valfs*47)<br>c.129+2T>C | C-HET<br>c.365+2T>C<br>c.1A>T |

**Legend:** AC = anterior commissure; ASD = atrial septal defect; ASM = anti-seizure medication(s); CBZ = carbamazepine; C-HET = compound heterozygote; CC = corpus callosum; CGH = comparative genomic hybridization; CK = creatine kinase; CTG = cardiotocography; DBS = deep brain stimulation; DD = developmental delay; DEXA = bone density scan; ECG = electrocardiogram; EEG = electroencephalogram; F = female; GA = gestational age; GDD = global developmental delay; HOM = homozygote; LEV = levetiracetam; M = male; MDD = motor developmental delay; MIBG = $^{123}$I-metaiodobenzylguanidine scintigraphy; mo(s) = month(s); MRI = magnetic resonance imaging; MRS = magnetic resonance spectroscopy; N/A = not applicable or not available; PD = Parkinson's disease; QF-PCR = quantitative fluorescence polymerase chain reaction; SLDD = speech and language developmental delay; SPECT = single-photon emission computed tomography; US = ultrasound; VEP = visual evoked potentials; VPA = valproic acid; wk(s) = week(s); yr(s) = year(s). *Clinical details not available (O-II-4 reported with a phenotype similar to his sister O-II-5). **Detailed clinical information is available upon request to the corresponding author.**
