## Supplementary Files for "Variants in the proteasome regulator PSMF1 cause a phenotypic spectrum from early-onset Parkinson’s disease to perinatal lethality and disrupt mitochondrial function"

**Supplementary File 1. Frequencies, pathogenicity predictions and computational analysis of 14 *PSMF1* variants detected in the study cohort**

### *a. PSMF1 non-splice variants*

| | V1 | V3 | V4 | V5 | V6 | V7 | V11 | V12 |
|---|---|---|---|---|---|---|---|---|
| **Variant *PSMF1* (GRCh38/hg38) [NM_006814.5]** | chr20:g.1164436:C>G<br>c.724C>G<br>p.(Arg242Gly) | chr20:g.1164436:C>T<br>c.724C>T<br>p.(Arg242Cys) | chr20:g.1164437:G>A<br>c.725G>A<br>p.(Arg242His) | chr20:g.1125525:C>A<br>c.157C>A<br>p.(Leu53Met) | chr20:g.1164394:C>G<br>c.682C>G<br>p.(Leu228Val) | chr20:g.1164403:C>T<br>c.691C>T<br>p.(Arg231*) | chr20:g.1118774:A>T<br>c.1A>T<br>p.(0) | chr20:g.1118872:CG>C<br>c.101del<br>p.(Gly34Valfs*47) |
| **gnomAD v2.1.1/v3.1.2/v4.1.0 (het, hom, allele frequency)** | v2.1.1: 7 het, 0 hom, *f* = 0.00002787<br>v3.1.2: not found<br>v4.1.0: 5 het, 0 hom, *f* = 0.000003098 | v2.1.1: 2 het, 0 hom, *f* = 0.000007964<br>v3.1.2: 3 het, 0 hom, *f* = 0.00001972<br>v4.1.0: 8 het, 0 hom, *f* = 0.000004956 | v2.1.1: 427 het, 0 hom, *f* = 0.001511<br>v3.1.2: 224 het, 0 hom, *f* = 0.001472<br>v4.1.0: 3454 het, 4 hom, *f* = 0.002140 | v2.1.1: not found<br>v3.1.2: not found<br>v4.1.0: not found | v2.1.1: not found<br>v3.1.2: not found<br>v4.1.0: not found | v2.1.1: not found<br>v3.1.2: not found<br>v4.1.0: 4 het, 0 hom, *f* = 0.000002478 | v2.1.1: 1 het, 0 hom, *f* = 0.000004074<br>v3.1.2: not found<br>v4.1.0: 1 het, 0 hom, *f* = $6.215e^{-7}$ | v2.1.1: not found<br>v3.1.2: 1 het, 0 hom, *f* = 0.000006568<br>v4.1.0: 4 het, 0 hom, *f* = 0.000002478 |
| **100,000 Genomes Project (aggV2, 78,195 germline gVCFs)** | Not found | Not found | 407 het, 1 hom, *f* = 0.0026152567 | Not found | Not found | Not found | Not found | Not found |
| **UK Biobank (accessed on 3 June 2025)** | Not found | 2 het, 0 hom, *f* = $2.72e^{-06}$<br>(total allele # 735,924) | 2016 het, 2 hom, *f* = 0.002739<br>(total allele # 735,924) | Not found | Not found | 2 het, 0 hom, *f* = $2.72e^{-06}$<br>(total allele # 735,888) | Not found | Not found |
| ***All of Us* Public Data Browser (accessed on 14 July 2025)** | 8 het, 0 hom, *f* = 0.00001<br>(total allele # 829,632) | 1 het, 0 hom, *f* = 0.000001<br>(total allele # 829,632) | 1462 het, 0 hom, *f* = 0.001762<br>(total allele # 829,624) | Not found | Not found | 14 het, 0 hom, *f* = 0.000017<br>(total allele # 829,628) | Not found | 4 het, 0 hom, *f* = 0.000005<br>(total allele # 829,648) |
| **Queen Square Genomics (35,574 samples)** | 3 het*, 0 hom, *f* = $4.25e^{-05}$<br>(total allele # 70,622) | 0 het, 1 hom**, *f* = $2.83e^{-05}$<br>(total allele # 70,622) | 81 het, 0 hom, *f* = 0.001147<br>(total allele # 70,640) | Not found | Not found | 3 het***, 0 hom, *f* = $4.30e^{-05}$<br>(total allele # 69,746) | Not found | Not found |
| **CADD v1.7** | 28.6 | 32 | 32 | 22.8 | 22.6 | 37 | 24.7 | N/A |
| **PolyPhen2 HVAR** | Possibly damaging<br>(score: 0.997) | Possibly damaging<br>(score: 0.999) | Possibly damaging<br>(0.858) | Possibly damaging<br>(score: 0.967) | Possibly damaging<br>(score: 0.967) | N/A | Possibly damaging<br>(score: 0.970) | N/A |
| **SIFT4G** | Damaging<br>(0, 0, 0, 0) | Damaging (0, 0, 0, 0) | Damaging (0, 0, 0, 0) | Tolerated<br>(0.226, 0.226, 0.226, 0.226) | Tolerated (0.228, 0.481, 0.228) | N/A | Tolerated/ Damaging (0.278, 0, 0.259, 0.278) | N/A |
| **PROVEAN** | Damaging<br>(-6.3, -6.3, -6.3, -3.21) | Damaging (-7.21, -7.21, -7.21, -4.01) | Damaging, Damaging, Damaging, Neutral (-4.5, -4.5, -4.5, -2.22) | Neutral (-1.15, -1.25, -1.05, -1.15) | Neutral (-0.33, -0.36, -0.33) | N/A | Neutral (-1.18, -1.14, -1.24, -1.18) | N/A |
| **MutationTaster** | Disease causing (prob: 0.999978910569413) | Disease causing (prob: 0.999999926286941) | Disease causing (prob: 0.999993833789465) | Disease causing (prob: 0.999846573719514) | Disease causing (prob: 0.988685291987796) | Disease causing (prob: 1) | Disease causing (prob: 1) | Disease causing (prob: 1) |
| **HSF** | No significant impact on splicing signals | Alteration of auxiliary sequences: Significant alteration of ESE / ESS motifs ratio (-5) | No significant impact on splicing signals | No significant impact on splicing signals | No significant impact on splicing signals | Alteration of auxiliary sequences: Significant alteration of ESE / ESS motifs ratio (-3) | Alteration of auxiliary sequences: Significant alteration of ESE / ESS motifs ratio (-6) | Alteration of auxiliary sequences: Significant alteration of ESE / ESS motifs ratio (-2)<br>New Donor splice site: Activation of a cryptic Donor site. Potential alteration of splicing (HSF) |
| **GERP (Franklin Genoox)** | 4.13 | 4.13 | 5.11 | 4.93 | 6.07 | 5.13 | 4.61 | N/A |
| **GERP (UCSC)** | 4.13 | 4.13 | 5.11 | 4.93 | 6.07 | 5.13 | 4.61 | 4.26 |

#### *b.* *PSMF1 splice variants*

| | V2 | V8 | V9 | V10 | V13 | V14 |
|---|---|---|---|---|---|---|
| **Variant *PSMF1* (GRCh38/hg38) [NM_006814.5]** | chr20:g.1125652:T>A<br>c.282+2T>A | chr20:g.1125655:G>A<br>c.282+5G>A | chr20:g.1163184:G>A<br>c.605+1G>A | chr20:g.1164481:G>A<br>c.764+5G>A | chr20:g.1118904:T>C<br>c.129+2T>C | chr20:g.1127510:T>C<br>c.365+2T>C |
| **gnomAD v2.1.1/v3.1.2/v4.1.0 (het, hom, allele frequency)** | v2.1.1: 1 het, 0 hom, $f$ = 0.000004052<br>v3.1.2: not found<br>v4.1.0: not found | v2.1.1: 4 het, 0 hom, $f$ = 0.00001448<br>v3.1.2: 1 het, 0 hom, $f$ = 0.000006571<br>v4.1.0: 9 het, 0 hom, $f$ = 0.000005597 | v2.1.1: 5 het, 0 hom, $f$ = 0.00001990<br>v3.1.2: 1 het, 0 hom, $f$ = 0.000006570<br>v4.1.0: 23 het, 0 hom, $f$ = 0.00001425 | v2.1.1: 2 het, 0 hom, $f$ = 0.000007979<br>v3.1.2: 2 het, 0 hom, $f$ = 0.00001314<br>v4.1.0: 23 het, 0 hom, $f$ = 0.000008055 | v2.1.1: not found<br>v3.1.2: not found<br>v4.1.0: 8 het, 0 hom, $f$ = 0.000004957 | v2.1.1: not found<br>v3.1.2: not found<br>v4.1.0: 5 het, 0 hom, $f$ = 0.000003167 |
| **100,000 Genomes Project (aggV2, 78,195 germline gVCFs)** | 1 het, 0 hom, $f = 6.39e^{-06}$ | 1 het, 0 hom, $f = 6.39e^{-06}$ | 3 het, 0 hom, $f = 1.92e^{-05}$ | Not found | Not found | 1 het, 0 hom, $f = 6.39e^{-06}$ |
| **UK Biobank (accessed on 3 June 2025)** | Not found | Not found | Not found | 4 het, 0 hom, $f = 5.44e^{-06}$<br>(total allele # 735,916) | 8 het, 0 hom, $f = 1.09e^{-05}$<br>(total allele # 735,902) | 1 het, 0 hom, $f = 1.42e^{-06}$<br>(total allele # 705,032) |
| ***All of Us* Public Data Browser (accessed on 14 July 2025)** | 2 het, 0 hom, $f$ = 0.000002<br>(total allele # 829,594) | 8 het, 0 hom, $f$ = 0.00001<br>(total allele # 829,630) | 2 het, 0 hom, $f$ = 0.000002<br>(total allele # 829,644) | 3 het, 0 hom, $f$ = 0.000004<br>(total allele # 829,612) | 4 het, 0 hom, $f$ = 0.000005<br>(total allele # 829,536) | Not found |
| **Queen Square Genomics exome database (35,574 samples)** | 2 het****, 0 hom, $f = 2.84e^{-05}$<br>(total allele # 70,440) | 1 het, 0 hom, $f = 1.42e^{-05}$<br>(total allele # 70,404) | Not found | 3 het*****, 2 hom******, $f = 9.93e^{-05}$<br>(total allele # 70,478) | Not found | Not found |
| **CADD v1.7** | 36 | 22.4 | 34 | 25 | 29.2 | 26.9 |
| **MutationTaster** | Disease causing (prob: 1) | Disease causing (prob: 1) | Disease causing (prob: 1) | Disease causing (prob: 1) | Disease causing (prob: 1) | Disease causing (prob: 1) |
| **HSF** | Broken WT Donor Site: Alteration of the WT Donor site, most probably affecting splicing (site donor broken) | Broken WT Donor Site: Alteration of the WT Donor site, most probably affecting splicing (site donor broken) | Broken WT Donor Site: Alteration of the WT Donor site, most probably affecting splicing (site donor broken) | Broken WT Donor Site: Alteration of the WT Donor site, most probably affecting splicing (site donor broken) | Broken WT Donor Site: Alteration of the WT Donor site, most probably affecting splicing (site donor broken) | Broken WT Donor Site: Alteration of the WT Donor site, most probably affecting splicing (site donor broken) |
| **SpliceSiteFinder-like [0-100]** | -100% | -13.9% | -100% | -100% | -2.5% | 0% |
| **MaxEntScan [0-12]** | -100% | -65.1% | -100% | -56.5% | -100% | -100% |
| **NNSPLICE [0-1]** | -100% | -100% | -100% | -56.2% | -100% | -100% |
| **GeneSplicer [0-24]** | -100% | -100% | -100% | -58.9% | -100% | -100% |
| **SpliceAI [≥0.2\|0.5\|0.8]** | 0.95 DG (15)<br>1.00 DL (-2) | 0.89 DG (12)<br>0.96 DL (-5) | 0.99 DL (-1) | 0.34 DG (11)<br>0.78 DL (-5) | 0.99 DL (-2) | 0.99 DL (-2) |
| **AbSplice [≥0.01\|0.05\|0.2]** | 0.33 | 0.27 | 0.36 | 0.31 | 0.14 | 0.22 |
| **GERP (Franklin Genoox)** | 4.93 | N/A | 5.36 | N/A | 3.96 | 4.53 |
| **GERP (UCSC)** | 4.93 | 0.225 | 5.36 | 5.95 | 3.96 | 4.53 |

**Legend:** AbSplice = https://absplice.cmm.cit.tum.de/; *All of Us* Public Data Browser = https://databrowser.researchallofus.org/snvsindels; CADD = Combined Annotation Dependent Depletion (https://cadd.gs.washington.edu/snv); ESE = Exonic Splicing Enhancer; ESS = Exonic Splicing Silencer; $f$ = allele frequency; GeneSplicer = https://ccb.jhu.edu/software/genesplicer/; GERP = Genomic Evolutionary Rate Profiling (http://mendel.stanford.edu/SidowLab/downloads/gerp/); GERP (UCSC) = Genomic Evolutionary Rate Profiling by University of California, Santa Cruz (https://genome.ucsc.edu/cgi-bin/hgTrackUi?db=hg19&g=allHg19RS_BW); gnomAD = The Genome Aggregation Database (https://gnomad.broadinstitute.org/); GRCh38/hg38 = Genome Reference Consortium Human Build 38 Organism: Homo sapiens (human); gVCFs = genomic variant call format (VCF); het = alternate allele count; hom = number of homozygotes; HSF = Human Splicing Finder (https://hsf.genomnis.com/home); MaxEntScan = http://hollywood.mit.edu/burgelab/maxent/Xmaxentscan_scoreseq.html; MutationTaster = http://www.mutationtaster.org/; N/A = not applicable/not available; NNSPLICE = https://www.fruitfly.org/seq_tools/splice.html; PolyPhen-2 HVAR: Polymorphism Phenotyping v2, HumVar model (http://genetics.bwh.harvard.edu/pph2/); Prob = probability; PROVEAN: Protein Variation Effect Analyzer (http://provean.jcvi.org/index.php); SIFT 4G: Sorting Intolerant From Tolerant Databases for Genome (https://sift.bii.a-star.edu.sg/sift4g/public//Homo_sapiens/); SpliceAI = https://spliceailookup.broadinstitute.org/; SpliceSiteFinder-like = https://www.interactive-biosoftware.com/; WT = wild type. Allele # = total number of alleles. V# indicates *PSMF1* variants (NM_006814.5) displayed with different font colors (Fig.1a-2a). *Correspond to individuals A-I-1, A-II-2, I-II-3 in this study cohort. **Corresponds to proband C-II-2 in this study cohort. ***One corresponds to proband I-II-3 in this study cohort. ****Correspond to individuals A-I-2 and A-II-2 in this study cohort. *****Two correspond to individuals M-I-1 and M-I-2 in this study cohort. ******Correspond to probands L-II-1 and M-II-6 in this study cohort.

**Supplementary File 2. Autozygosity mapping of probands carrying homozygous *PSMF1* variants and their relatives**

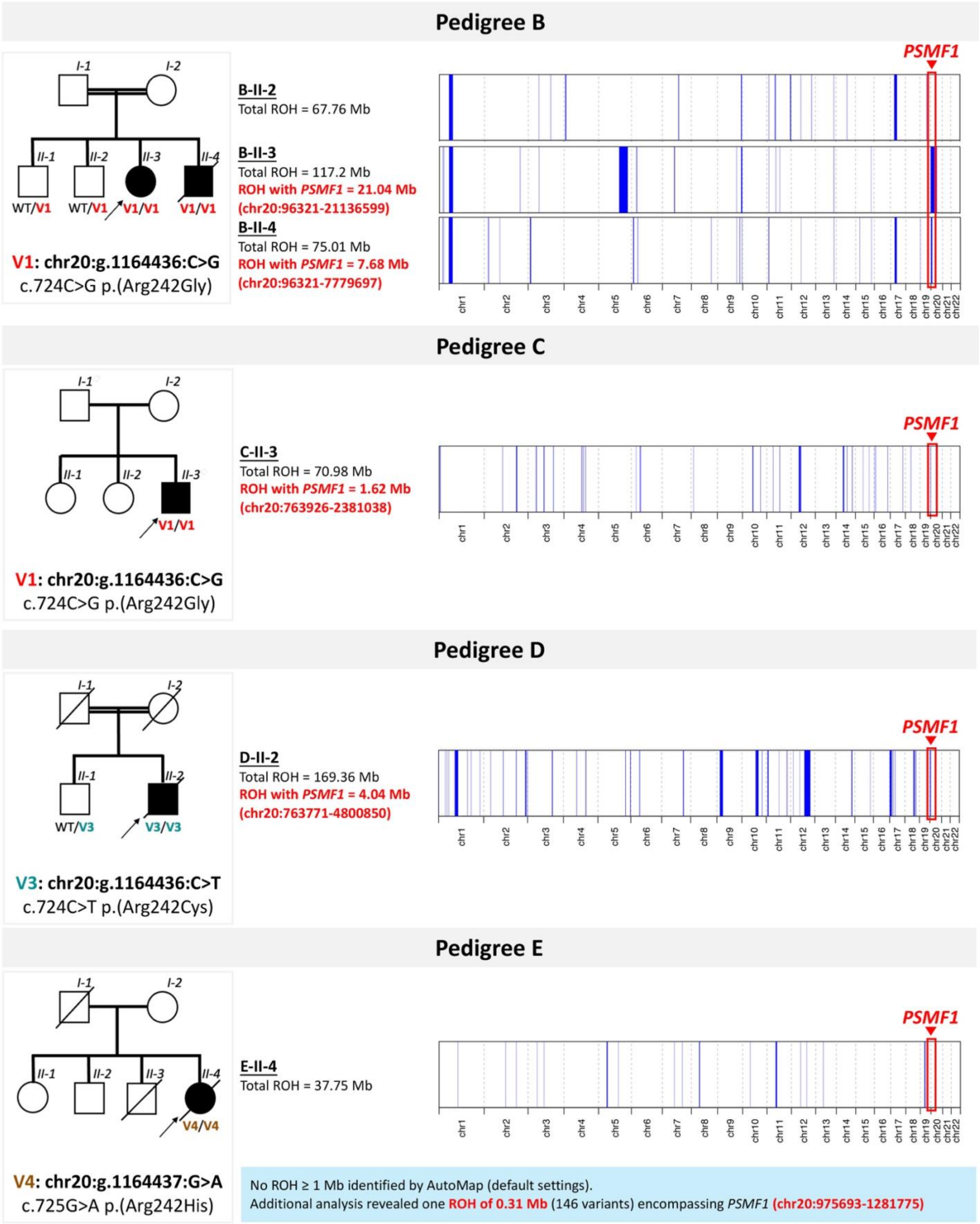

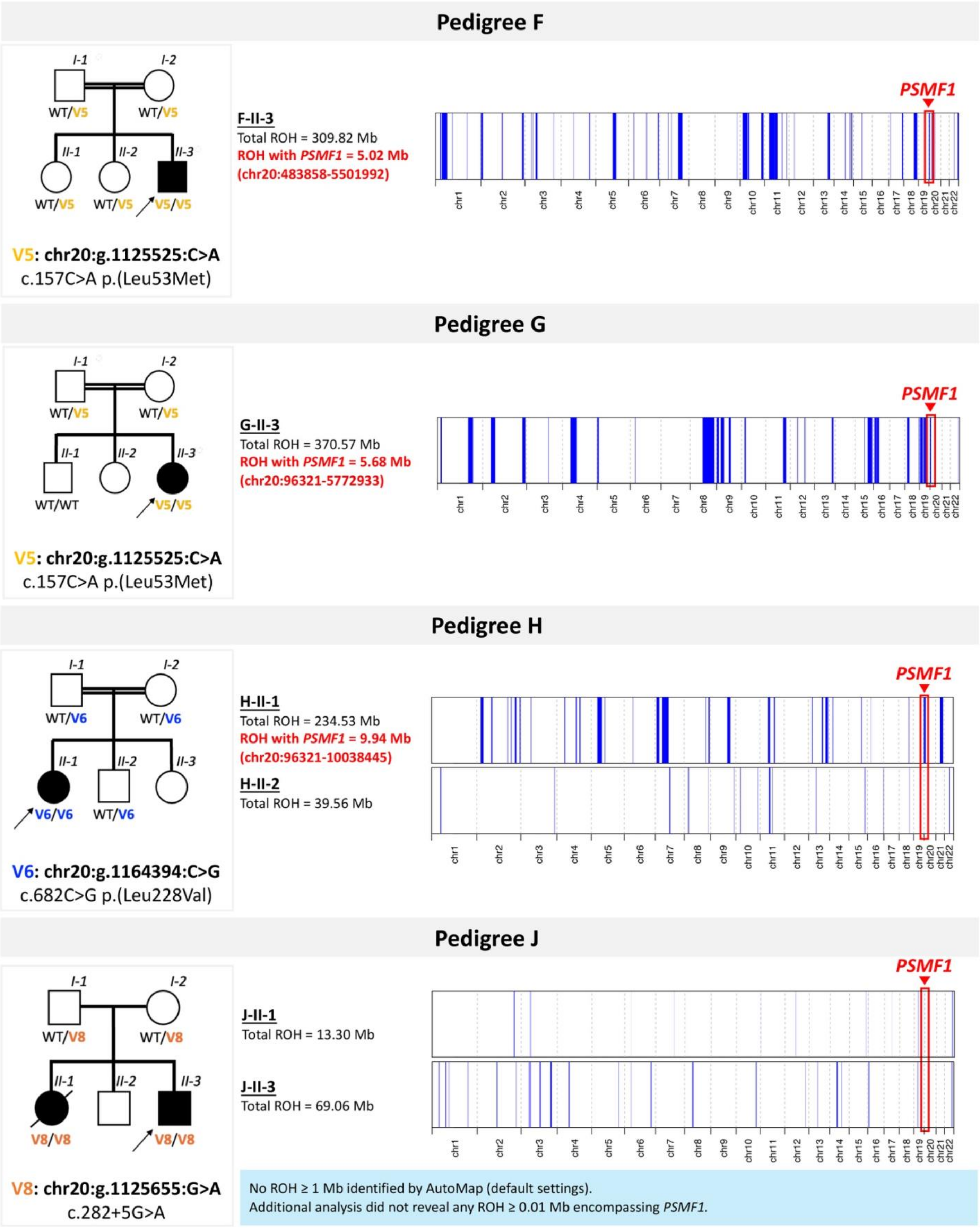

Pedigree F
I-1
I-2
WT/V5
WT/V5
II-1
II-2
II-3
WT/V5
WT/V5
V5/V5
V5: chr20:g.1125525:C>A
c.157C>A p.(Leu53Met)
F-II-3
Total ROH = 309.82 Mb
ROH with PSMF1 = 5.02 Mb
(chr20:483858-5501992)
PSMF1
chr1 chr2 chr3 chr4 chr5 chr6 chr7 chr8 chr9 chr10 chr11 chr12 chr13 chr14 chr15 chr16 chr17 chr18 chr19 chr20 chr21 chr22
Pedigree G
I-1
I-2
WT/V5
WT/V5
II-1
II-2
II-3
WT/WT
V5/V5
V5: chr20:g.1125525:C>A
c.157C>A p.(Leu53Met)
G-II-3
Total ROH = 370.57 Mb
ROH with PSMF1 = 5.68 Mb
(chr20:96321-5772933)
PSMF1
chr1 chr2 chr3 chr4 chr5 chr6 chr7 chr8 chr9 chr10 chr11 chr12 chr13 chr14 chr15 chr16 chr17 chr18 chr19 chr20 chr21 chr22
Pedigree H
I-1
I-2
WT/V6
WT/V6
II-1
II-2
II-3
V6/V6
WT/V6
V6: chr20:g.1164394:C>G
c.682C>G p.(Leu228Val)
H-II-1
Total ROH = 234.53 Mb
ROH with PSMF1 = 9.94 Mb
(chr20:96321-10038445)
H-II-2
Total ROH = 39.56 Mb
PSMF1
chr1 chr2 chr3 chr4 chr5 chr6 chr7 chr8 chr9 chr10 chr11 chr12 chr13 chr14 chr15 chr16 chr17 chr18 chr19 chr20 chr21 chr22
Pedigree J
I-1
I-2
WT/V8
WT/V8
II-1
II-2
II-3
V8/V8
V8/V8
V8: chr20:g.1125655:G>A
c.282+5G>A
J-II-1
Total ROH = 13.30 Mb
J-II-3
Total ROH = 69.06 Mb
PSMF1
chr1 chr2 chr3 chr4 chr5 chr6 chr7 chr8 chr9 chr10 chr11 chr12 chr13 chr14 chr15 chr16 chr17 chr18 chr19 chr20 chr21 chr22
No ROH ≥ 1 Mb identified by AutoMap (default settings).
Additional analysis did not reveal any ROH ≥ 0.01 Mb encompassing PSMF1.

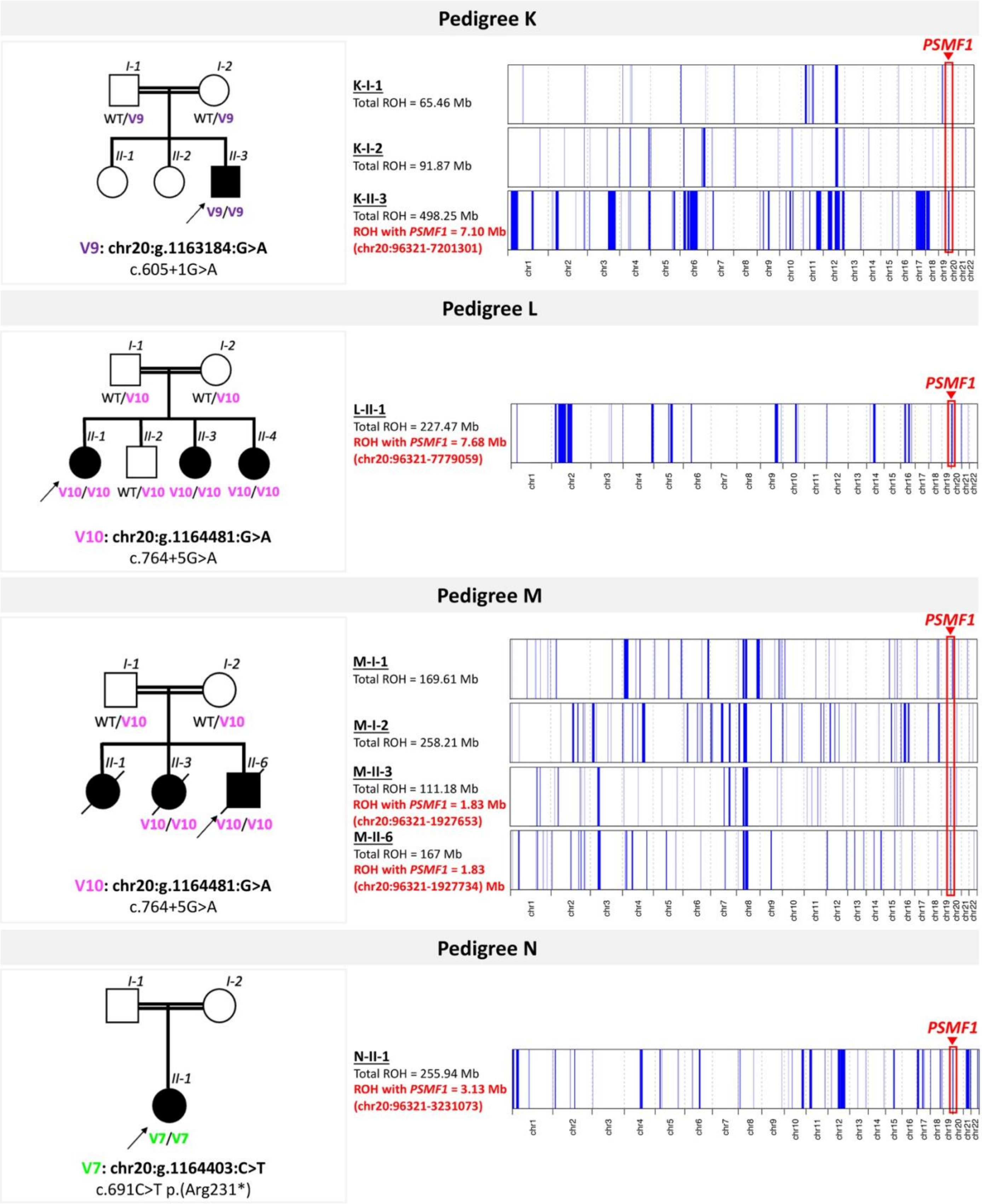
Pedigree K
I-1
I-2
WT/V9
WT/V9
II-1
II-2
II-3
V9/V9
V9: chr20:g.1163184:G>A
c.605+1G>A
PSMF1
K-I-1
Total ROH = 65.46 Mb
K-I-2
Total ROH = 91.87 Mb
K-II-3
Total ROH = 498.25 Mb
ROH with PSMF1 = 7.10 Mb
(chr20:96321-7201301)
Pedigree L
I-1
I-2
WT/V10
WT/V10
II-1
II-2
II-3
II-4
V10/V10
WT/V10
V10/V10
V10/V10
V10: chr20:g.1164481:G>A
c.764+5G>A
PSMF1
L-II-1
Total ROH = 227.47 Mb
ROH with PSMF1 = 7.68 Mb
(chr20:96321-7779059)
Pedigree M
I-1
I-2
WT/V10
WT/V10
II-1
II-3
II-6
V10/V10
V10/V10
V10: chr20:g.1164481:G>A
c.764+5G>A
PSMF1
M-I-1
Total ROH = 169.61 Mb
M-I-2
Total ROH = 258.21 Mb
M-II-3
Total ROH = 111.18 Mb
ROH with PSMF1 = 1.83 Mb
(chr20:96321-1927653)
M-II-6
Total ROH = 167 Mb
ROH with PSMF1 = 1.83
(chr20:96321-1927734) Mb
Pedigree N
I-1
I-2
II-1
V7/V7
V7: chr20:g.1164403:C>T
c.691C>T p.(Arg231*)
PSMF1
N-II-1
Total ROH = 255.94 Mb
ROH with PSMF1 = 3.13 Mb
(chr20:96321-3231073)
chr1
chr2
chr3
chr4
chr5
chr6
chr7
chr8
chr9
chr10
chr11
chr12
chr13
chr14
chr15
chr16
chr17
chr18
chr19
chr20
chr21
chr22

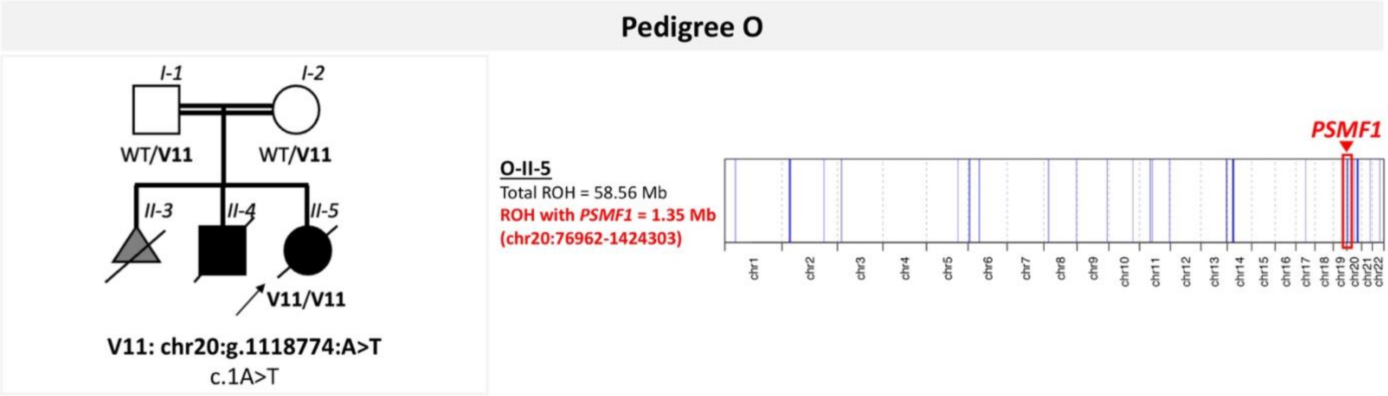


**Legend:** Autozygosity mapping using the tool AutoMap[4] identified regions of homozygosity larger than ≥1 Mb in all probands carrying homozygous *PSMF1* variants and their family members with raw data from ES or GS (VCF or FASTQ files) available. Simplified pedigrees and genomic coordinates (GRCh38/hg38) of *PSMF1* variants segregating in the families are displayed on the left as a reference. V# indicates *PSMF1* variants referenced to *PSMF1* transcript NM_006814.5 and displayed with different font colors (Fig.1a-2a). Regions of homozygosity identified in single individuals are shown in blue, with their total genomic extension across autosomes (in Mb) reported in the middle part of each panel. The genomic region encompassing *PSMF1* is marked with a red box and arrowhead. If a region of homozygosity including *PSMF1* was detected, its size (in Mb) and genomic coordinates are displayed in red in single individuals. When AutoMap (default parameters) did not identify a region of homozygosity encompassing *PSMF1* (Pedigrees E-J), results of additional analysis for regions of homozygosity ≥ 0.01 Mb are provided in a light blue box. GRCh38/hg38 = Genome Reference Consortium Human Build 38 Organism: Homo sapiens (human); Mb = megabase(s); ROH = region(s) of homozygosity.

**Supplementary File 3. Haplotype analyses of three *PSMF1* variants detected in the homozygous state in affected individuals from multiple families**

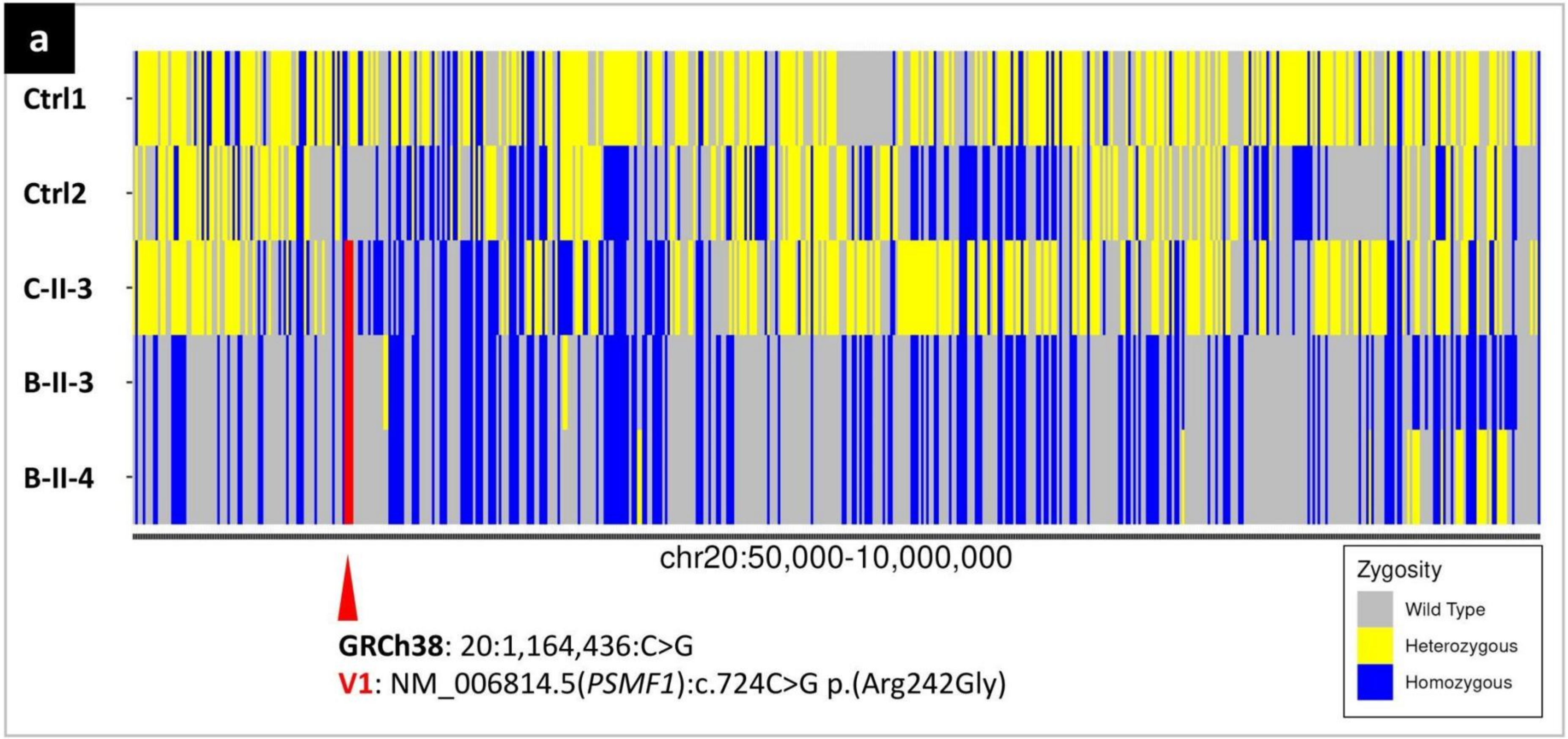

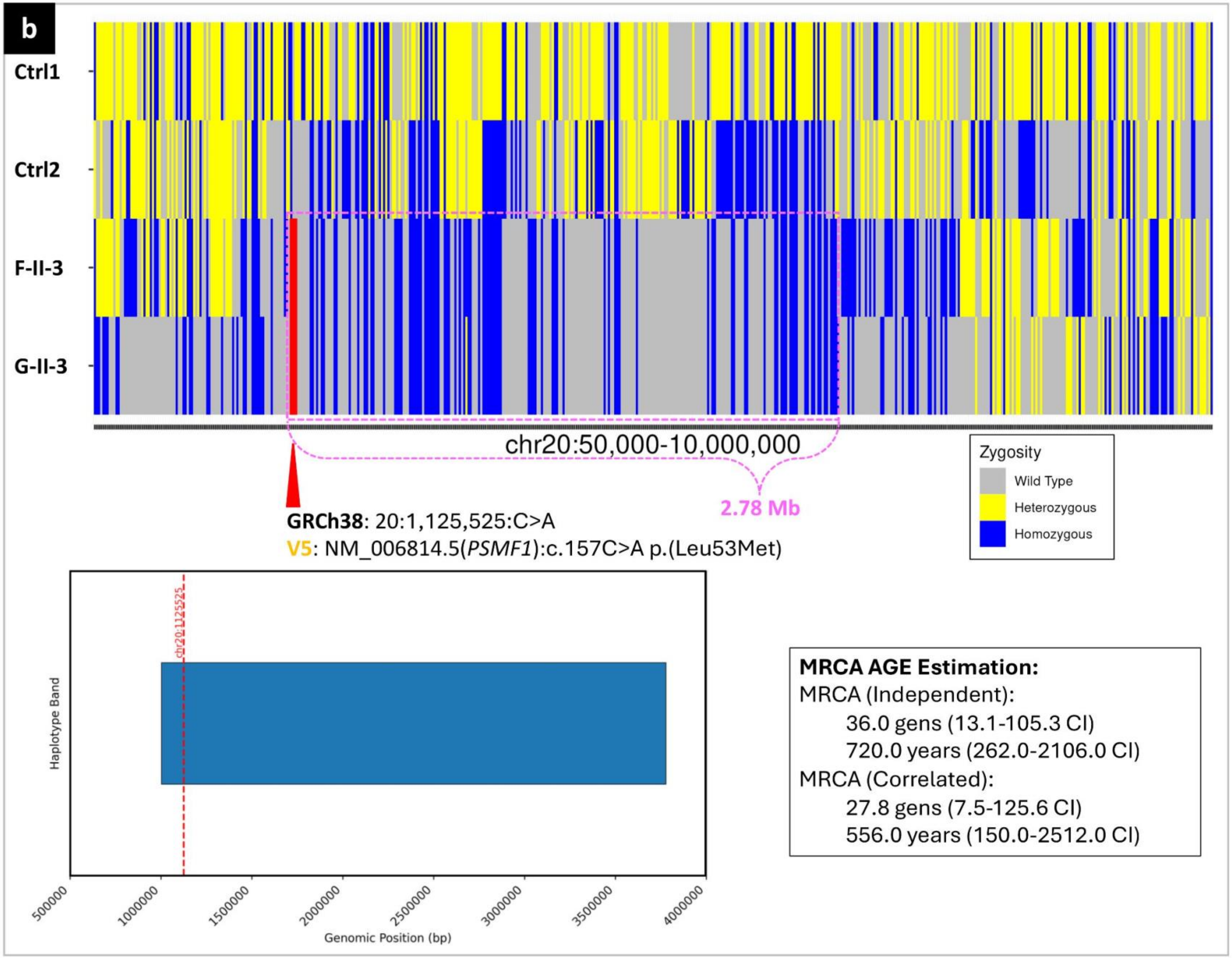
b
Ctrl1
Ctrl2
F-II-3
G-II-3
chr20:50,000-10,000,000
2.78 Mb
GRCh38: 20:1,125,525:C>A
V5: NM_006814.5(PSMF1):c.157C>A p.(Leu53Met)
Zygosity
Wild Type
Heterozygous
Homozygous
chr20:1125525
Haplotype Band
500000
1000000
1500000
2000000
2500000
3000000
3500000
4000000
Genomic Position (bp)
MRCA AGE Estimation:
MRCA (Independent):
36.0 gens (13.1-105.3 CI)
720.0 years (262.0-2106.0 CI)
MRCA (Correlated):
27.8 gens (7.5-125.6 CI)
556.0 years (150.0-2512.0 CI)

170

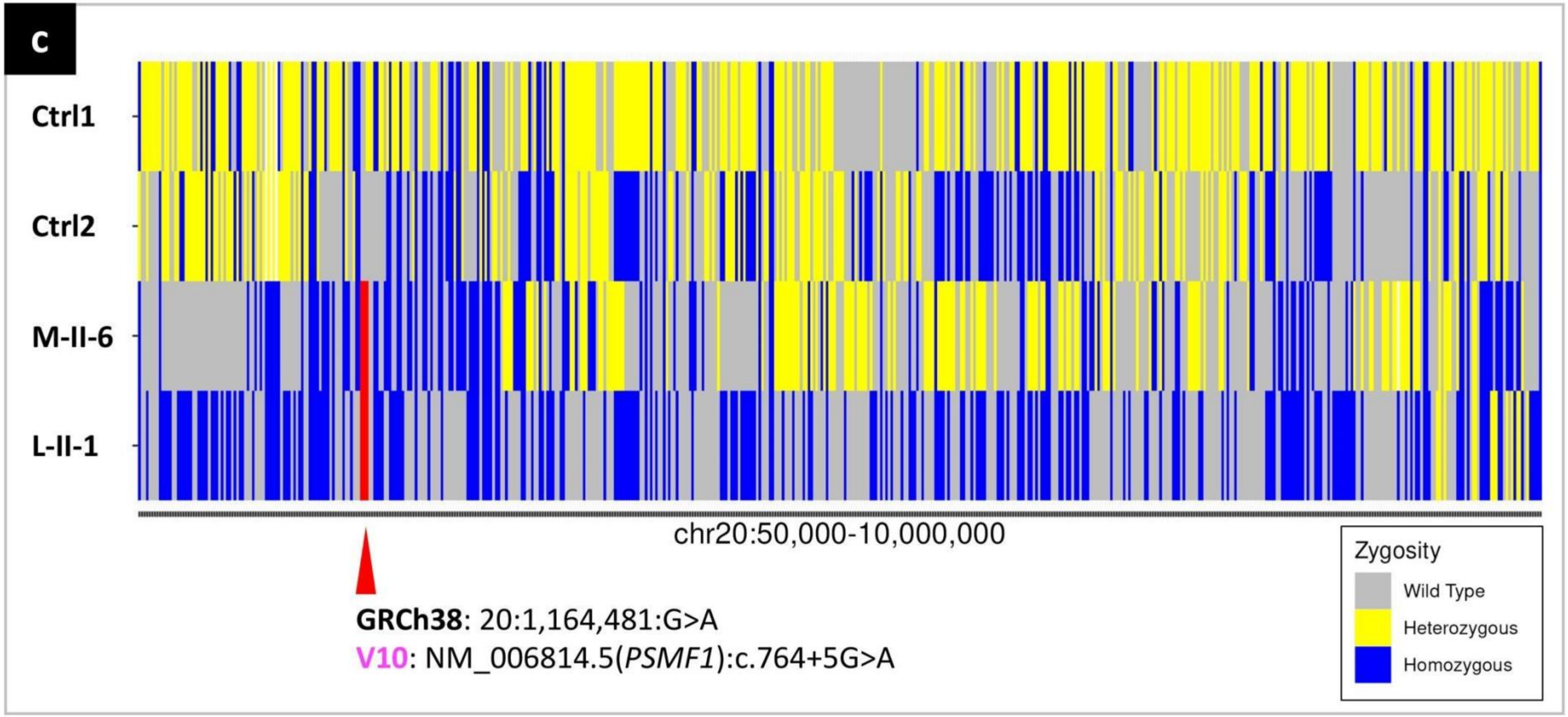


**Legend:** Haplotype analysis of the *PSMF1* (NM_006814.5) variants c.724C>G p.(Arg242Gly), c.157C>A p.(Leu53Met) and c.764+5G>A which were detected in the homozygous state in the affected individuals of Pedigrees B-C (B-II-3, B-II-4, C-II-3), Pedigrees F-G (F-II-3, G-II-3), and Pedigrees L-M (L-II-1, M-II-6; Fig.1a), respectively. Color banding identifies the *PSMF1* variant under investigation in red, homozygous variants in blue, heterozygous variants in yellow and wild-type bases in gray. **(a-c)** Different pattern of variants flanking the *PSMF1* variants c.724C>G p.(Arg242Gly) in affected individuals from Pedigrees B-C and c.764+5G>A in probands from Pedigrees L-M. **(b)** The same pattern of variants flanking the *PSMF1* variant c.157C>A p.(Leu53Met) in probands F-II-3 and G-II-3 identified a homozygosity block of 2.78 Mb (fuchsia dashed rectangle) inherited from a distant common ancestor, with the founder variant emerging between 720 to 556 years ago. Genomic coordinates are based on the human reference genome assembly GRCh38/hg38. Ctrl = control; gens = generations; GRCh38/hg38 = Genome Reference Consortium Human Build 38 Organism: Homo sapiens (human); MRCA = most recent common ancestor.

**Supplementary File 4. AlphaFold 3 modelling of PSMF1-FBXO7 complex**

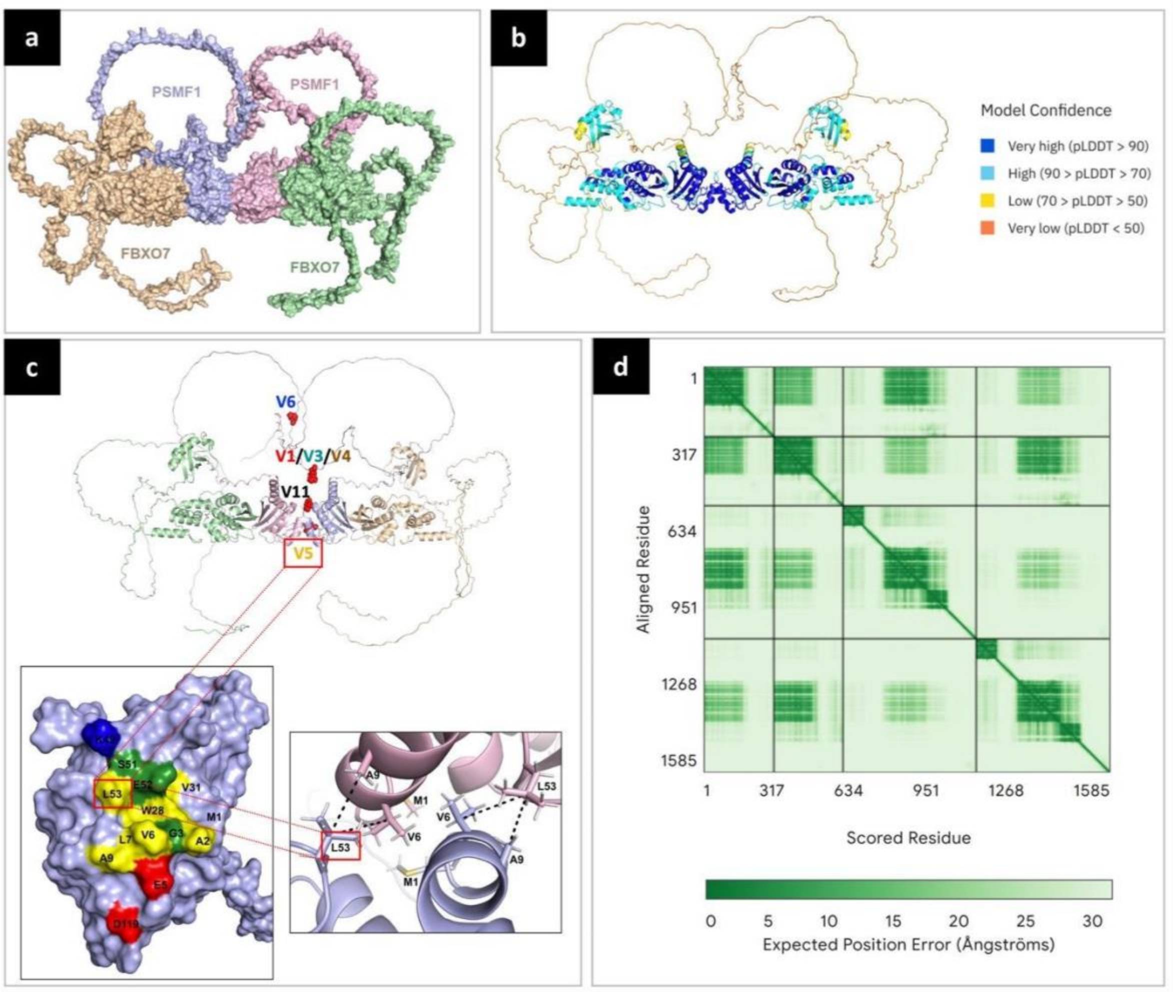


**Legend:** **(a)** AlphaFold 3 modelling of PSMF1 (UniProt: Q92530) and FBXO7 (Q9Y3I1-1) binding predicted that these two proteins form a heterotetramer.[5] The predicted structures of the models obtained were visualized and analyzed using PyMOL 2.5.5. Predictions consistently showed the same heterodimerization interfaces engaged in binding between PSMF1 and FBXO7 as well as the same homodimerization interfaces of PSMF1, with **(b)** very high pLDDT scores. When analyzing *PSMF1* missense and start-loss variants identified in the study cohort, none of the affected amino acid residues lies at the interface of PSMF1 and FBXO7 binding, which makes unlikely for these variants to impact the PSMF1-FBXO7 interaction. The residues p.Leu228 and p.Arg242 are located within the disordered C-terminal region of PSMF1, which makes their impact on binding other proteins difficult to predict. On the contrary, **(c)** the residue p.Leu53 sits on the dimerization interface of PSMF1 (Fig.2c), forming direct hydrophobic interactions with the residues p.Val6 and p.Ala9 of the second PSMF1 monomer. Based on this prediction, the PSMF1 residue p.Leu53Met (V5 in Fig.1a and Supplementary File 1a; Fig.2a-b-c) identified in Pedigrees F-G could disrupt the binding within the PSMF1 homodimer. pLDDT = predicted local distance difference test (per-residue measure of local confidence; score: 0-100, with higher scores indicating higher confidence and usually a more accurate prediction); *PSMF1* variants (NM_006814.5): V1 = c.724C>G p.(Arg242Gly); V3 = c.724C>T p.(Arg242Cys); V4 = c.725G>A p.(Arg242His); V5 = c.157C>A p.(Leu53Met); V6 = c.682C>G p.(Leu228Val); V11 = c.1A>T. **(d)** The Predicted Aligned Error (PAE) plot depicts the predicted alignment error for each residue pair in the model of the PMSF1-FBXO7 complex model generated by AlphaFold 3. Lower PAE values (green) indicate higher confidence in the relative positioning of residue pairs, while higher values (white) reflect greater uncertainty. The diagonal represents intra-chain confidence, whereas off-diagonal blocks reflect inter-chain interaction confidence between PMSF1 and FBXO7. High-confidence interaction regions are visible as dark green off-diagonal areas, suggesting a well-defined interface between the two proteins.

**Supplementary File 5. Quantitative proteomic analysis of mitochondrial and lysosomal proteins in *PSMF1* patient- and carrier-derived primary dermal fibroblasts.**

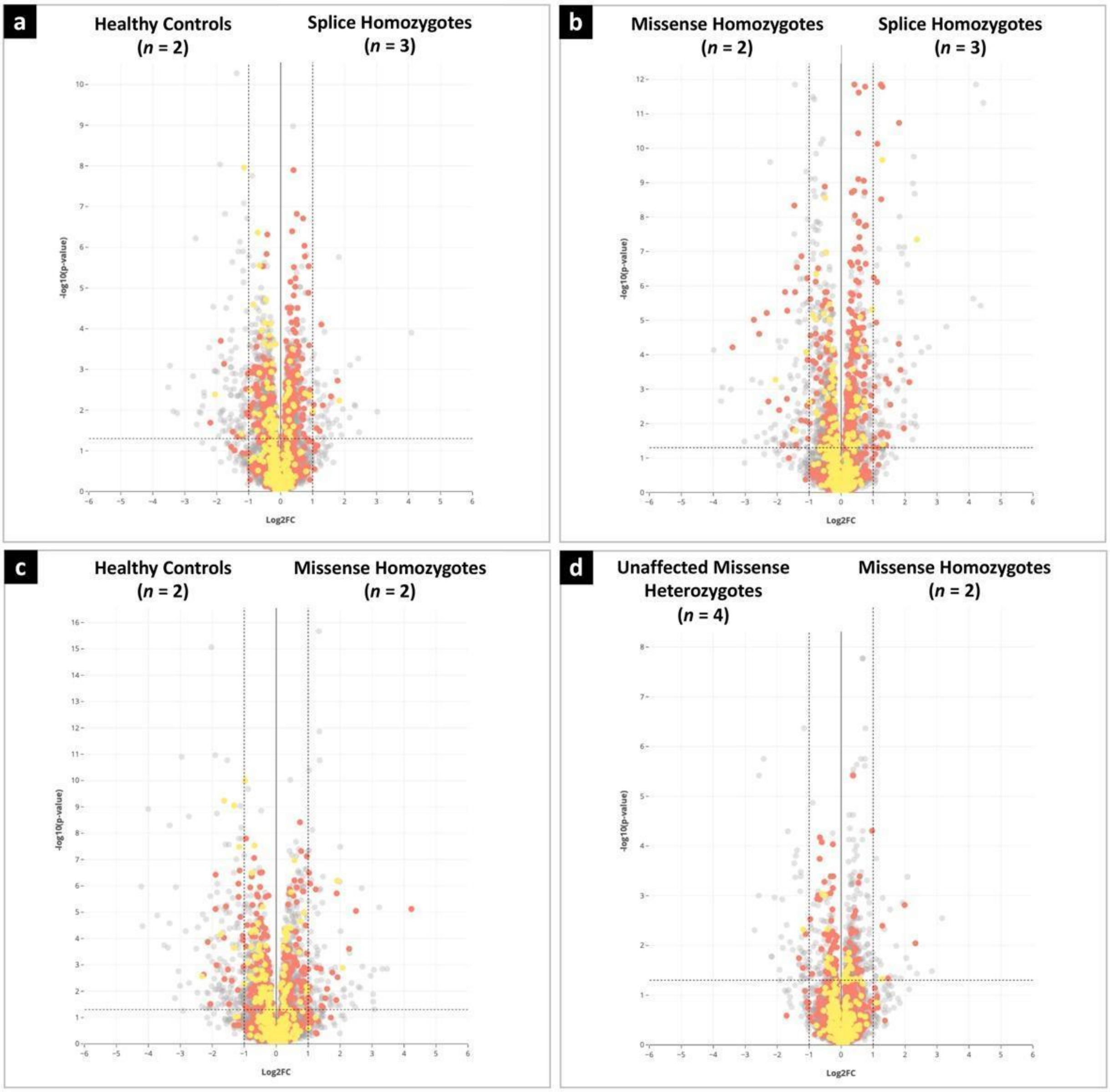


**Legend.** Volcano plots displaying enrichment of mitochondrial and lysosomal proteins. Further experiments including undertaking organelle enrichment studies are needed to comprehensively elucidate the mechanistic impacts of *PSMF1* gene on protein abundance regarding proteasomal, mitochondrial and other relevant pathways due to the high number of mitochondrial/lysosomal-associated proteins detected within the dataset. Groups: splice homozygotes (*n* = 3; J-II-3, L-II-4, M-II-6), missense homozygotes (*n* = 2; B-II-3, F-II-3), unaffected missense heterozygotes (*n* = 4; A-I-1, B-II-1, F-I-1, F-I-2), healthy controls (HC).

**Supplementary File 6. Frequency of *PSMF1* variants in the UK Biobank (UKB) and the Accelerating Medicines Partnership program for Parkinson's disease (AMP-PD) and gene burden analysis**

We searched for *PSMF1* missense (nonsynonymous, nonframeshift indels) and loss-of-function (splice, nonsense, frameshift) variants in the ES data within the UK Biobank (UKB, https://www.ukbiobank.ac.uk/)[6] and in the GS data within the Accelerating Medicines Partnership-Parkinson's Disease Initiative (AMP-PD, https://amp-pd.org/) in September 2023.[7] By setting minor allele frequency (MAF) < 0.005, we were able to capture most *PSMF1* variants, as documented by the histograms below.

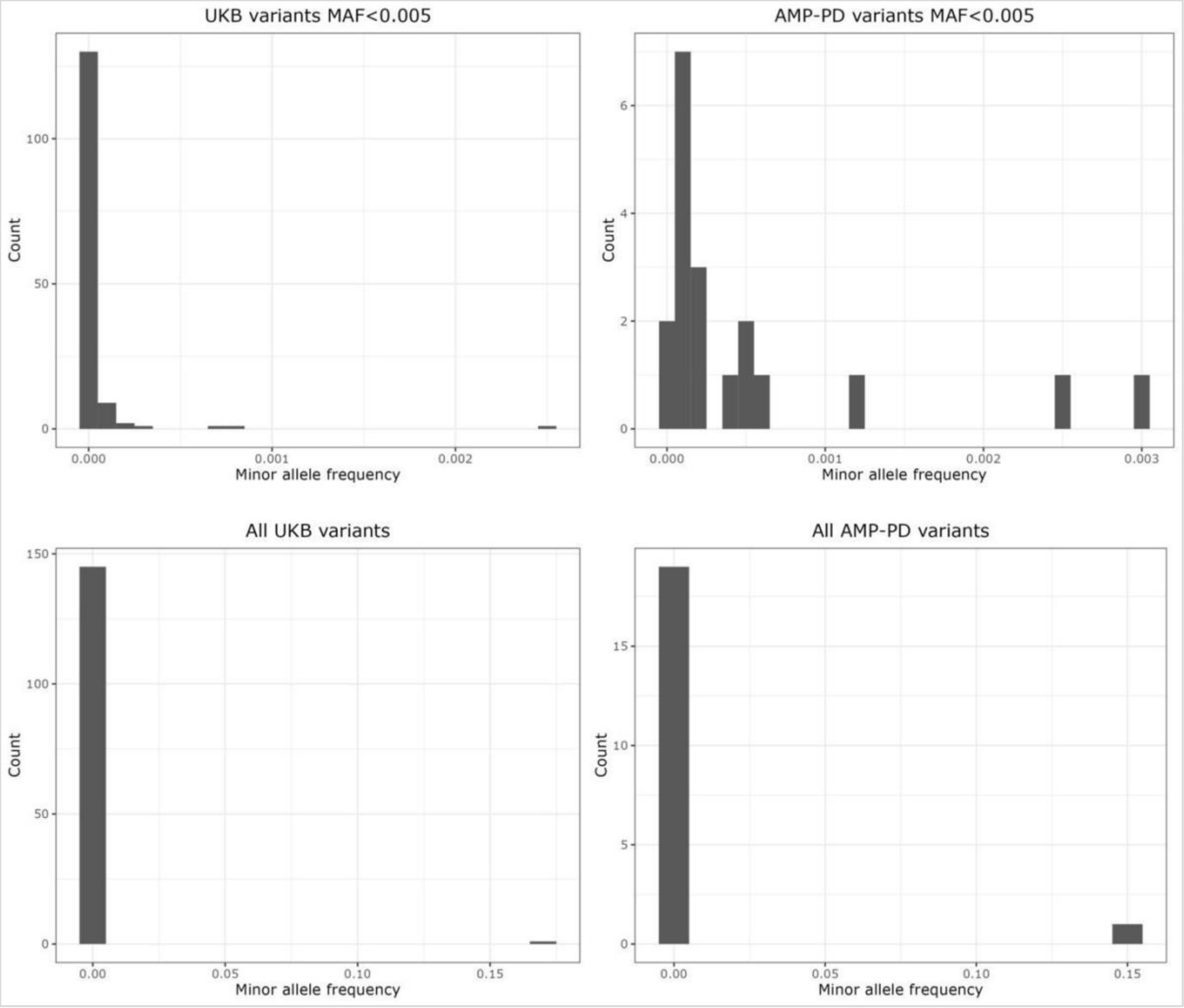


*PSMF1* was not significantly associated with Parkinson's disease (PD) in the UKB ES data and the AMP-PD GS data, and gene burden analysis revealed that all p values for *PSMF1* in the different categories were > 0.05 (not statistically significant).

In UKB, 45,857 people were included (38,051 controls, 1,105 cases, and 6,701 proxies comprised of 6,033 parents and 668 siblings). Using a MAF limit of 0.005, we found 131 missense and 14 loss-of-function variants in *PSMF1*. Single variant association testing (using the --model flag), revealed no association with PD in a recessive model. Specifically, we did not identify any individual who was homozygous alternate for any *PSMF1* variant. In this dataset, 560 individuals carried heterozygous non-reference alleles for 1/131 variants, and 2 individuals were heterozygotes for 2/131 variants. Overall, these 562 heterozygous carriers consist of 468 controls and 94 cases, resulting in a frequency of 0.83 for controls and 0.17 for cases, resulting in a 0.66 difference between cases and controls. Using the Sequencing Kernel Association optimal unified test (Skat-O), Combined Multivariate and Collapsing (CMC) Wald test and CMC burden testing, *PSMF1* resulted in non-significant p values (all *PSMF1* missense variants as defined by ANNOVAR: 0.91 for Skat-O, 0.84 for CMCWald, and 0.84 for CMC; all *PSMF1* missense variants as defined by SnpEff: 0.91 for Skat-O, 0.84 for CMCWald, and 0.84 for CMC). As for *PSMF1* loss-of-function variants in the UKB dataset, we did not identify any homozygous individual for *PSMF1* non-reference alleles in any of the 14 variants. There were 14 heterozygous individuals for 1/19 variants in *PSMF1*. Those 14 heterozygous carriers consisted of 12 controls and 2 cases, resulting in a frequency of 0.86 for controls and 0.14 for cases, corresponding to a 0.72 difference between cases and controls. Using Skat-O, CMCWald, and CMC burden testing, *PSMF1* resulted in non-significant p values (all *PSMF1* loss-of-function variants as defined by ANNOVAR: 0.78 for Skat-O, 0.98 for CMCWald, and 0.98 for CMC; all *PSMF1* loss-of-function variants as defined by SnpEff: 0.48 for Skat-O, 0.30 for CMCWald, and 0.30 for CMC).

In the AMP-PD, 4,007 people were included (2,556 controls, 1,451 cases). Using an upper MAF of 0.005, we found 19 *PSMF1* missense and no *PSMF1* loss-of-function variants. Single variant association testing (using the --model flag) revealed no associations with PD in a recessive model. As for *PSMF1* variants, we did not identify any individual with a homozygous *PSMF1* non-reference allele in 19 missense variants detected, whereas 83 individuals were heterozygous for 1/19 variants. Those in total 83 heterozygous carriers consisted of 51 controls and 32 cases, resulting in a frequency of 0.61 for controls and 0.39 for cases, thus corresponding to a 0.22 difference between cases and controls. Using Skat-O, CMCWald, and CMC burden testing, *PSMF1* resulted in non-significant p values (all *PSMF1* missense variants as defined by ANNOVAR: 0.21 for Skat-O, 0.95 for CMCWald, and 0.95 for CMC; all *PSMF1* missense variants as defined by SnpEff: 0.27 for Skat-O, 0.572 for CMCWald, and 0.72 for CMC).

**Supplementary File 7. Detection of α-synuclein seeding activity by CSF and skin real-time quaking-induced conversion assay in individuals with biallelic *PSMF1* variants**

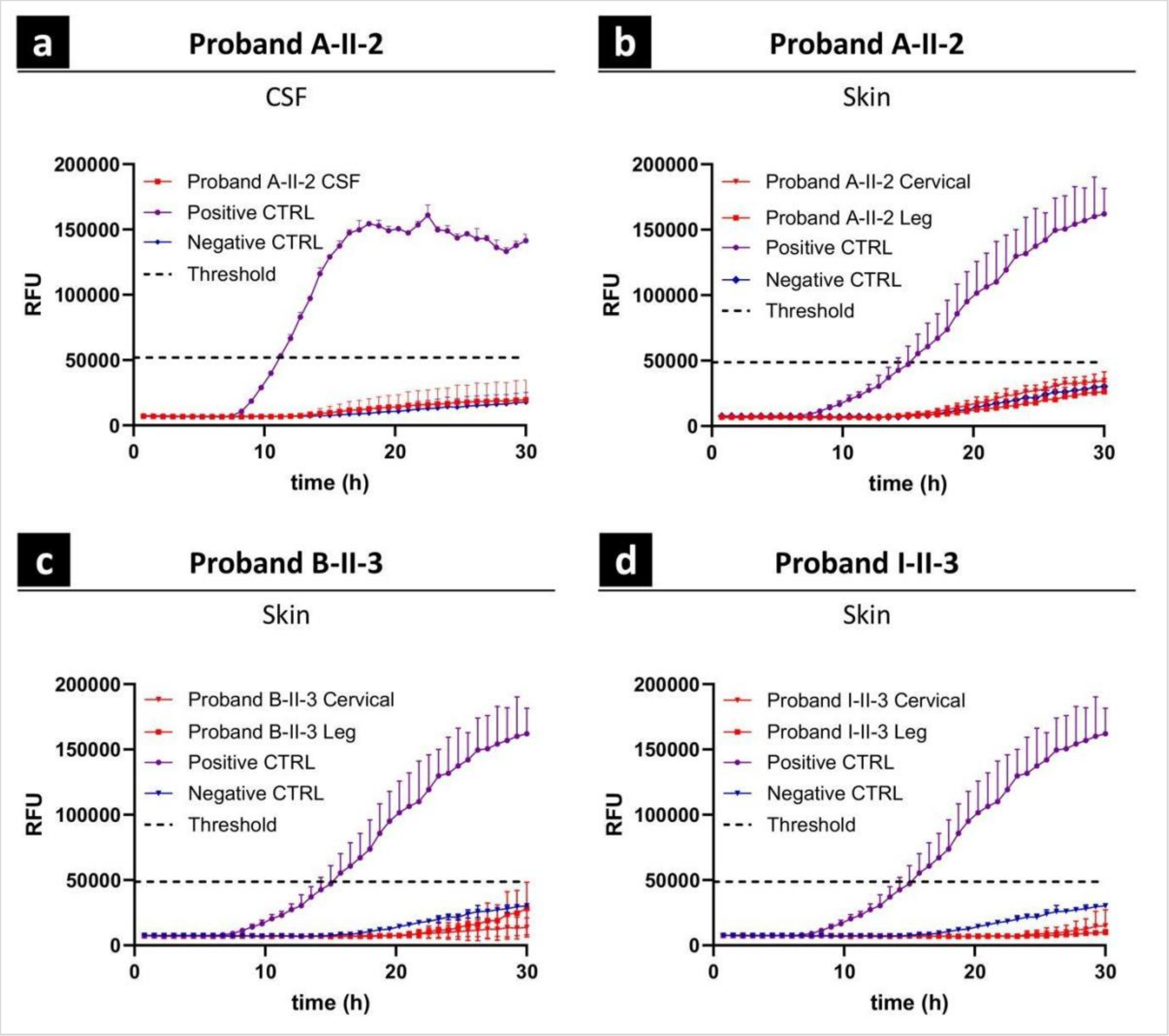


**Legend:** RT-QuIC analysis of CSF from proband A-II-2 **(a)** and skin from probands A-II-2 **(b)**, B-II-3 **(c)** and I-II-3 **(d)** compared to CSF and skin samples from two positive and two negative control subjects, respectively. Skin biopsies were performed from the C7-C8 dermatomes (cervical) and from the distal lower limb, 10 cm proximally to the lateral malleolus (leg). Each curve represents the mean fluorescence of four replicates for *PSMF1* probands' samples and the group mean ($n$ = 2) for the control samples. Error bars indicate the standard deviation. The black dashed line represents the positivity threshold, arbitrarily set at 30% of the median $I_{max}$ value of the positive control replicates. CTRL = control; h = hours; RFU = relative fluorescence units; RT-QuIC = real-time quaking-induced conversion.

**Supplementary File 8. Native gel electrophoresis and Western blotting for proteasome assays**

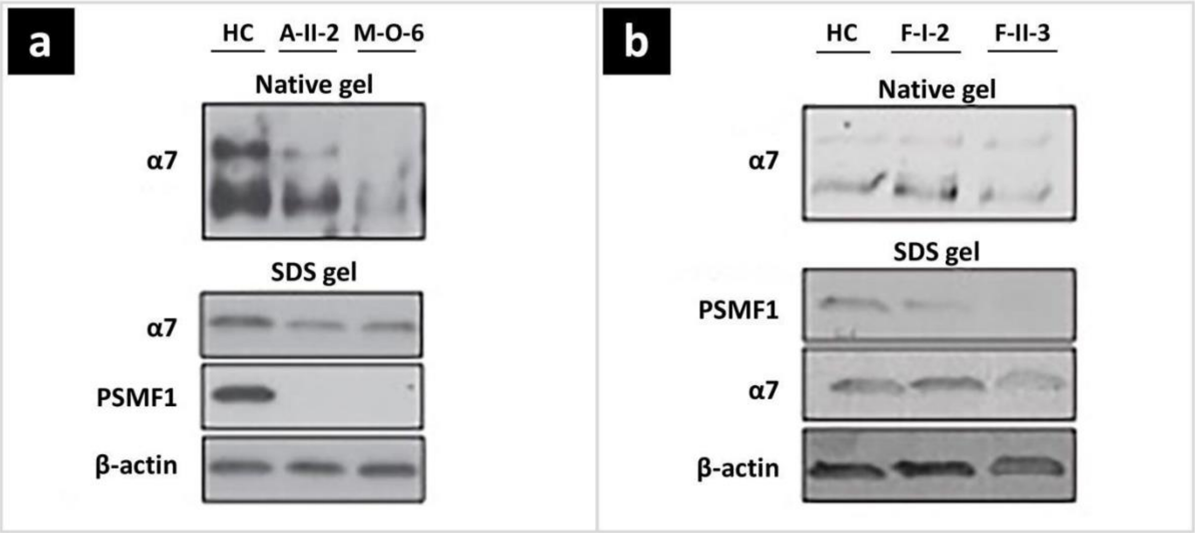


**Legend: (a)** Native gel assays showing a reduction in the amount of both 20S and 26S proteasome particles in *PSMF1* proband-derived fibroblasts (A-II-2, M-O-6) compared to a healthy control, as well as a reduction in the ratio of assembled 26S particles. **(b)** Alpha-7 subunit immunoreactivity was reduced in fibroblast samples from the *PSFM1* proband F-II-3 by both native gel and Western blotting analyses.

**Supplementary File 9. TdTomato (red) expression as marker of recombination in brain regions of *Psmf1*$^{fl/fl}$ Ai14 UBC-Cre-ERT2 mice**

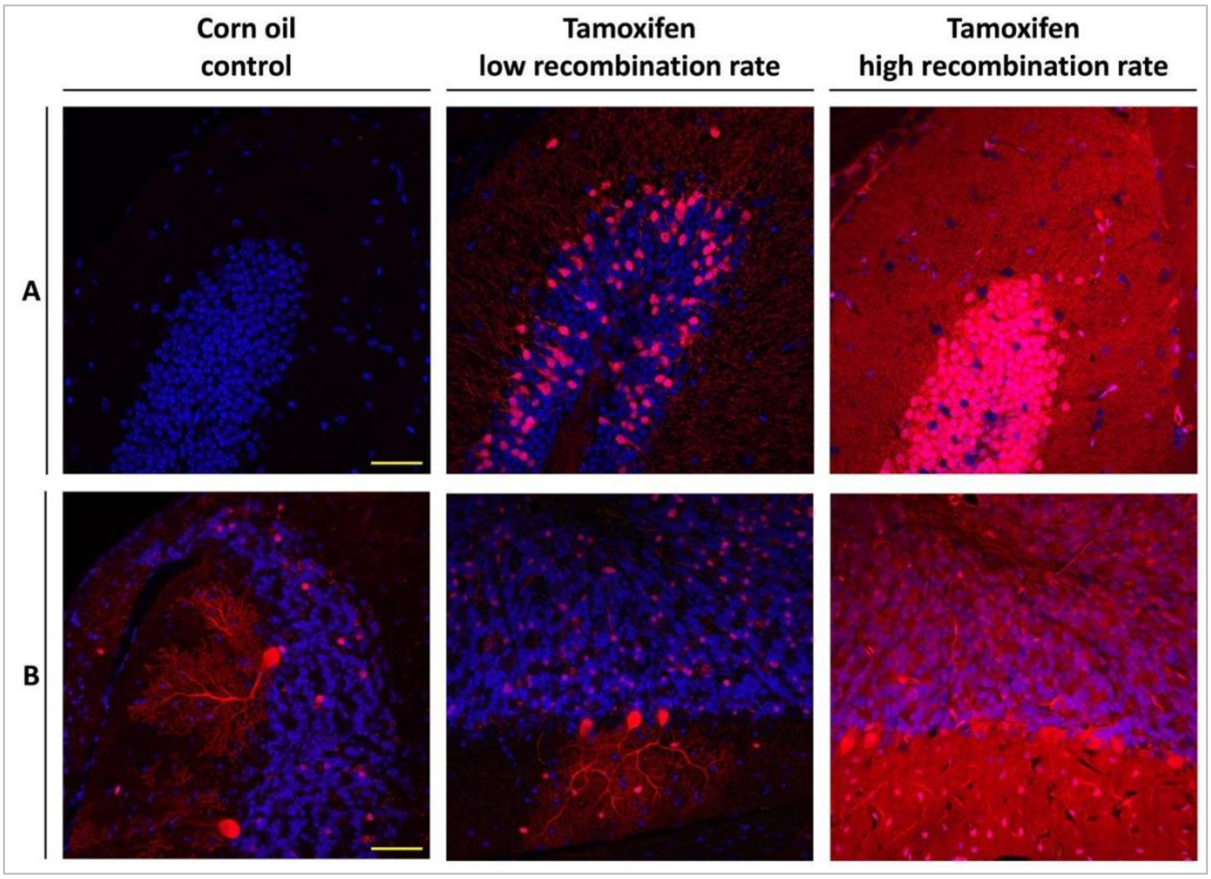


**Legend:** Tamoxifen-induced recombination in *Psmf1*$^{fl/fl}$ Ai14 UBC-Cre-ERT2 mice induces tdTomato (red) expression, which marks cells in which *Psmf1* is inactivated. Nuclei are stained with Hoechst 33342 (blue). Panels from left to right: control (corn oil), low recombination (tamoxifen; 30-50%) and high recombination (tamoxifen; ~90%). (A) Hippocampus; (B) Cerebellum. Scale bars indicate 50 µm.

**Supplementary File 10. Immunohistochemistry of different brain regions of *Psmf1*$^{fl/fl}$ Ai14 UBC-Cre-ERT2 mice 28 days after tamoxifen injection**

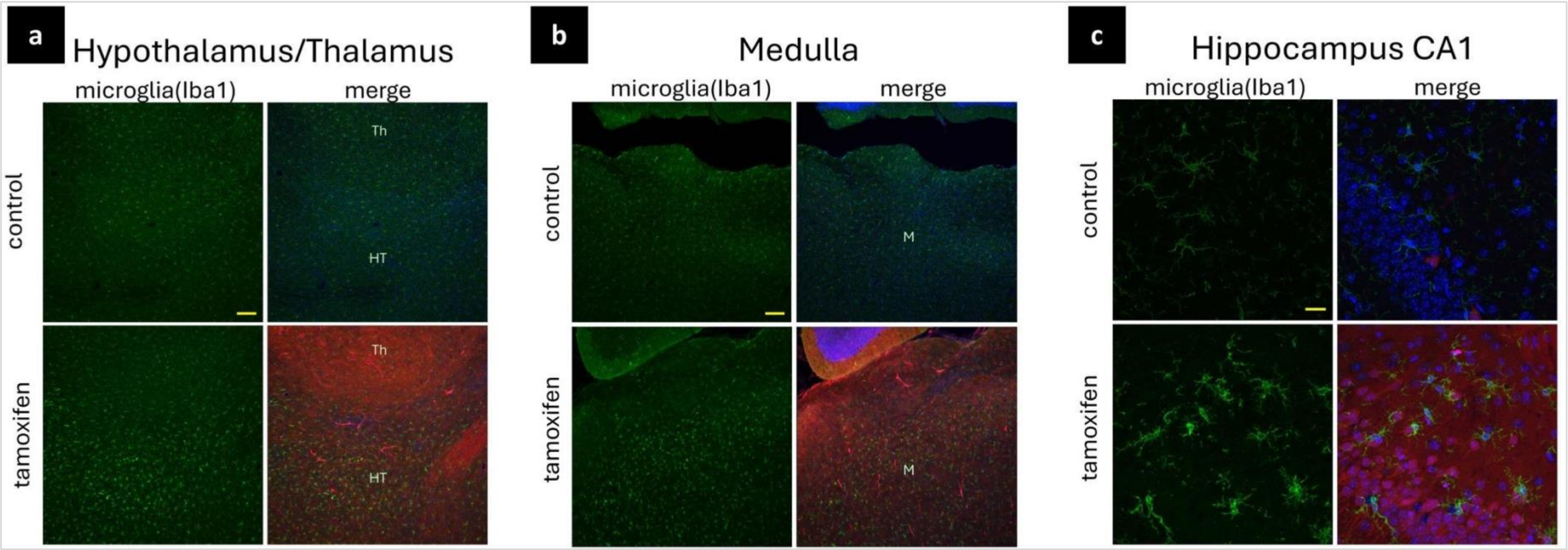


**Legend:** *Psmf1*$^{fl/fl}$ Ai14 UBC-Cre-ERT2 mice were injected with either corn oil (control) or tamoxifen to induce Cre recombination of floxed *Psmf1* exon 3 for loss of Psmf1 and a stop cassette in Ai14 for expression of tdTomato (red) to evaluate recombination efficiency. Mice were sacrificed 28 days after tamoxifen injection. Sagittal brain sections were stained with Iba1 (green) for microglia and Hoechst 33342 for nuclei (blue). An increase in reactive microglia was found in the **(a)** hypothalamus (HT) and **(b)** medulla (M) (scale bar 100 µm) as well as in the **(c)** hippocampus CA1 (scale bar 20 µm). Th = thalamus. TdTomato (red) fluorescence was used as indicator of recombination efficiency.

**Supplementary File 11. Immunohistochemistry of the substantia nigra and ventral tegmental area of *Psmf1*$^{fl/fl}$ Ai14 UBC-Cre-ERT2 mice 28 days after tamoxifen injection**

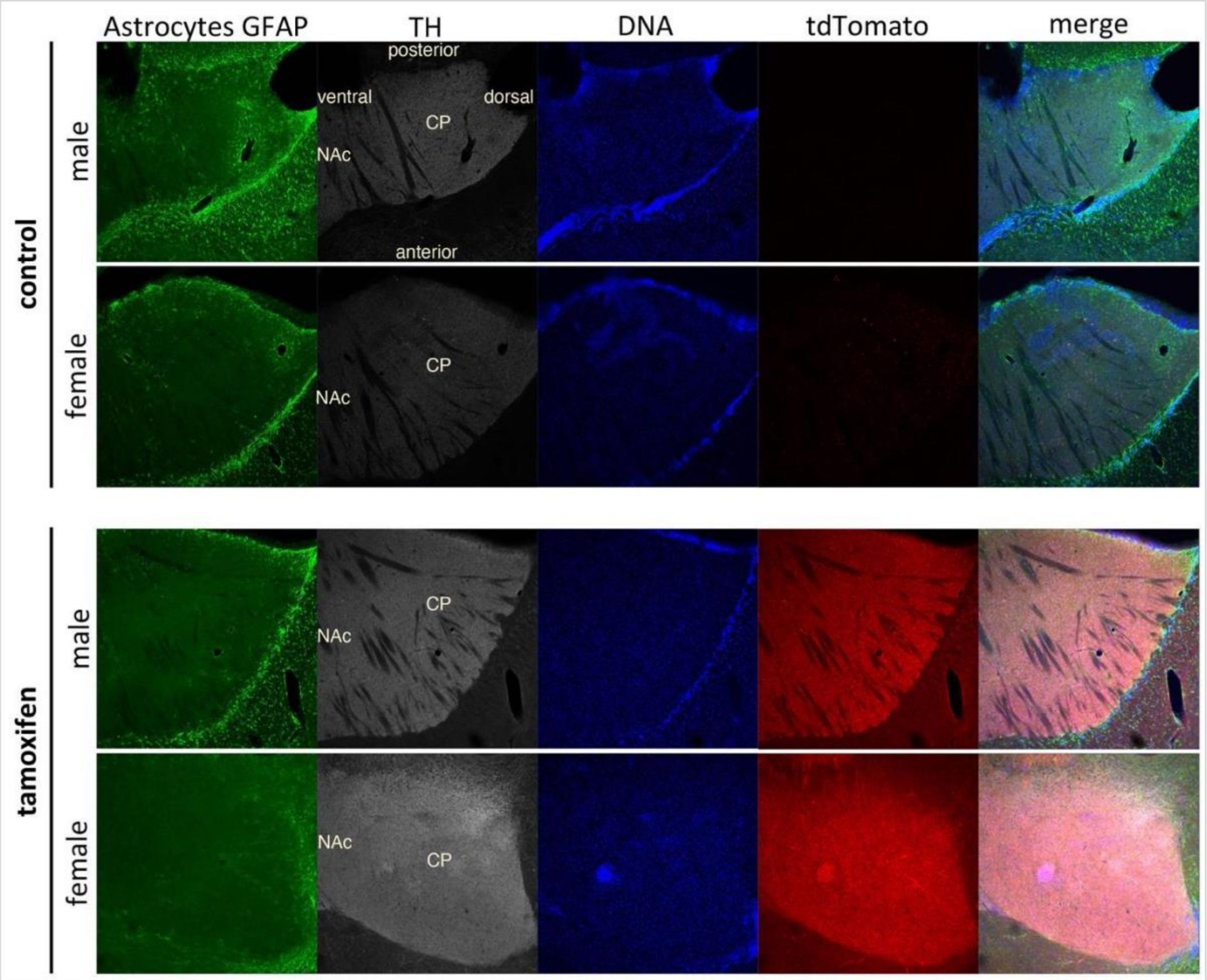


**Legend:** At Day 28 after tamoxifen injection, there was no evidence of increased gliosis at the axonal ends of tyrosine hydroxylase (TH)-positive dopaminergic neurons in the subcortex (striatum: nucleus accumbens [NAc] and caudate putamen [CP]) of *Psmf1*$^{fl/fl}$ Ai14 UBC-Cre-ERT2 mice, as reflected by lack of immunoreactivity for glial fibrillary acidic protein (GFAP) in astrocytes and Iba1 in microglial cells. This could reflect that mice were sacrificed too early for dopaminergic neurons to exhibit pathological phenotype. Nuclei (DNA) were stained with Hoechst 33342 (blue). TdTomato (red) fluorescence was used as indicator of recombination efficiency.

**Supplementary File 12. List of primers used for the splicing (minigene) assay of *PSMF1* variants c.365+2T>C and c.605+1G>A (NM_006814.5)**

| **Primer name** | **Primer sequence (5'-3')** | **Purpose** |
|---|---|---|
| hu_PSMF1_Ex3_XhoI_F | aattctcgagATCTCCGTGTTGCTCACCTG | Amplification of gDNA and cloning |
| hu_PSMF1_Ex3_BamHI_R | attggatccGTGGACTCTTGACTGCACCA | Amplification of gDNA and cloning |
| hu_PSMF1_Ex5_XhoI_F | aattctcgagTAGGTTTGTGTTCCGCCCTT | Amplification of gDNA and cloning |
| hu_PSMF1_Ex5_BamHI_R | attggatccACCGGGAAATGGCAGAAGAG | Amplification of gDNA and cloning |
| PSMF1 c.365+2T_C Mut F | CTTCCACAGGcACTTCTAAATGATGTC | Site-directed mutagenesis |
| PSMF1 c.365+2T_C Mut R | TCACCCAGGTGTTCTGCA | Site-directed mutagenesis |
| PSMF1 c.605+1G_A Mut F | ACCCTTTTGGaTGAGTACTGC | Site-directed mutagenesis |
| PSMF1 c.605+1G_A Mut R | CTAAGTCTTCTCCCCCGA | Site-directed mutagenesis |
| SD6 F | TCTGAGTCACCTGGACAACC | Colony PCR and RT-PCR |
| SA2 R | ATCTCAGTGGTATTTGTGAGC | RT-PCR |

**Legend:** gDNA = genomic DNA; PCR = polymerase chain reaction; RT-PCR = real-time (RT) PCR.

**Supplementary File 13. CURTAIN web links to proteomics datasets**

| Figure | Comparison | CURTAIN Web Link |
|---|---|---|
| **3a** | Splice Homozygotes vs HC | https://curtain.proteo.info/#/6fa3a44a-7708-424b-972b-1f10e585b78e |
| **3b** | Splice Homozygotes vs Missense Homozygotes | https://curtain.proteo.info/#/acc6f322-a5b8-498b-ac30-c1143add90c1 |
| **3c** | Missense Homozygotes vs HC | https://curtain.proteo.info/#/f96a5ff0-27ab-4392-93ce-b8f667ad6ea7 |
| **3d** | Missense Homozygotes vs Unaffected Missense Heterozygotes | https://curtain.proteo.info/#/6b5c0cb0-c151-41fe-a44b-0682b1aa8a9f |

| File | Comparison | CURTAIN Web Link |
|---|---|---|
| **Supplementary File 5a** | Splice Homozygotes vs HC | https://curtain.proteo.info/#/da26e84d-a928-41d2-8f09-0192c7576f46 |
| **Supplementary File 5b** | Splice Homozygotes vs Missense Homozygotes | https://curtain.proteo.info/#/9e2152d9-9c7f-4ce0-8f28-e9074c642474 |
| **Supplementary File 5c** | Missense Homozygotes vs HC | https://curtain.proteo.info/#/6081d90f-31cf-4d6c-b43c-c4b22774f904 |
| **Supplementary File 5d** | Missense Homozygotes vs Unaffected Missense Heterozygotes | https://curtain.proteo.info/#/95f2c1f9-86f1-4d9f-bede-6acd7a0a338f |

**Legend:** HC = healthy controls.

**Supplementary material - References**
